# Evaluating genetically-predicted causal effects of lipoprotein(a) in human diseases: a phenome-wide Mendelian randomization study

**DOI:** 10.1101/2024.07.24.24310950

**Authors:** Jingchunzi Shi, Suyash S. Shringarpure, David Hinds, 23andMe Research Team, Adam Auton, Michael V Holmes

## Abstract

**Background:** Lipoprotein(a) (Lp[a]) is a circulating plasma lipoprotein that is emerging as an important independent risk factor for vascular disease. Lp(a) levels are 75-90% heritable, predominantly determined by copy number variation and single nucleotide polymorphisms (SNPs) at the *LPA* gene.

**Methods:** Using ∼370K individuals with serum measurements of Lp(a) in the UK Biobank European cohort, we constructed a genetic risk score (GRS) consisting of 29 SNPs in the vicinity of *LPA* which explained 68.18% of variation in Lp(a). Using the *LPA* GRS to instrument Lp(a), we conducted phenome-wide Mendelian randomization analysis (MR-PheWAS) across a spectrum of 489 medically-relevant phenotypes in ∼7.3M individuals from the 23andMe, Inc. database, and compared effects to those derived from a GRS for low-density lipoprotein cholesterol (LDL-C) and apolipoprotein B (apoB). Through multivariable MR, we sought to assess the direct causal effect of Lp(a) on cardiovascular disease risks while keeping LDL-C or apoB constant.

**Results:** MR-PheWAS confirmed previously reported Lp(a) causal effects on coronary artery disease (CAD: OR = 1.199, 95% CI = [1.193, 1.205], p-value < 2.23×10^-308^, for every 59.632 nmol/L higher Lp(a) instrumented by the *LPA* GRS), and revealed additional genetically-predicted effects largely confined to cardiovascular endpoints, including a novel effect for restrictive cardiomyopathy (OR = 1.101, 95% CI = [1.068, 1.134], p-value = 3×10^-10^). We scaled the *LPA*, LDL-C and apoB GRS such that they each had the same OR for MACE (major adverse cardiovascular events). Using the scaling rubric, similar magnitudes of effect were seen for the three lipid traits for most vascular diseases, with the exception of peripheral artery disease, aortic stenosis and dilated cardiomyopathy, where Lp(a) had larger genetically–predicted effect sizes compared to LDL-C and apoB. Multivariable MR identified Lp(a) to retain a causal effect on MACE while accounting for LDL-C or apoB. To achieve the 25% relative risk reduction in major vascular events, as seen with a 1 mmol/L reduction in LDL-C from statin trials, we anticipate that Lp(a) ought to be reduced by ∼ 90 mg/dL (200 nmol/L), highlighting the importance of not only using therapies that have a profound impact on Lp(a) lowering, but also selecting individuals that have high Lp(a) concentrations at baseline.

**Conclusion:** Lp(a) has genetically-predicted causal effects on a broad range of cardiovascular diseases beyond CAD, with minimal effects seen for non-vascular disease.

## Introduction

Lipoprotein(a) - Lp(a) - a circulating plasma lipoprotein, is formed by a cholesterol-rich low-density lipoprotein (LDL) particle that is covalently bonded with the glycoprotein apolipoprotein(a) (apo[a]). Epidemiological and genomic studies have established strong and consistent evidence to associate elevated plasma Lp(a) levels with increased risk of coronary heart disease (CHD), peripheral vascular disease, heart failure (HF), and aortic stenosis (Emerging Risk Factors Collaboration, 2009; Emdin, C.A. *et al*., 2016; Arnett, D.K. *et al*., 2019), with consequent development of Lp(a)-lowering therapies in ongoing phase III cardiovascular outcome trials (ClinicalTrials.gov ID: NCT04023552, NCT05581303).

Lp(a) is distinguished from other cardiovascular disease (CVD) risk factors by its pronounced almost oligogenic genetic architecture. Previous studies have reported Lp(a) concentration to be 75% to 90% heritable, with the variability primarily determined by single nucleotide polymorphisms (SNPs) and a variable number of KIV (kringle IV) repeats at the *LPA* gene (Boerwinkle, E. *et al*., 1992; Clarke, R. *et al*., 2009; Zekavat, S.M. *et al*., 2018). These features have enabled construction of *LPA* genetic risk scores (GRS) which explained as much as 60% of the variability in Lp(a) levels among individuals of European genetic ancestry (Burgess, S. *et al*., 2018; Trinder, M. *et al*., 2021). In contrast, genome-wide GRS for the lipid trait low-density lipoprotein cholesterol (LDL-C) explains only 11-17.5% of the variability (Graham, S.E. *et al*., 2021; Sinnott-Armstrong, N. *et al*., 2021; Privé, F. *et al*., 2022; Weissbrod, O. *et al*., 2022).

Despite public health and pharmaceutical interest in Lp(a), evidence gaps remain in the understanding of the causal effects and therefore potential clinical utility of Lp(a) lowering therapies. Specifically, although *LPA* variants have consistently indicated a causal relationship with CVD (Clarke, R. *et al*., 2009; Mack, S. *et al*., 2017; Saleheen, D. *et al*., 2017; Burgess, S. *et al*., 2018; Zekavat, S.M. *et al*., 2018; Said, M.A. *et al*., 2021), the characterization of Lp(a) on a full spectrum of disease endpoints, especially the pathogenesis of non-CVD phenotypes, remains largely unexplored (Mora, S. *et al*., 2010; Sawabe, M. *et al*., 2012; Emdin, C.A. *et al*., 2016). Moreover, while statins, PCSK9 inhibitors and ezetimibe are proven to effectively lower LDL-C levels, a well-recognized major aetiological determinant of CVD, evidence indicates that a substantial residual burden of CVD remains (Cohen, J.C. *et al*., 2006; Rohatgi, A. *et al*., 2014; Degoma, E.M. *et al*., 2016; Nicholls, S.J. *et al*., 2016; Sabatine, M.S., 2019; Groenen, A.G., *et al*., 2021; Tall, A.R. *et al*., 2022). Recently, the causal role of apolipoprotein B (apoB) has gained traction as providing insight into why some lipid modifying therapeutic targets may differ in their vascular disease associations (Ference, B.A. *et al*., 2017; Ference, B.A. *et al*., 2019). Importantly, each Lp(a) particle includes an LDL and an apoB particle meaning that when interpreting the effects of Lp(a) on CVD risk, consideration ought also be placed on these LDL and apoB constituents. For example, the commonly used LDL-C assays also measure the cholesterol in Lp(a). Whilst it is eminently plausible that Lp(a) plays a causal role in CVD risk independent of LDL and apoB, it is also feasible, that, once accounting for the structural properties of Lp(a), the causal effects of Lp(a) attenuates. Elucidating the extent to which the relationship between Lp(a) and CVD risk is independent of changes in LDL-C and apoB level has important ramifications in strengthening our understanding of the etiology of CVD. Furthermore, a recent study suggested that Lp(a) has a six-fold larger causal effect per particle basis on CHD as compared to comparable changes in apoB concentration (Björnson, E. *et al*., 2024). Therefore, a better understanding of the comparable causal estimates of these lipoprotein and lipid entities on CVD disease endpoints is needed.

The human genome is valuable in helping estimate the probable efficacy and safety of pharmacological modulation (Plenge, R.M. *et al*., 2013; Nelson, M.R. *et al*., 2015; Vallabh Minikel, E. and Nelson, M.R., 2023). In this study, we aimed to use human genetics to: 1) systematically investigate the causal role of Lp(a) on a phenome-wide basis, to shed light on a broader spectrum of potential beneficial and adverse consequences; 2) compare the relative magnitude of disease associations of Lp(a) versus LDL-C and versus apoB, while calibrating their effect estimates to the same risk reduction in a composite cardiovascular outcome; and 3) distinguish the direct effect of Lp(a) on CVD, after accounting for LDL-C or apoB. In order to accomplish those goals, we first constructed *LPA*, LDL-C and apoB GRS using the UK Biobank (UKB) blood biochemistry biomarker data. Next, we assessed the causal role of Lp(a) across a spectrum of 489 phenotypes from the 23andMe database using two-sample univariable Mendelian randomisation (MR). For any disease outcome that the univariable MR suggested Lp(a) being of putative causal relevance, we obtained the corresponding causal estimates of LDL-C and apoB instrumented using their respective GRS. In order to derive a clinically meaningful comparison between the magnitude of disease associations for Lp(a) vs LDL-C vs apoB, we defined a composite endpoint (which we denoted as major adverse cardiovascular endpoint, or, MACE) derived from the constituents of the primary efficacy endpoints of ongoing clinical trials (ClinicalTrials.gov ID: NCT04023552). We scaled the Lp(a), LDL-C and apoB GRS such that their univariable MR effect sizes on the MACE composite endpoint were the same. We then applied the MACE-derived scaling factors to Lp(a), LDL-C and apoB GRS respectively for their univariable MR analyses among other phenotypes with positive Lp(a) MR findings, to anchor their effects and to facilitate comparison across other diseases. Finally, through multivariable Mendelian randomisation, we sought to assess the direct causal effect of Lp(a) on MACE, accounting for genetically-predicted changes in LDL-C or apoB.

## Methods and Materials

### UK Biobank Participants

A detailed description of the UKB cohort can be found in Sudlow, C. *et al*., 2015. In short, the UKB is a population-based prospective cohort conducted in the United Kingdom. The study recruited approximately 500,000 participants aged between 40 and 69 years from 2006 to 2010. All participants provided informed consent. Details of participant data are available at https://biobank.ndph.ox.ac.uk/showcase. This research was conducted using the UK Biobank data under the approved application number 95801.

### UK Biobank Genotyping and Imputation

UKB genetic data were assayed with one of the two custom Affymetrix Axiom arrays. The genotype data were then phased and imputed against the Haplotype Reference Consortium (HRC) panel, augmented by the merged UK10K and 1000 Genomes phase 3 reference panels for variants that are not present in HRC. Detailed descriptions of the UKB genotyping, imputation, and quality control procedures can be found in Bycroft, C. *et al*., 2018.

### UK Biobank Blood Biochemistry Biomarker Measurements

Lp(a), LDL-C and apoB levels were measured in biological samples collected at baseline (2006 - 2010) among the ∼500,000 participants.

Lp(a) levels were measured in nmol/L, using immunoturbidimetric analysis on a Randox AU5800. In this project, we used the full range Lp(a) measurements (https://biobank.ndph.ox.ac.uk/showcase/dset.cgi?id=2321), rather than the constrained version which truncated the maximum reported range at 189 nmol/L and the minimum reported range at 3.8 nmol/L (https://biobank.ndph.ox.ac.uk/showcase/field.cgi?id=30790).

LDL-C levels were measured using the direct test in mmol/L by an enzymatic protective selection analysis on a Beckman Coulter AU5800 (https://biobank.ndph.ox.ac.uk/showcase/field.cgi?id=30780).

ApoB levels were measured in g/L, using immunoturbidimetric analysis on a Beckman Coulter AU5800 (https://biobank.ndph.ox.ac.uk/showcase/field.cgi?id=30640).

We removed sex mismatch, sex chromosome aneuploidy and heterozygosity outliers for quality control purposes. We restricted our analyses to the European (EUR) population with self-reported ethnicities being Caucasian/White. For each of the three lipid biomarkers, we split participants with available lab measurements into a 90% training dataset and a 10% testing dataset with random sampling, in order to construct the lipid biomarker GRS and evaluate the GRS prediction performance.

### 23andMe Research Participants

We selected approximately 7.3 million European research participants from the 23andMe customer base, who provided informed consent to participate in the research online, under a protocol approved by the external AAHRPP-accredited IRB, Ethical & Independent Review Services (E&I Review, now Salus IRB). Inclusion criteria of the research participants was based on their consent status at the time when the data analysis was initiated.

### 23andMe Genotyping and Imputation

Saliva samples of the 23andMe research participants were genotyped on one of five genotyping platforms. The genotype data obtained from each genotyping platform were then phased and imputed separately against an imputation panel that combines three independent reference panels: the HRC panel, the UK BioBank 200K Whole Exome Sequencing (WES) reference panels and the 23andMe reference panel, which was built by 23andMe using internal and external cohorts. Detailed descriptions of the 23andMe genotyping, imputation, and quality control procedures can be found at Reynoso, A. *et al*., 2024.

### 23andMe Disease Outcomes

We collected self-reported disease diagnosis history from the 23andMe research participants through web-based surveys, and used information derived from the surveys to define 489 binary / quantitative phenotypes for the phenome-wide MR analyses. In the web-based surveys, participants were asked to respond to the question “Have you ever been diagnosed with, or treated for, a specific disease type?”, with possible responses “Yes / No / I’m not sure”. We defined the binary phenotype with cases as those who responded “Yes” to the given diagnosis/treatment survey question, and controls as those who answered “No” to that question. Those 489 phenotypes covered a wide spectrum of disease areas, including autoimmune, cardiovascular, metabolic and cancer. We further defined a composite cardiovascular endpoint (which we denoted as major adverse cardiovascular event, or, MACE for short) on a set of expanded major cardiovascular adverse events, including coronary artery disease (CAD), heart attack, and stroke, where cases were those with at least one of the above listed conditions, controls had none of those conditions. The MACE endpoint was chosen to approximate the primary efficacy outcome of ongoing Lp(a) phase III cardiovascular outcome trials (ClinicalTrials.gov ID: NCT04023552).

### Lipid Biomarker Genetic Risk Scores

Genetic risk scores (GRS) aggregate an individual’s common genetic liability into a single value estimate. For each individual, GRS were constructed as a sum of imputed dosages on a set of SNPs, weighted by corresponding effect size estimates derived from the genetic association analyses in UKB for Lp(a), LDL-C and apoB, respectively.

To construct the Lp(a) GRS, we restricted our analyses to the *LPA* locus owing to the oligogenic genetic architecture of Lp(a). Specifically, we focused on the region within 1Mb of the transcription start site (TSS) of the *LPA* gene (GRCh38: chr6: 159664259-161664259). To increase the predictive accuracy of *LPA* GRS on Lp(a) measurements, we performed Bayesian fine-mapping with SuSiE (Wang, G. *et al*., 2020) on the locus. We used the top SNP with highest posterior inclusion probability (PIP) from each SuSiE credible set as the candidate genetic instruments, and selected 29 SNPs that retained a conditionally genome-wide significant association with Lp(a) to build the *LPA* GRS (details in Supplementary Notes, *LPA* GRS Instrument Selection for Univariable MR).

Unlike the concentration of Lp(a), which is predominately explained by a single locus, LDL-C and apoB levels are highly polygenic (Supplementary Figure 2, 3). Therefore, we ran genome-wide association studies (GWAS) followed by distance-based clumping, and selected SNPs reaching genome-wide significance (p-value <= 5×10^-8^) to construct GRS for LDL-C and apoB, which comprised of 169 and 175 SNPs, respectively (details in Supplementary Notes, LDL-C and apoB GRS Instrument Selection for Univariable MR).

### Mendelian Randomization and Sensitivity Analysis

We first conducted two-sample univariable Mendelian randomization analysis with individual level data, implemented with the two-stage least-squares regression (2SLS) approach. We ran a phenome-wide regression of 489 phenotypes on the *LPA* GRS in the 23andMe European cohort with a linear/logistic regression model, adjusting for age, sex, the top five within-europe principal components (PC) to account for residual population structure, and dummy variables for genotype platforms to account for genotype batch effects. We applied a Bonferroni correction threshold of 10^-4^ (≅ 0.05/489) to control for type I error inflation due to multiple testing. For each phenotype that reached a p-value ≤ 10^-4^ in the phenome-wide MR analysis with *LPA* GRS, we further tested their associations with the LDL-C and apoB GRS under the same linear/logistic model setting. To facilitate a like-for-like comparison on disease endpoints among Lp(a) vs LDL-C vs apoB, we calibrated their causal estimates to the same magnitude of effect for an endpoint conventionally used in randomized clinical trials for CVD, and then looked into whether the three lipid biomarkers had divergent effects across other diseases after calibration. Specifically, we first scaled the *LPA*, LDL-C and apoB GRS so that their univariable MR effect sizes on the MACE composite endpoint were the same. We then applied the MACE-derived scaling factors to *LPA*, LDL-C and apoB GRS respectively for univariable MR analyses on other diseases which had positive MR findings for Lp(a), anchoring the MR estimates to the same risk increase in MACE. We then performed likelihood ratio tests (LRT) to quantify whether genetically predicted Lp(a) imposed similar disease risk for non-MACE endpoints compared to LDL-C and apoB (details in Supplementary Notes, Likelihood Ratio Test).

Besides the primary 2SLS approach on individual-level data, we performed additional univariable two-sample MR analyses (2SMR) with the summary-level statistics to verify robustness of our agnostic MR findings. We implemented the 2SMR analysis in the “TwoSampleMR” package v0.56 in R v3.6.2, with the exposure summary statistics extracted from the UKB Lp(a), LDL-C, or apoB genetic association analysis, and the disease outcome estimates obtained from the 23andMe database (details in Supplementary Notes, Univariable Mendelian Randomization Sensitivity Analysis).

We also performed colocalization analysis to evaluate whether Lp(a) and disease outcomes shared causal variants in the *LPA* locus, using the “coloc” package v5.1.0 in R v3.6.2 (details in Supplementary Notes, Colocalization Analysis). It is possible that the genetic predictors of Lp(a) could be in LD with other variants that independently influence the disease outcomes, which might lead to violation of the exclusion restriction assumption (i.e. the instrument affects the outcome through only the exposure of interest) of MR inferences and thus leads to bias in the MR estimates. Therefore, having colocalization evidence to establish the presence of shared causal variants between the exposure (Lp[a]) and the disease outcome can increase the robustness of causal inference.

In order to assess the direct causal effects of Lp(a) on MACE, accounting for its cholesterol content (measured via the LDL-C assay) and apoB content (measured via the apoB assay), we conducted the two-sample multivariable MR analysis with individual level data, implemented using the 2SLS approach. Specifically, we regressed the binary MACE case/control status on the *LPA* and LDL-C GRS, as well as *LPA* and apoB GRS in a logistic regression model, with the same set of adjusting covariates as described in the univariable 2SLS setting (details in Supplementary Notes, GRS Instrument Selection for Multivariable MR). We did not perform multivariable MR analysis accounting for Lp(a), LDL-C and apoB simultaneously, due to the high degree of correlation between the LDL-C and apoB measurements observed in UKB (Supplementary Table 1). We used the conditional F statistics to assess the instrumental variable strengths and conditional Cochran’s Q statistics to assess horizontal pleiotropy. We obtained those statistics from the “MVMR” package v0.374 in R v3.6.2, with the exposure summary statistics extracted from the UKB Lp(a), LDL-C, or apoB genetic association analysis, and the disease outcome summary statistics extracted from the 23andMe GWASs.

## Results

### Cohort Characteristics

Among individuals in the UKB self-reporting as European/Caucasian, data were available for 379,520 participants with measurements of Lp(a), 387,768 with LDL-C, and 386,596 with apoB to construct GRS and evaluate the GRS prediction performance. The median plasma Lp(a) concentration was 18.69 nmol/L (Q1: 7.38 nmol/L, Q3: 73.65 nmol/L) in the 90% training cohort. Corresponding values for LDL-C were median 3.53 mmol/L (Q1: 2.96 mmol/L, Q3: 4.13 mmol/L) and for apoB median 1.02 g/L (Q1: 0.87 g/L, Q3: 1.19 g/L). The summary statistics for Lp(a), LDL-C and apoB were very similar in their 10% testing cohorts compared with those in their corresponding training cohorts (Table 1; Figure 1-3: Panel A, B).

**Figure 1.**
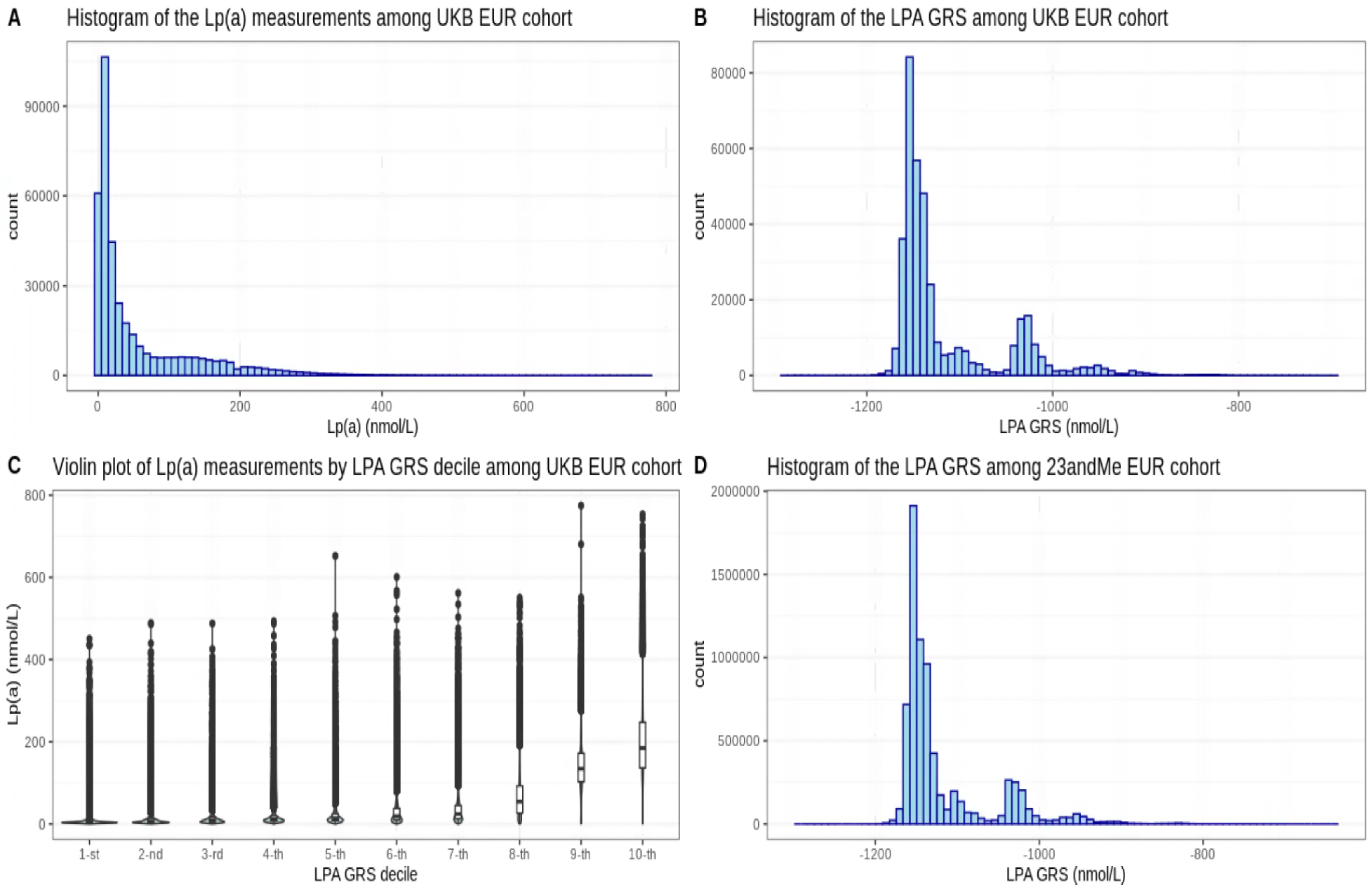
Distributions of measured lipoprotein(a) and *LPA* genetic risk scores (UKB EUR v.s 23andMe EUR). (A) Histogram of measured Lp(a) levels in UKB full EUR cohort; (B) Histogram of *LPA* genetic risk score (GRS) in UKB full EUR cohort; (C) Violin plot of measured Lp(a) levels by *LPA* GRS decile in UKB full EUR cohort; (D) Histogram of *LPA* GRS in 23andMe EUR cohort.

**Figure 2.**
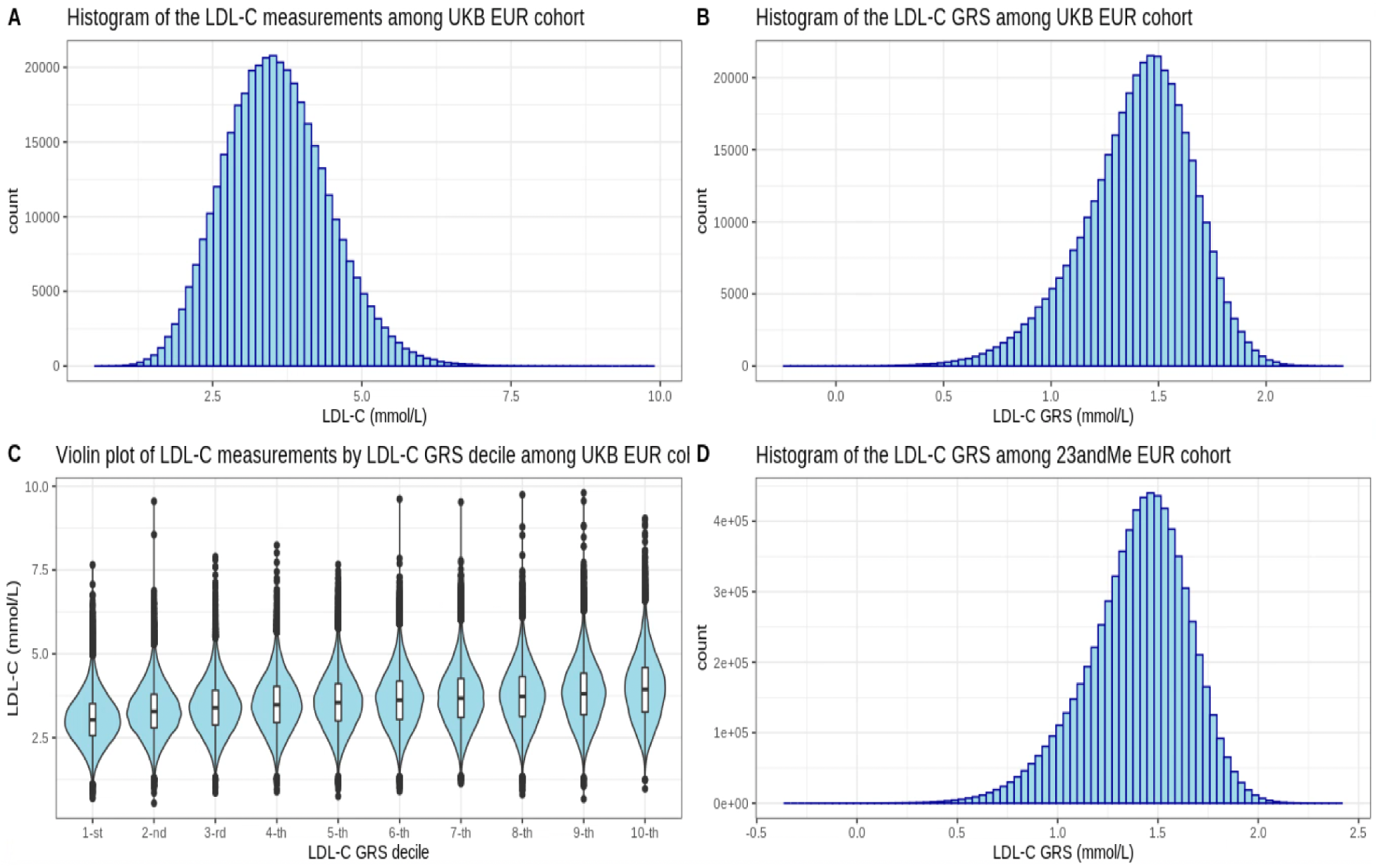
Distributions of measured LDL-C and LDL-C genetic risk scores (UKB EUR v.s 23andMe EUR). (A) Histogram of measured LDL-C levels in UKB full EUR cohort; (B) Histogram of LDL-C genetic risk score (GRS) in UKB full EUR cohort; (C) Violin plot of measured LDL-C levels by LDL-C GRS decile in UKB full EUR cohort; (D) Histogram of LDL-C GRS in 23andMe EUR cohort.

**Figure 3.**
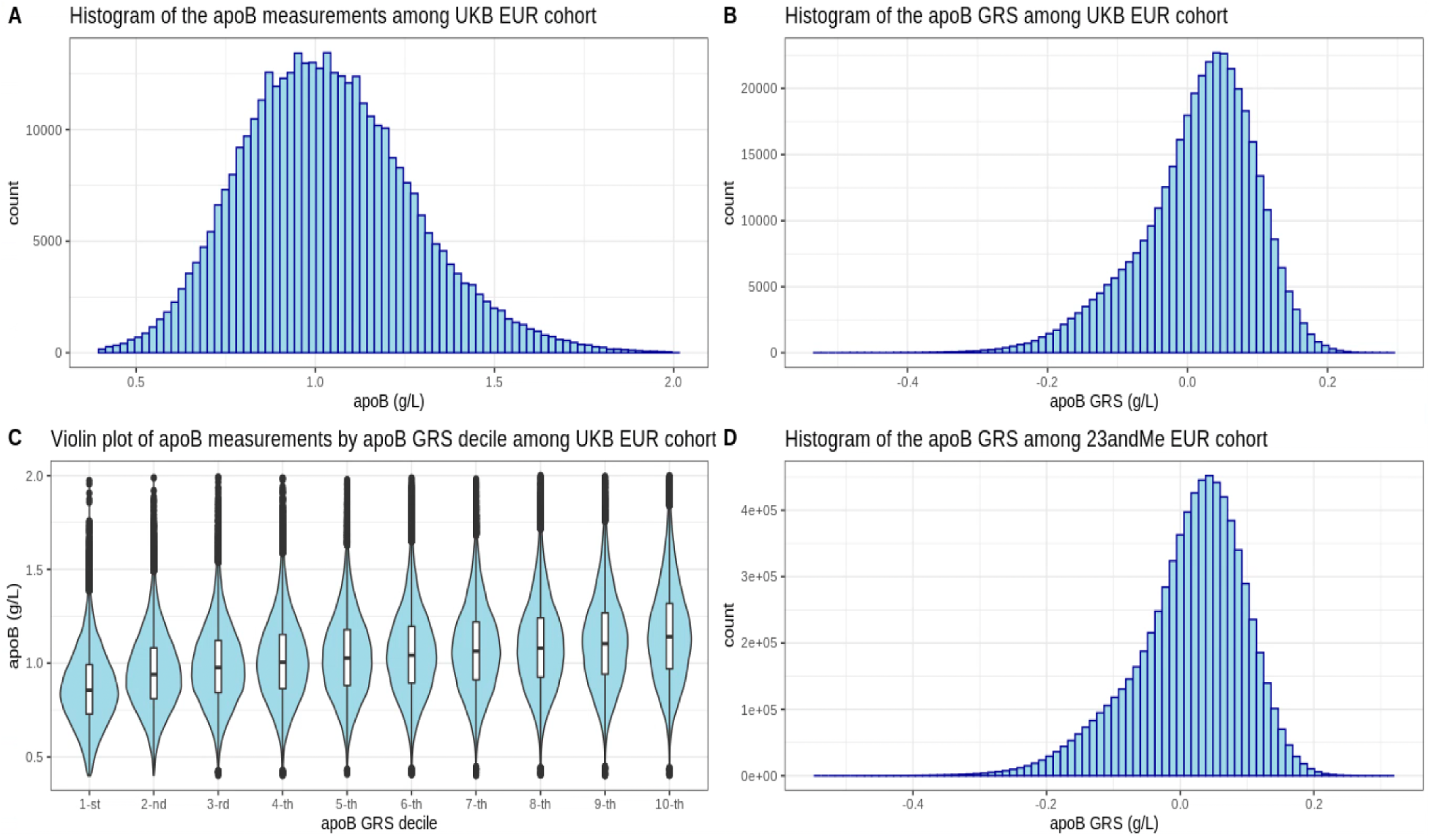
Distributions of measured apoB and apoB genetic risk scores (UKB EUR v.s 23andMe EUR). (A) Histogram of measured apoB levels in UKB full EUR cohort; (B) Histogram of apoB genetic risk score (GRS) in UKB full EUR cohort; (C) Violin plot of measured apoB levels by apoB GRS decile in UKB full EUR cohort; (D) Histogram of apoB GRS in 23andMe EUR cohort.

**Table 1:**
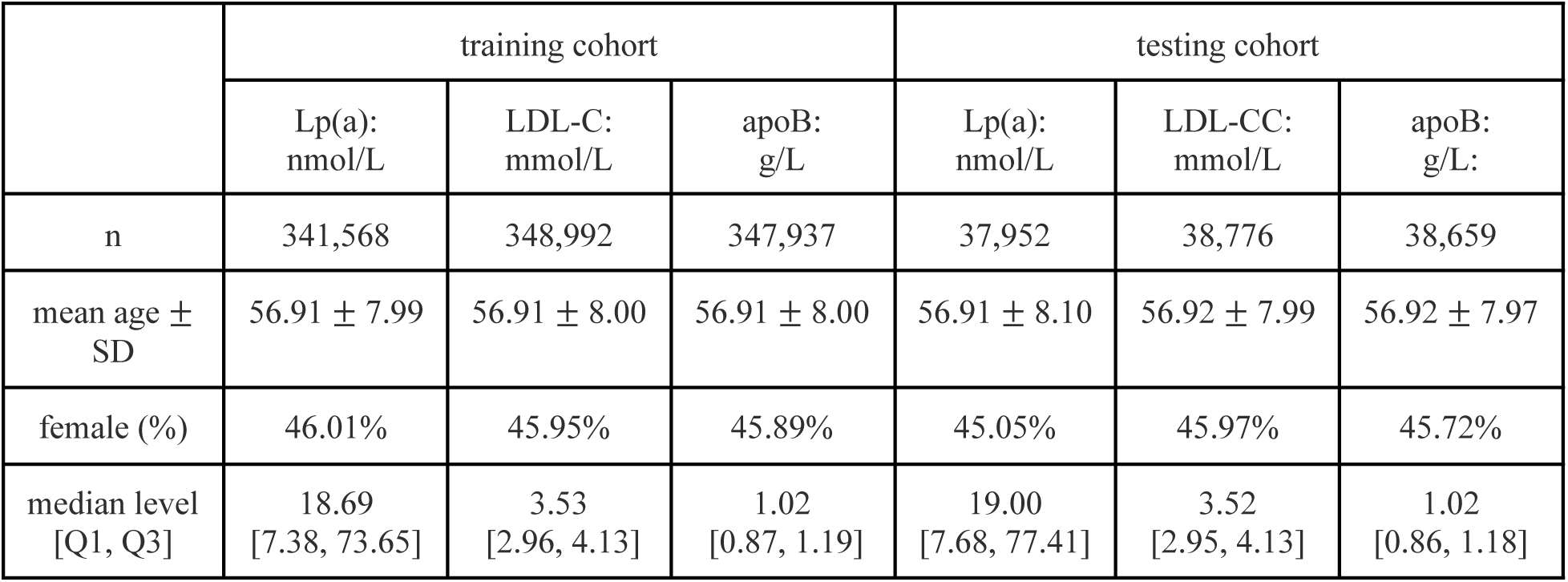
Baseline characteristics of UK Biobank European study population. Abbreviations: Lp(a), lipoprotein(a); LDL-C, low density lipoprotein cholesterol; apoB: apolipoprotein B; Q1, first quartile; Q3, third quartile.

|  | training cohort |  |  | testing cohort |  |  |
| --- | --- | --- | --- | --- | --- | --- |
|  | Lp(a):<br>nmol/L | LDL-C:<br>mmol/L | apoB:<br>g/L | Lp(a):<br>nmol/L | LDL-CC:<br>mmol/L | apoB:<br>g/L: |
| n | 341,568 | 348,992 | 347,937 | 37,952 | 38,776 | 38,659 |
| mean age ± SD | 56.91 ± 7.99 | 56.91 ± 8.00 | 56.91 ± 8.00 | 56.91 ± 8.10 | 56.92 ± 7.99 | 56.92 ± 7.97 |
| female (%) | 46.01% | 45.95% | 45.89% | 45.05% | 45.97% | 45.72% |
| median level<br>[Q1, Q3] | 18.69<br>[7.38, 73.65] | 3.53<br>[2.96, 4.13] | 1.02<br>[0.87, 1.19] | 19.00<br>[7.68, 77.41] | 3.52<br>[2.95, 4.13] | 1.02<br>[0.86, 1.18] |

A total of ∼7.3 million participants of European genetic ancestry from the 23andMe research cohort with at least one phenotypic value were included in the phenome-wide MR analyses. A list of disease-specific sample size counts with positive Lp(a) MR findings can be found in Supplementary Table 5.

### Variance Explained by the Lipid Biomarker GRS

Using 341,568 training samples from the UKB (i.e. a 90% training cohort), we selected 29 genetic variants identified by the SuSiE fine-mapping algorithm around the *LPA* locus to construct the genetic risk score for Lp(a) (Supplementary Table 2). The *LPA* GRS comprising those 29 variants explained 68.18% of the phenotypic variance of Lp(a) concentration among 37,952 UKB hold out samples (i.e. a 10% testing cohort), where the phenotypic variance was measured as squared pearson correlation coefficient between the Lp(a) levels and *LPA* GRS. We note that among the 29 Lp(a) genetic instruments, the two most significant SNPs, namely rs10455872 and rs140570886, accounted for 63.99% of the total variance explained (Supplementary Table 2).

We constructed LDL-C GRS with 169 genome-wide significant SNPs selected from the UKB LDL-C GWAS based on 348,992 training samples (Supplementary Table 3). The LDL-C GRS explained 9.18% of the variance in LDL-C concentration among the 38,776 UKB hold out samples. Similarly, the apoB GRS was constructed using 175 genome-wide significant SNPs (n=347,937 training samples) with the GRS explaining 11.16% of the apoB variance (n=38,659 testing samples; Supplementary Table 4).

We note that the LDL-C and apoB measurements showed a high degree of correlation, both phenotypically and genetically. Specifically, the two lipid measurements had a Pearson correlation coefficient equal to 0.958, and their GWAS summary statistics had a cross-trait LD Score (LDSC) regression estimate equal to 0.946. Consequently, the LDL-C and apoB GRS shared 98 SNPs in common, with the Pearson correlation coefficient equal to 0.915 in the UKB cohort (Supplementary Table 1).

### Phenome-wide Univariable Mendelian Randomization Analysis

After evaluating the performance of those lipid biomarker GRS in the UK Biobank cohort, we built the *LPA*, LDL-C and apoB GRS in approximately 7.3M samples from the 23andMe cohort in order to proceed with the two sample 2SLS MR analyses (Figure 1, 2, 3: Panel D). With genetically-instrumented Lp(a) across 489 binary/quantitative traits in the 23andMe European cohort, the univariable MR-PheWAS yielded 25 genetically-predicted causal effects of Lp(a) that surpassed multiple testing (Bonferroni correction p-value ≤ 10^-4^). Those 25 signals confirmed previously reported Lp(a) associations and revealed additional associations that were all largely confined to diseases of the cardiovascular system (Figure 4, 5).

**Figure 4.**
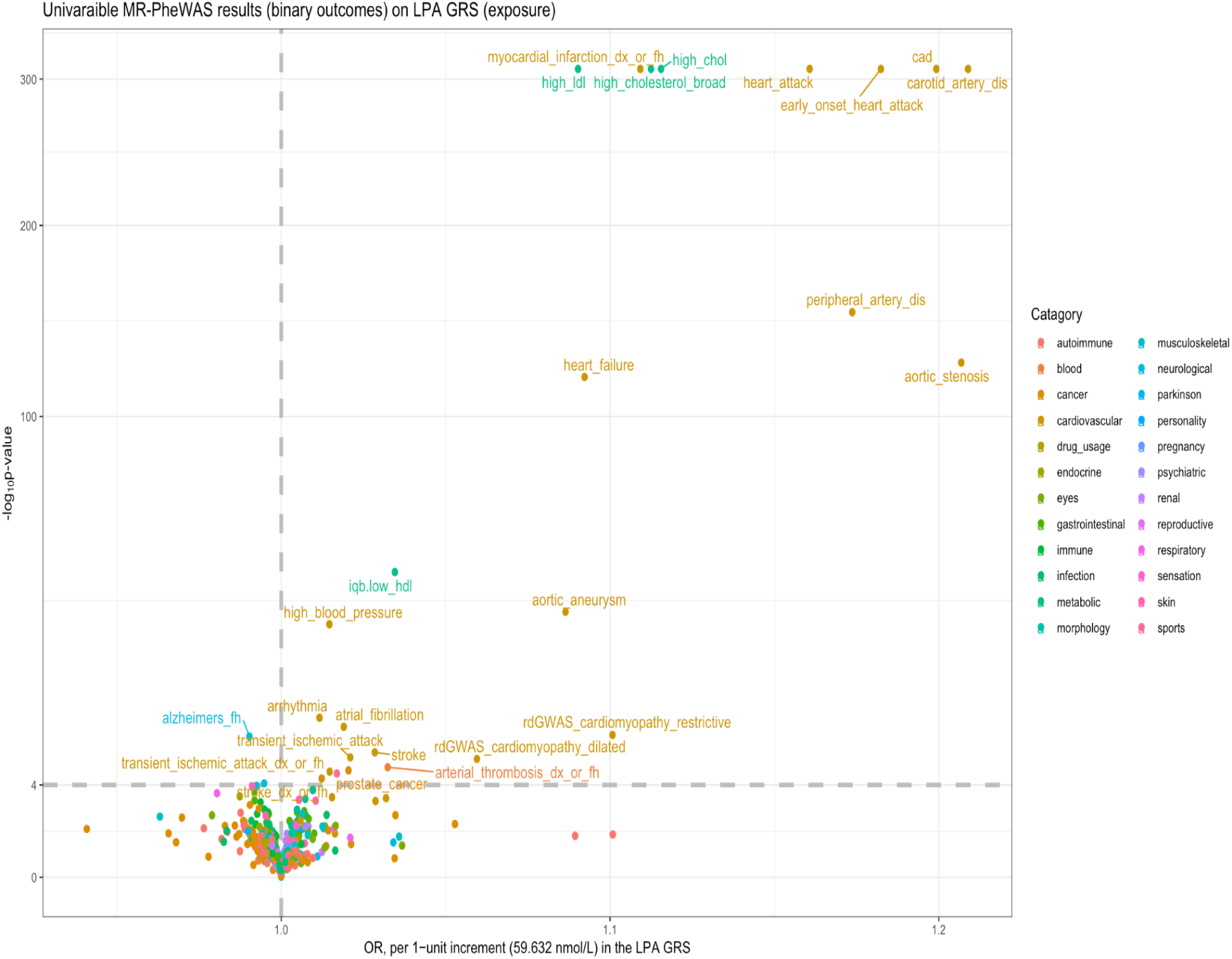
Scatter plots of the 2SLS univariable MR-PheWAS results of *LPA* GRS (exposure) on binary phenotypes (outcome). Summary of MR-PheWAS results among 442 binary outcomes (each represented by a dot, colored according to their disease categories). We note that none of the 47 quantitative phenotypes reached the significant threshold at p-value ≤ 10^-4^ in MR-PheWAS. The x-axis denotes the OR and y-axis denotes the -log10(p-value). Dots above the dashed horizontal line, drawn at -log10(p-value) = 4, are those 25 agnostic signals that surpassed multiple testing, with their phenotypic names displayed. Phenotypic definitions for each of those 25 signals can be found in Supplementary Table 5. The dashed vertical line at OR = 1 represents the null effect for binary outcomes.

Using the *LPA* GRS, we identified genetically-predicted causal effects of Lp(a) on atherosclerotic vascular disease including CAD, myocardial infarction, stroke, transient ischemic attack (TIA) and carotid artery disease. Additional vascular disease effects were evident for peripheral arterial disease, aortic aneurysm and arterial thrombosis. We also found effects on structural and functional heart disease including aortic stenosis, heart failure, cardiomyopathies (both restrictive and dilated) as well as atrial fibrillation. We also identified signals for established CVD risk factors, including high blood pressure, high cholesterol, high LDL, and low HDL (Supplementary Table 5).

Beyond cardiovascular diseases and their risk factors, MR-PheWAS also suggested provisional causal relationships between elevated Lp(a) instrumented through the *LPA* GRS and prostate cancer risk (OR = 1.020, 95% CI = [1.012, 1.029], p-value = 4.2×10^-6^), and a lower risk of family history of Alzheimer’s disease (OR = 0.990, 95% CI = [0.987, 0.993], p-value = 4.7×10^-10^) (Supplementary Table 5).

To facilitate a comparison of effect sizes, we repeated the univariable MR analyses for the 25 endpoints using genetic instruments for LDL-C and apoB (Figure 5; Supplementary Table 5). We scaled the LDL-C and apoB GRS such that their univariable MR effect sizes on the MACE composite endpoint were both equal to that of the *LPA* GRS (i.e. OR = 1.10 on MACE for the three biomarkers). This provided a scaling rubric for each GRS, which was equivalent to 59.632 nmol/L higher Lp(a), 0.298 mmol/L higher LDL-C, and 0.091 g/L higher apoB. Among the 18 cardiovascular related diseases, genetically-instrumented Lp(a) had similar effect sizes compared to genetically-instrumented LDL-C and apoB, with the exception of peripheral artery disease, aortic stenosis and dilated cardiomyopathy, where Lp(a) had significantly larger effect sizes than LDL-C and apoB (with LRT p-values < 10^-3^; Supplementary Table 6).

**Figure 5.**
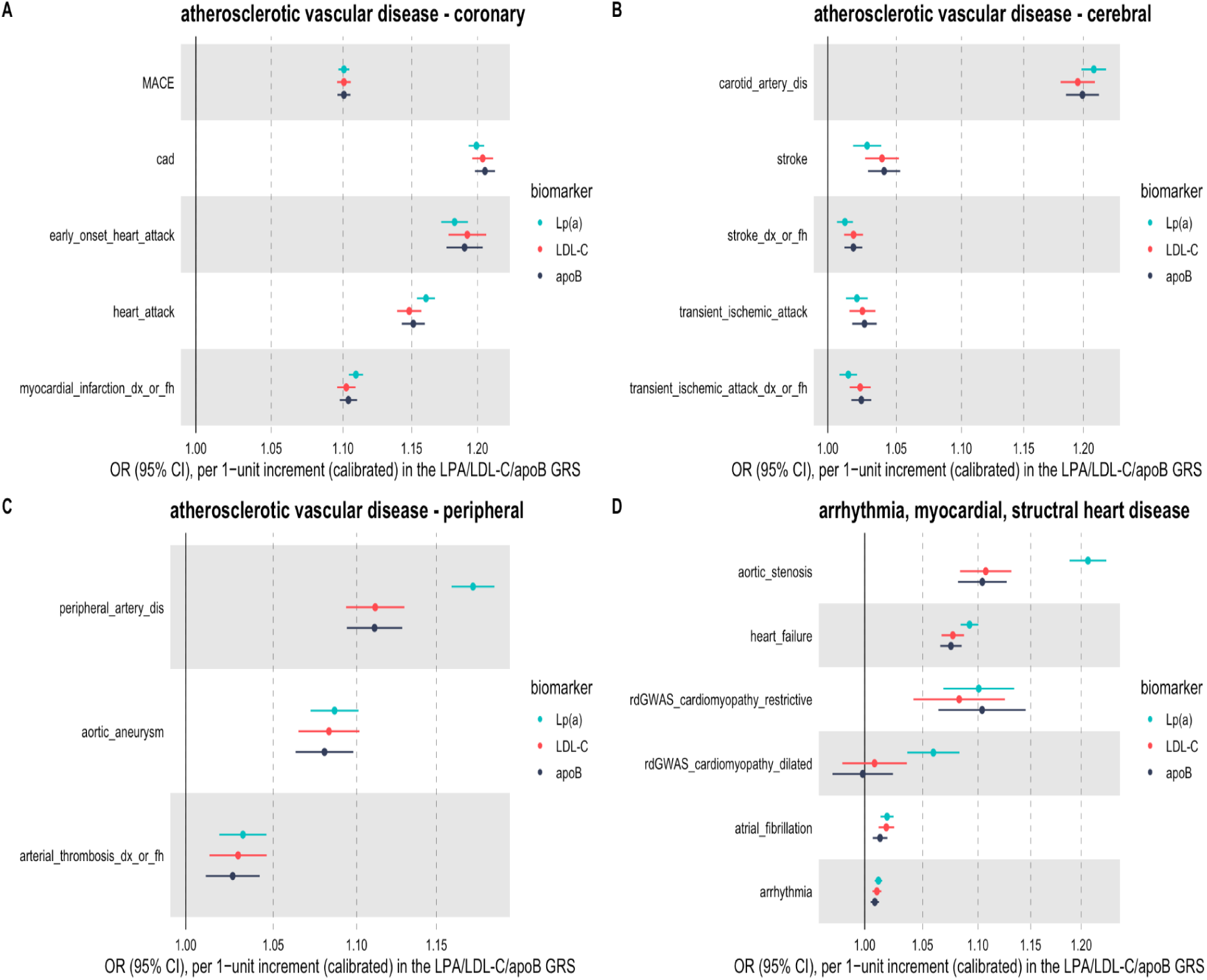
Forest plot of the univariable 2SLS MR results of genetically-instrumented Lp(a), LDL-C and apoB onto MACE and cardiovascular disease outcomes. The univariable MR OR and 95% CI for each lipid biomarker were based on their corresponding per 1-unit increment, with every 59.632 nmol/L increase in the *LPA* GRS, every 0.298 mmol/L increase in the LDL-C GRS, and every 0.091 g/L increase in the apoB GRS. We divided the cardiovascular disease outcomes into multiple categories: (A) atherosclerotic vascular disease - coronary; (B) atherosclerotic vascular disease - cerebral; (C) atherosclerotic vascular disease - peripheral; (D) arrhythmia, myocardial, structural heart disease. Phenotypic definitions for each of those cardiovascular disease outcomes can be found in Supplementary Table 5.

Colocalization analysis suggested that of those 25 diseases/traits identified as potentially being the causal consequence of genetically-instrumented elevated Lp(a), 22 had strong evidence (posterior probability [PP] >0.8) of sharing the same underlying causal variant with Lp(a) in the region (Supplementary Table 7; Supplementary Figure 9.1 - 9.25), which provided additional evidence in supporting the MR assumptions. In summary, among the 18 cardiovascular related diseases, colocalization signals were observed on most of the Lp(a) vs disease pairs, except for transient ischemic attack and dilated cardiomyopathy. We also observed colocalization analysis among all five risk factors for CVD, and the family history of Alzheimir’s disease. Colocalization between Lp(a) and prostate cancer suggested that they have different causal variants in the region.

Additional univariable 2SMR approaches (IVW, MR-Egger and weighted median) that are more robust to potential violation of the exclusion restriction criteria yielded similar MR estimates to the corresponding 2SLS MR estimates (Supplementary Table 8, 9, 10). Intercepts from MR-Egger suggested no evidence of directional horizontal pleiotropy for the Lp(a) MR inference among all 25 phenotypes (p-value > 0.05; Supplementary Table 11), and no evidence of directional horizontal pleiotropy for most of the LDL-C and apoB MR inference, with the exception of dilated cardiomyopathy, high cholesterol, high LDL and high blood pressure (Supplementary Table 12, 13). Cochran’s Q statistics suggested evidence of effect size heterogeneity (Supplementary Table 14, 15, 16). Thus we applied heterogeneity and Steiger filtering to each lipid biomarkers’ genetic instruments (Hemani G. *et al*., 2017). The 2SMR analysis on the remaining set of SNPs yielded directionally consistent effects compared to using the full set of SNPs (Supplementary Table 17, 18, 19; Supplementary Figures 10.1.1 - 10.25.2, 11.1.1 - 11.25.2, 12.1.1 - 12.25.2). We note that Lp(a) no longer had a significant MR signal with prostate cancer following SNP filtering (Supplementary Table 17; Supplementary Figure 10.24.2).

### Multivariable Mendelian Randomization Analysis

In order to establish the direct effect of Lp(a) on the composite outcome MACE, we performed multivariable MR accounting for cholesterol (measured via the LDL-C direct assay) and apoB (measured via the apoB assay), with the genetic instruments for the three lipid entities scaled using the same rubric as in the univariable MR analysis. When including genetic instruments for LDL-C or apoB, multivariable MR estimates for Lp(a) were very similar to estimates derived from univariable MR, suggesting that the Lp(a) effects on risk of MACE are largely independent of LDL-C or apoB (Supplementary Table 20). For comparison, estimates from both the univariable and multivariable MR models are depicted in Figure 6.

**Figure 6.**
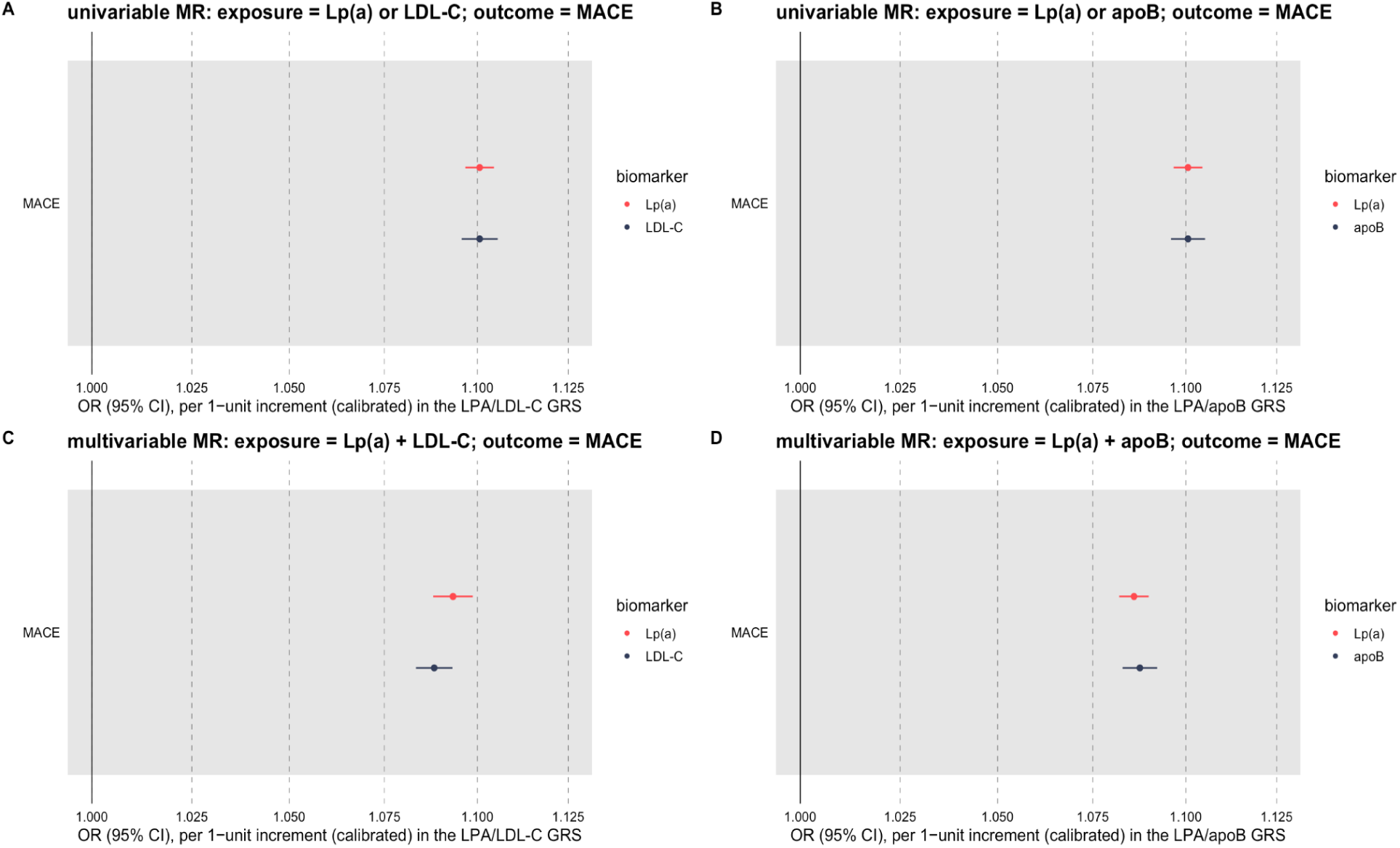
Forest plots to compare the univariable vs multivariable 2SLS MR estimates for MACE. The univariable and multivariable MR OR and 95% CI for each lipid biomarker were based on their corresponding per 1-unit increment, with every 59.632 nmol/L increase in the *LPA* GRS, every 0.298 mmol/L increase in the LDL-C GRS, and every 0.091 g/L increase in the apoB GRS. For comparison, we presented in (A) the univariable 2SLS MR estimates of genetically-instrumented Lp(a) or LDL-C onto MACE; (B) the univariable 2SLS MR estimates of genetically-instrumented Lp(a) or apoB onto MACE; (C) the multivariable 2SLS MR estimates of genetically-instrumented Lp(a) and LDL-C onto MACE; and (D) the multivariable 2SLS MR estimates of genetically-instrumented Lp(a) and apoB onto MACE.

## Discussion

In this study, by combining a powerful genetic instrument for Lp(a) from UK Biobank, together with extensive health outcome survey responses at unprecedented sample size from 23andMe, we were able to characterize the genetically-predicted causal architecture of plasma Lp(a) across a broad spectrum of diseases/traits. Agnostic univariable MR-PheWAS analysis not only confirmed previously reported genetically-predicted Lp(a) effects on coronary artery disease and aortic stenosis, but also revealed novel effects largely confined to cardiovascular endpoints. Furthermore, multivariable MR analyses showed that Lp(a) levels are genetically predicted to increase the risk of cardiovascular events, even when accounting for LDL-C or apoB.

These findings could have clinical implications in five ways. First, our univariable MR findings provide support that drugs targeting Lp(a) will impact a broad range of cardiovascular diseases beyond CAD, with novel evidence of Lp(a) having a predicted causal effect on both restrictive and dilated cardiomyopathy. Consequently, by including endpoints that are the causal consequence of Lp(a), clinical trials that include multiple measures of cardiovascular disease in addition to CAD as part of a composite endpoint are likely to yield a positive clinical trial outcome. Second, the causal consequences of genetically-predicted Lp(a) are mostly constrained to cardiovascular traits meaning that Lp(a) lowering therapies are unlikely to have target-mediated unintended adverse effects. Third, our effect size comparison of Lp(a) vs LDL-C vs apoB can serve as a reference for the clinical trial dosage of Lp(a) lowering treatments. Our findings suggest that lowering of Lp(a) by 90 mg/dL or 200 nmol/L are needed to achieve a 25% relative risk reduction (see next paragraph for more details); notably currently ongoing phase III RCTs are recruiting individuals with elevated Lp(a) values close to or more than 90 mg/dL, coupled with treatments that lower Lp(a) by up to 95% (PR Newswire, 2023). Fourth, our multivariable MR results indicate that patients with high plasma Lp(a) concentrations are likely to derive cardiovascular benefit from therapy with pharmacological lowering of Lp(a), even if they have well-controlled plasma LDL-C or apoB levels. Fifth, our comparison of Lp(a) to LDL-C and apoB scaled to the same effect on MACE identified Lp(a) to have a larger magnitude of effect on aortic valve stenosis, peripheral arterial disease and dilated cardiomyopathy. The differential effects of Lp(a) on these diseases may be explained by characteristics unique to Lp(a). For example, Lp(a) induces calcification of the aortic valve thereby promoting progression of aortic stenosis (Tsimikas, S., 2019). The peripheral arterial disease effects of Lp(a) may be driven by the apolipoprotein(a) glycoprotein which is not found in LDL-C: apolipoprotein(a) has prothrombotic properties due to its homology with plasminogen, thus interfering with fibrinolysis, and exhibits pro-inflammatory effects owing to oxidized phospholipids bound to apolipoprotein(a) (Boffa, M.B. and Koschinsky, M.L., 2016). The latter properties may also account for the excess risk of Lp(a) with dilated cardiomyopathy, where inflammation of the myocardium can be found on autopsy (Sikking, M.A. *et al*., 2023). The clinical implications are that, scaled to a same reduction in CVD as that derived from LDL-C or apoB lowering, Lp(a) lowering ought to yield larger comparative risk reductions in these individual vascular diseases.

Previous MR studies have shown that magnitudes of effect derived from Mendelian randomization in relation to blood lipids and vascular disease is several-fold larger than that seen in interventional trials of drugs targeting the same lipid trait (Holmes, M.V. and Smith, G.D., 2017; Ference, B.A. *et al*., 2017). This is principally considered to be due to the cumulative effect of lipids on the atherosclerotic disease process, and that genetic instruments for e.g. LDL-C yield lifelong disease associations versus a comparable effect size derived from a randomized controlled trial of shorter duration. A study by Burgess and colleagues (Burgess, S. *et al*., 2018) found that, when both lipid measures were quantified in mg/dL, it took 2.63 fold more Lp(a) than LDL-C to achieve the same CHD risk reduction. In our study, a similar calculation derived a ratio of 2.37 (details in Supplementary Notes, Deriving an Estimate of Lp(a) Lowering Required to Achieve a CVD Risk Reduction Equivalent to that of LDL-C). Extrapolating this to achieve a 25% relative risk reduction in major vascular events, as seen with a 1 mmol/L reduction in LDL-C from statin trials (Collins, R. *et al*., 2016)., we anticipate that Lp(a) would be required to be lowered by about 90 mg/dL or 200 nmol/L, highlighting the importance of not only using Lp(a) lowering therapies that have a profound impact on Lp(a) lowering, but also selecting individuals that have sufficiently high Lp(a) concentrations at baseline. Given that, in contrast to other circulating blood lipid traits such as LDL-C or apoB, genetic variation plays such a critical role in regulating Lp(a) levels, use of a polygenic risk score might have clinical utility in identifying individuals with elevated Lp(a) who may be suitable for receiving Lp(a) lowering therapies, once such drugs receive marketing authorizations.

There are several limitations to our study. First, our phenotypic endpoints relied on self-reported disease status and lacked outcome adjudication. Bias in self-reporting collected from survey questionnaires may have affected certain results; for example, fatal CVD events wouldn’t be captured in our composite MACE endpoint derived based on the self-reported CVD events, which may impact the magnitude of MR effect estimates. The observed inverse associations with a family history of Alzheimer’s disease (rather than the participants themselves getting the diagnosis) with elevated Lp(a) levels merits further investigation. Past studies on the relationship between Lp(a) and Alzheimer disease risk were inconclusive. For example, Röhr, F. *et al*. (2020) suggested sex-specific low plasma concentrations of Lp(a) were associated with better cognitive performance in males only, although the signal was rather weak at merely a nominal significance level and was inconclusive after correcting for multiple testing. In contrast, Larsson, S.C. *et al*. (2020) revealed a weak inverse association of genetically predicted Lp(a) levels with self-reported parental history of Alzheimer disease or dementia. When conducting the MR-PheWAS analysis, we sought to curate a representative list of the phenome from the available survey response data, in order to derive a comprehensive understanding of the downstream consequences of elevated Lp(a) on human health. Although the majority of our PheWAS phenotypes are binary (including clinical events and dichotomized quantitative traits such as blood lipids), our analyses can serve as a foundation for establishing the full spectrum of phenotypic characterization of genetically elevated Lp(a) levels. Second, owing to the right skewed distribution of serum Lp(a) concentration, several studies have applied log-transformation to the Lp(a) levels and examined the corresponding association with CVD risks (Bennet, A. *et al*., 2008; Emerging Risk Factors Collaboration, 2009; Nordestgaard, B.G., *et al*., 2010; Emdin, C.A. *et al*., 2016). However, Burgess, S. *et al*. (2018) proposed that log-transformation of Lp(a) may not be suitable for assessing the potential clinical benefit of Lp(a) lowering, since the same proportional changes represented on a log scale can result in substantially different absolute changes, depending on the baseline Lp(a) levels. Furthermore, they showed that fixed changes in absolute Lp(a) concentrations led to equal odds ratios (ORs) for CHD regardless of the starting Lp(a) concentration, as displayed in their Figure 2A. In addition, we observed that log transformation led to a left skewed and bi-modal distribution (Supplementary Figure 6), and the genetic association patterns on the log transformed Lp(a) were very similar to those on the raw Lp(a) measurements (Supplementary Figure 7.1, 7.2). Consequently, we anticipate that the MR-PheWAS analysis would yield a near-identical set of phenotypic associations if we were to log-transform Lp(a), since the most significant genetic instrument remained the same (namely, rs10455872) and explained most of the Lp(a) variation on both the raw and log-transformed scales. Last, our study primarily focused on the population of European genetic ancestry, additional studies in the non-European populations are needed in order to assess the generalizability of our study conclusions into other ancestry groups. Satterfield, B.A. *et al*. (2021) reported that increase in Lp(a) level had non-significant associations with coronary heart disease and cerebrovascular disease in the African ancestry. From the powerful 23andMe database, we observed that Latino and African American populations had strong genetic association signals for rs10455872 (i.e. the strongest genetic instrument for Lp[a] which accounts for majority of the variance explained) in CAD, heart attack and myocardial infarction GWASs. Therefore, we speculate that the associations of high plasma Lp(a) concentration with cardiovascular diseases may pertain in those non-European populations (Lee, M.P. *et al*., 2023). Notably, the distribution of Lp(a) concentrations has pronounced ancestral differences, with Lp(a) levels in populations of African ancestry being up to 4 times higher than those in populations of European ancestry (Kraft, H.G. *et al*., 1996; Marcovina, S.M. *et al*., 1996; Li, J. *et al*., 2015). Such marked differences in Lp(a) distribution may be attributed to heterogeneity in Lp(a) genetic architecture, with recent studies showing SNPs strongly associated with Lp(a) among African Americans are monomorphic in non-Africans, speaking to the need for ancestry-specific genetic instruments. Expanding ancestral diversity to include African populations in future studies will be critical to elucidate the ancestry-specific causal effects of serum Lp(a), and ensure equitable use of therapeutics targeting Lp(a).

In summary, our study finds that Lp(a) is predicted to causally impact a broad range of cardiovascular endpoints including cardiomyopathies. Provided a suitable reduction in Lp(a), novel Lp(a) lowering drugs currently under development are predicted to have a substantial impact on addressing the unmet clinical need of treating and preventing residual CVD among individuals with elevated Lp(a). Once these drugs receive marketing authorizations, GRS for Lp(a) may provide a suitable approach to identify individuals who would benefit from Lp(a) testing and treatment.

## Data Availability

All data produced in the present study are available upon reasonable request to the authors.

## Acknowledgments

This research has been conducted using the UK Biobank resource under application number 95801. We thank 23andMe customers who consented to participate in research for enabling this study. We also thank employees of 23andMe who contributed to the development of the infrastructure that made this research possible.

The following members of the 23andMe Research Team contributed to this study: Stella Aslibekyan, Adam Auton, Elizabeth Babalola, Robert K. Bell, Jessica Bielenberg, Ninad S. Chaudhary, Zayn Cochinwala, Sayantan Das, Emily DelloRusso, Payam Dibaeinia, Sarah L. Elson, Nicholas Eriksson, Chris Eijsbouts, Teresa Filshtein, Pierre Fontanillas, Davide Foletti, Will Freyman, Zach Fuller, Julie M. Granka, Chris German, Éadaoin Harney, Alejandro Hernandez, Barry Hicks, David A. Hinds, M. Reza Jabalameli, Ethan M. Jewett, Yunxuan Jiang, Sotiris Karagounis, Lucy Kaufmann, Matt Kmiecik, Katelyn Kukar, Alan Kwong, Keng-Han Lin, Yanyu Liang, Bianca A. Llamas, Aly Khan, Steven J. Micheletti, Matthew H. McIntyre, Meghan E. Moreno, Priyanka Nandakumar, Dominique T. Nguyen, Jared O’Connell, Steve Pitts, G. David Poznik, Alexandra Reynoso, Shubham Saini, Morgan Schumacher, Leah Selcer, Anjali J. Shastri, Suyash Shringarpure, Keaton Stagaman, Teague Sterling, Qiaojuan Jane Su, Joyce Y. Tung, Susana A. Tat, Vinh Tran, Xin Wang, Wei Wang, Catherine H. Weldon, Peter Wilton.

## Supplementary Materials

Supplementary Notes

Supplementary Figures 1-7: UK Biobank lipid biomarkers GWAS plots

Supplementary Figure 8: GRS Instrument Selection Scheme for Multivariable MR

Supplementary Figures 9.1- 9.25: Lp(a) vs disease colocalization scatter plots

Supplementary Figures 10.1.1 - 10.25.2: MR scatter plots for Lp(a)

Supplementary Figures 11.1.1 - 11.25.2: MR scatter plots for LDL-C

Supplementary Figures 12.1.1 - 12.25.2: MR scatter plots for apoB

Supplementary Table 1: Correlation between Lp(a), LDL-C and apoB

Supplementary Table 2: Lp(a) genetic instruments for univariable MR

Supplementary Table 3: LDL-C genetic instruments for univariable MR

Supplementary Table 4: apoB genetic instruments for univariable MR

Supplementary Table 5: univariable 2SLS MR estimates (Lp[a], LDL-C, apoB)

Supplementary Table 6: pairwise LRT for the univariable 2SLS MR estimates

Supplementary Table 7: Lp(a) vs disease colocalization posterior probabilities

Supplementary Table 8: univariable 2SMR Lp(a) MR estimates

Supplementary Table 9: univariable 2SMR LDL-C MR estimates

Supplementary Table 10: univariable 2SMR sensitivity analysis for apoB, robust MR estimates

Supplementary Table 11: univariable 2SMR MR-Egger intercepts for Lp(a)

Supplementary Table 12: univariable 2SMR MR-Egger intercepts for LDL-C

Supplementary Table 13: univariable 2SMR MR-Egger intercepts for apoB

Supplementary Table 14: univariable 2SMR Cochran’s Q statistics for Lp(a)

Supplementary Table 15: univariable 2SMR Cochran’s Q statistics for LDL-C

Supplementary Table 16: univariable 2SMR Cochran’s Q statistics for apoB

Supplementary Table 17: univariable 2SMR Lp(a) MR estimates, after SNP filtering

Supplementary Table 18: univariable 2SMR LDL-C MR estimates, after SNP filtering

Supplementary Table 19: univariable 2SMR apoB MR estimates, after SNP filtering

Supplementary Table 20: multivariable 2SLS MR estimates

