## Supplementary Figures for "Evaluating genetically-predicted causal effects of lipoprotein(a) in human diseases: a phenome-wide Mendelian randomization study": Supplementary_Figure_1_to_7.docx

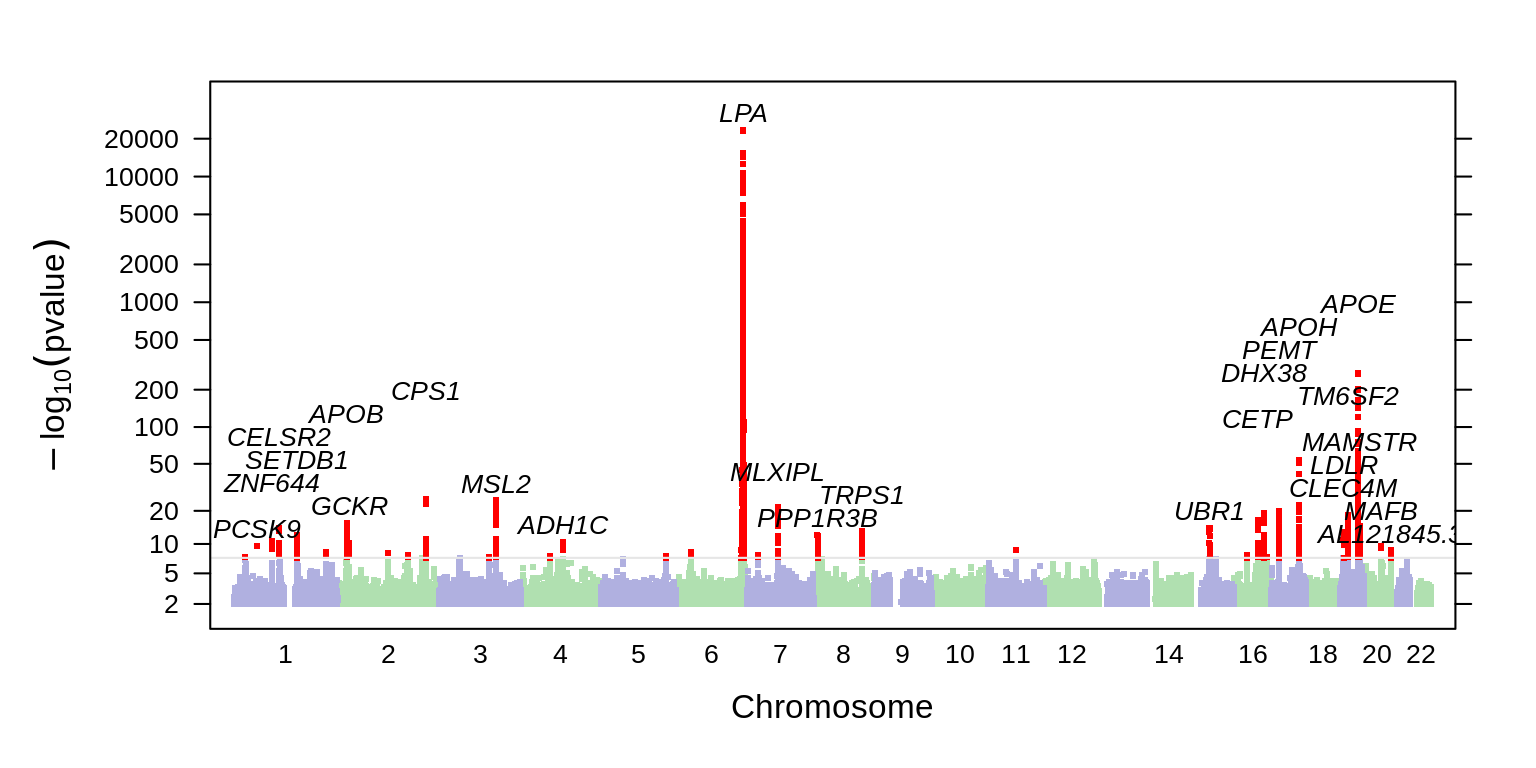

Supplementary Figure 1.1: Manhattan plot showing results from the Lp(a) GWAS in UKB training cohort. Top-associated variants at each locus are labeled with their nearest gene context.

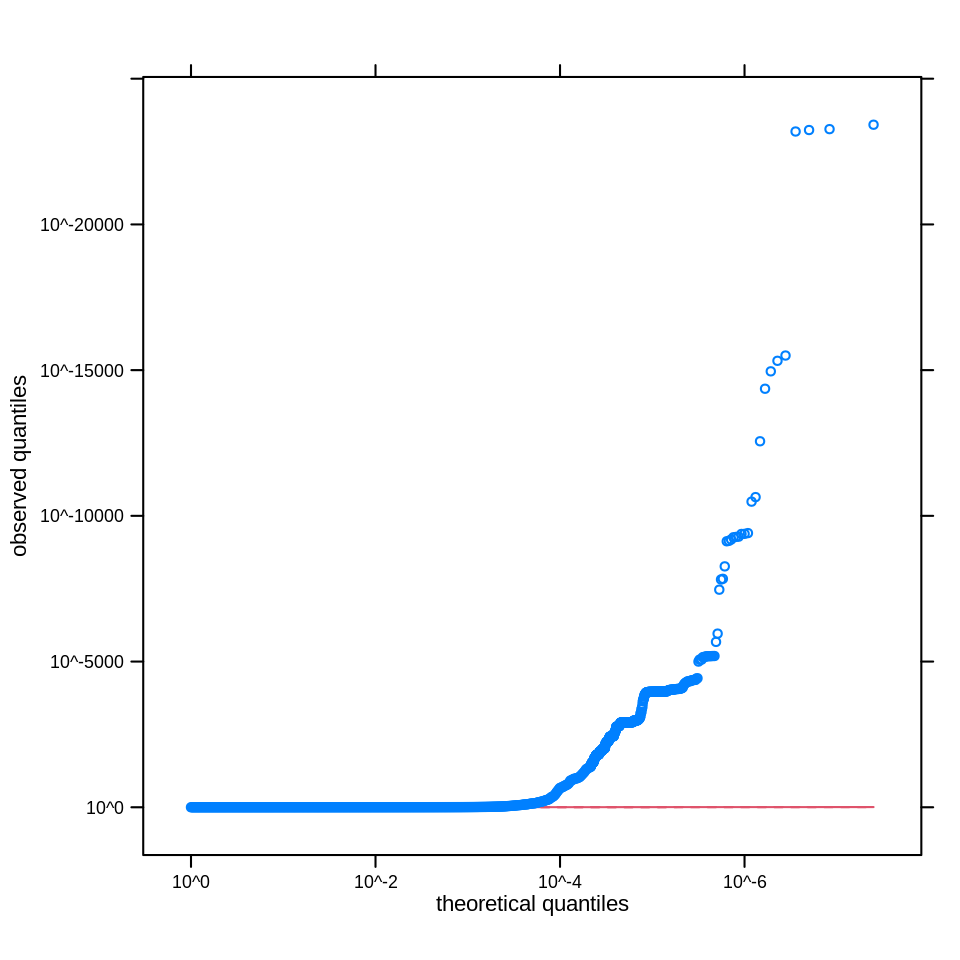

Supplementary Figure 1.2: Q-Q plot of observed versus expected quantiles for the Lp(a) GWAS p-values, where the expected distribution of p-values is uniform under the null hypothesis, plotted on a log scale, and a solid read line is shown with a slope of 1.

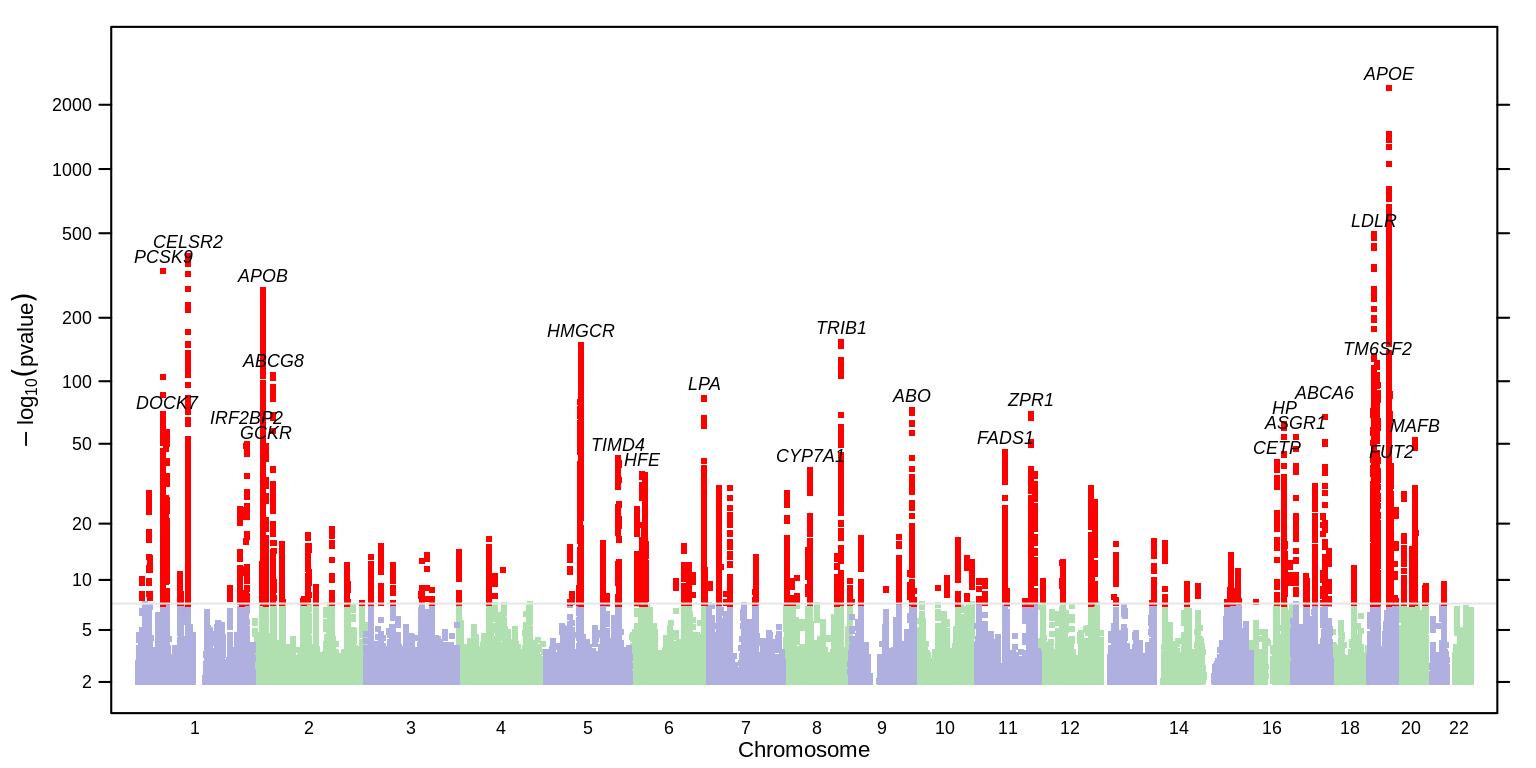

Supplementary Figure 2.1: Manhattan plot showing results from the LDL-C GWAS in UKB training cohort. Top-associated variants at each locus are labeled with their nearest gene context.

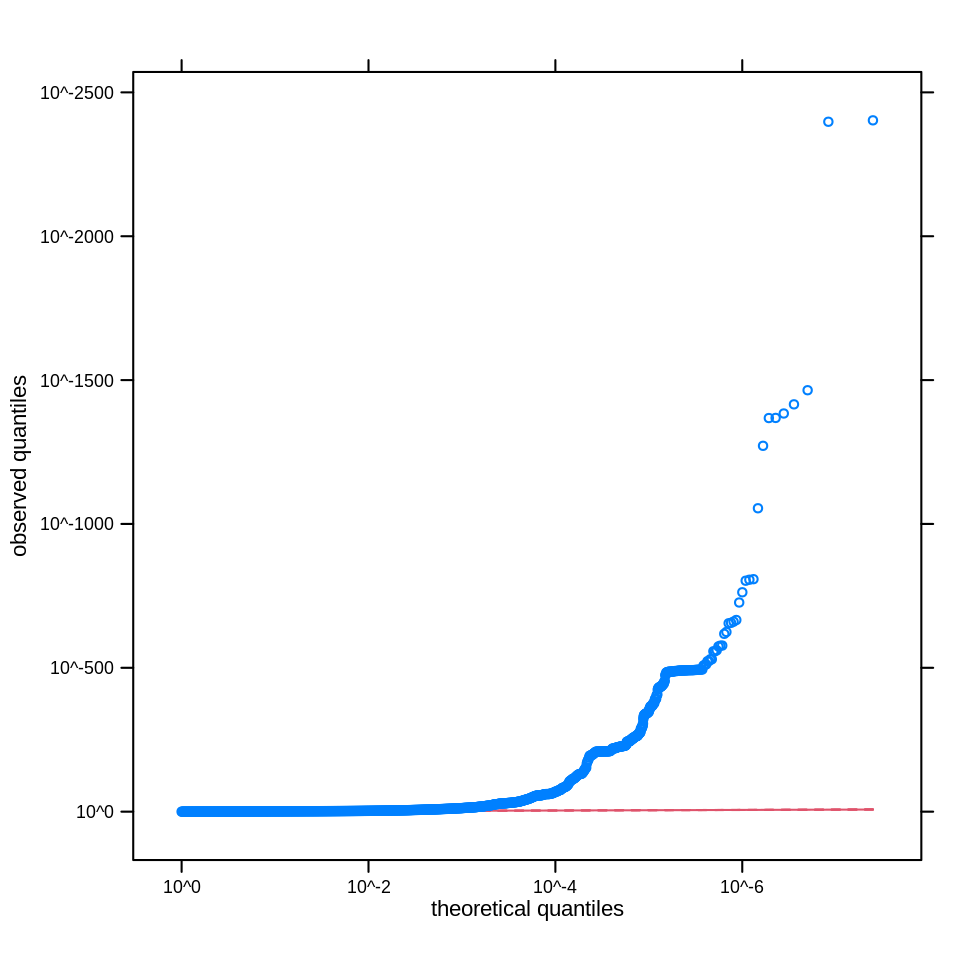

Supplementary Figure 2.2: Q-Q plot of observed versus expected quantiles for the LDL-C GWAS p-values, where the expected distribution of p-values is uniform under the null hypothesis, plotted on a log scale, and a solid read line is shown with a slope of 1.

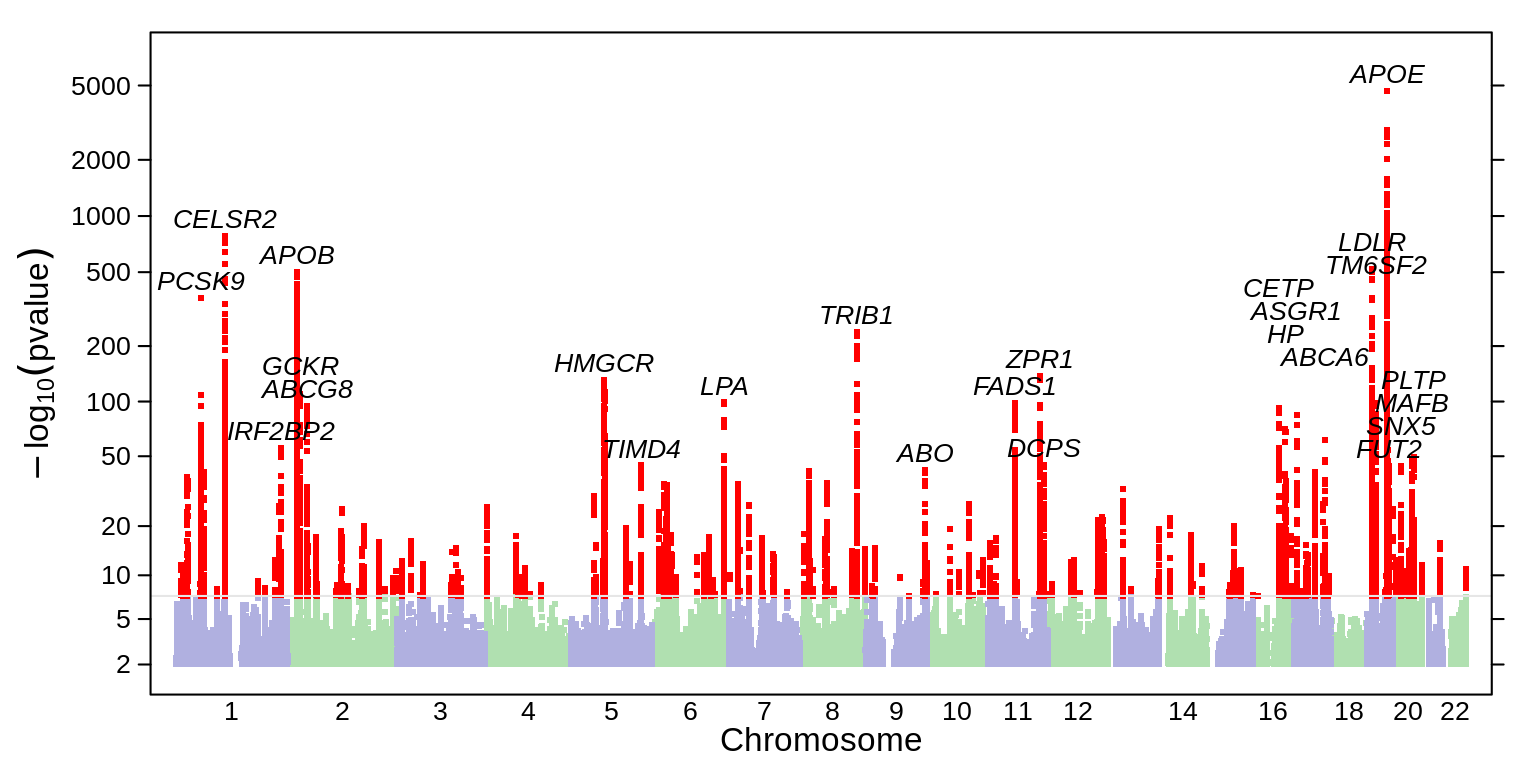

Supplementary Figure 3.1: Manhattan plot showing results from the apoB GWAS in UKB training cohort. Top-associated variants at each locus are labeled with their nearest gene context.

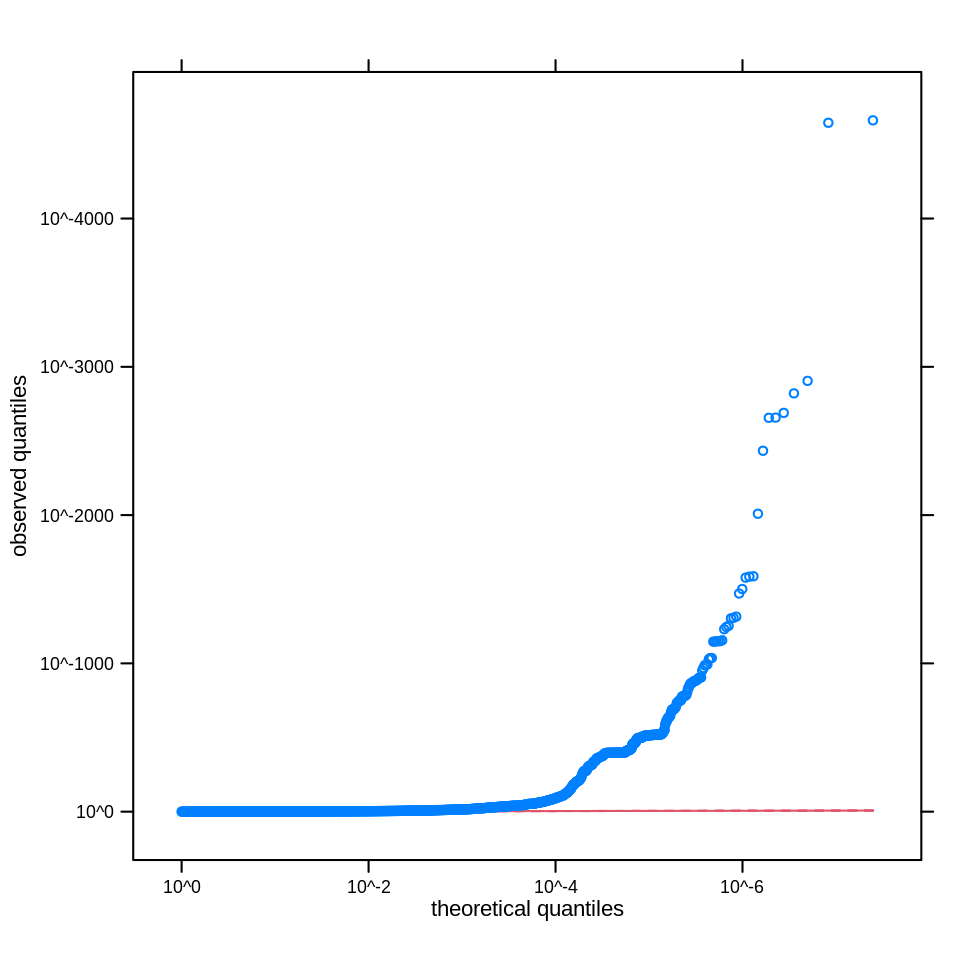

Supplementary Figure 3.2: Q-Q plot of observed versus expected quantiles for the apoB GWAS p-values, where the expected distribution of p-values is uniform under the null hypothesis, plotted on a log scale, and a solid read line is shown with a slope of 1.

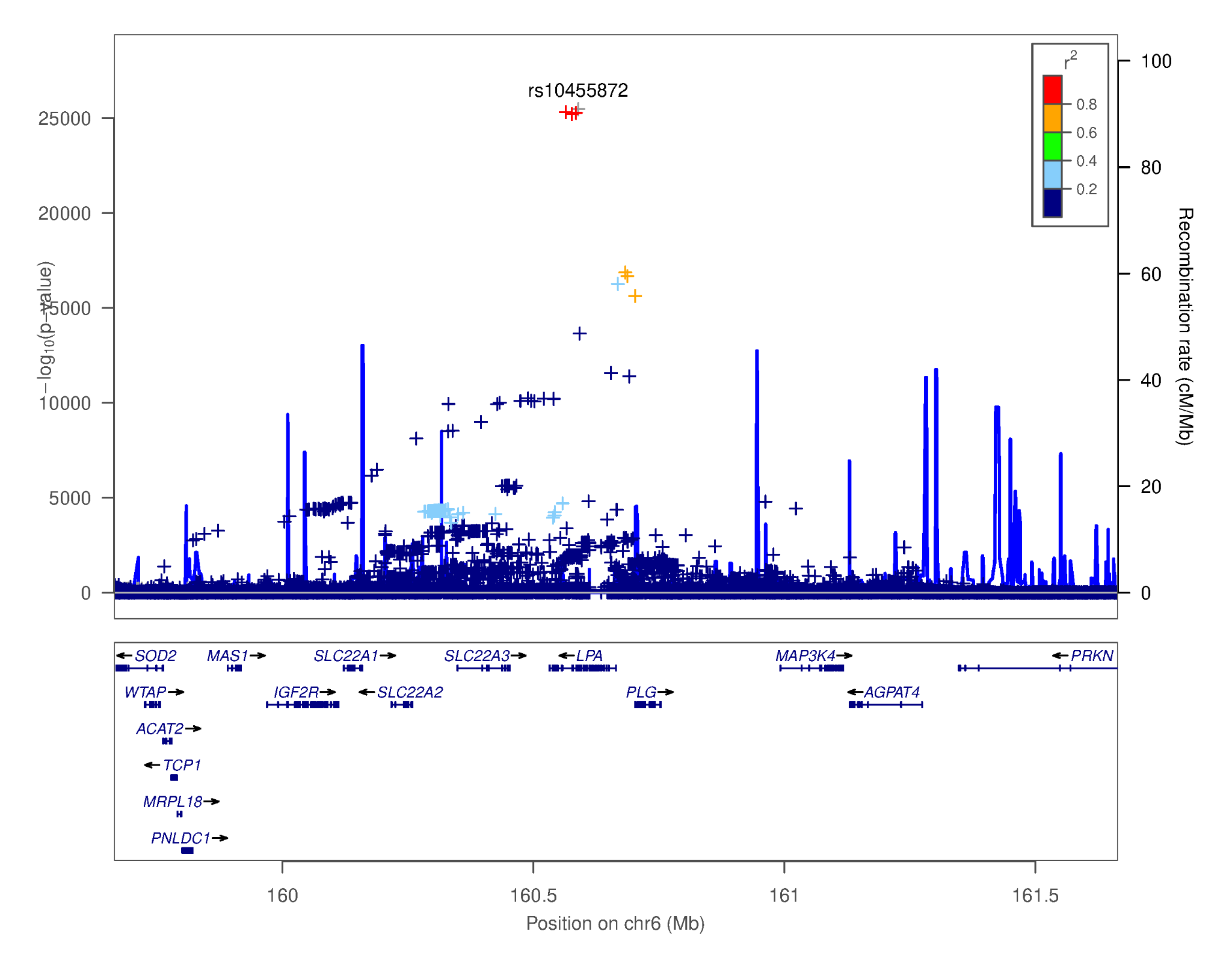

Supplementary Figure 4.1: LocusZoom plot of the Lp(a) GWAS test statistics versus position in the vicinity of the *LPA* gene (GRCh38: chr6: 159,664,259 - 161,664,259). In the plot, a ‘+’ indicates an imputed non-coding variant, whereas a ‘x’ indicates an imputed protein-altering variant.

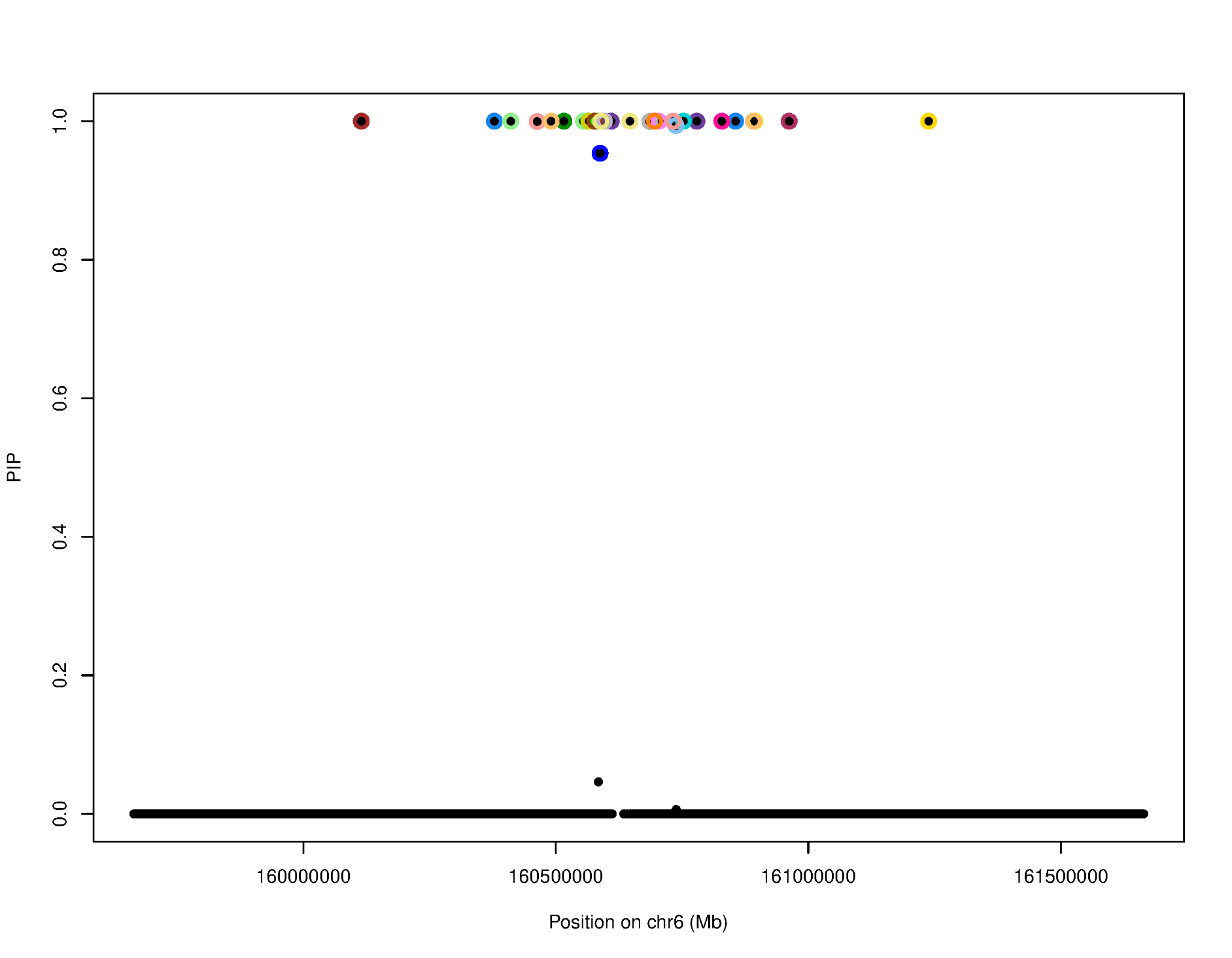

Supplementary Figure 4.2: SuSiE credible sets posterior inclusion probability (PIP) plot of the *LPA* locus. SNPs (represented by the black dots) in different credible sets are enclosed by different colored circles.

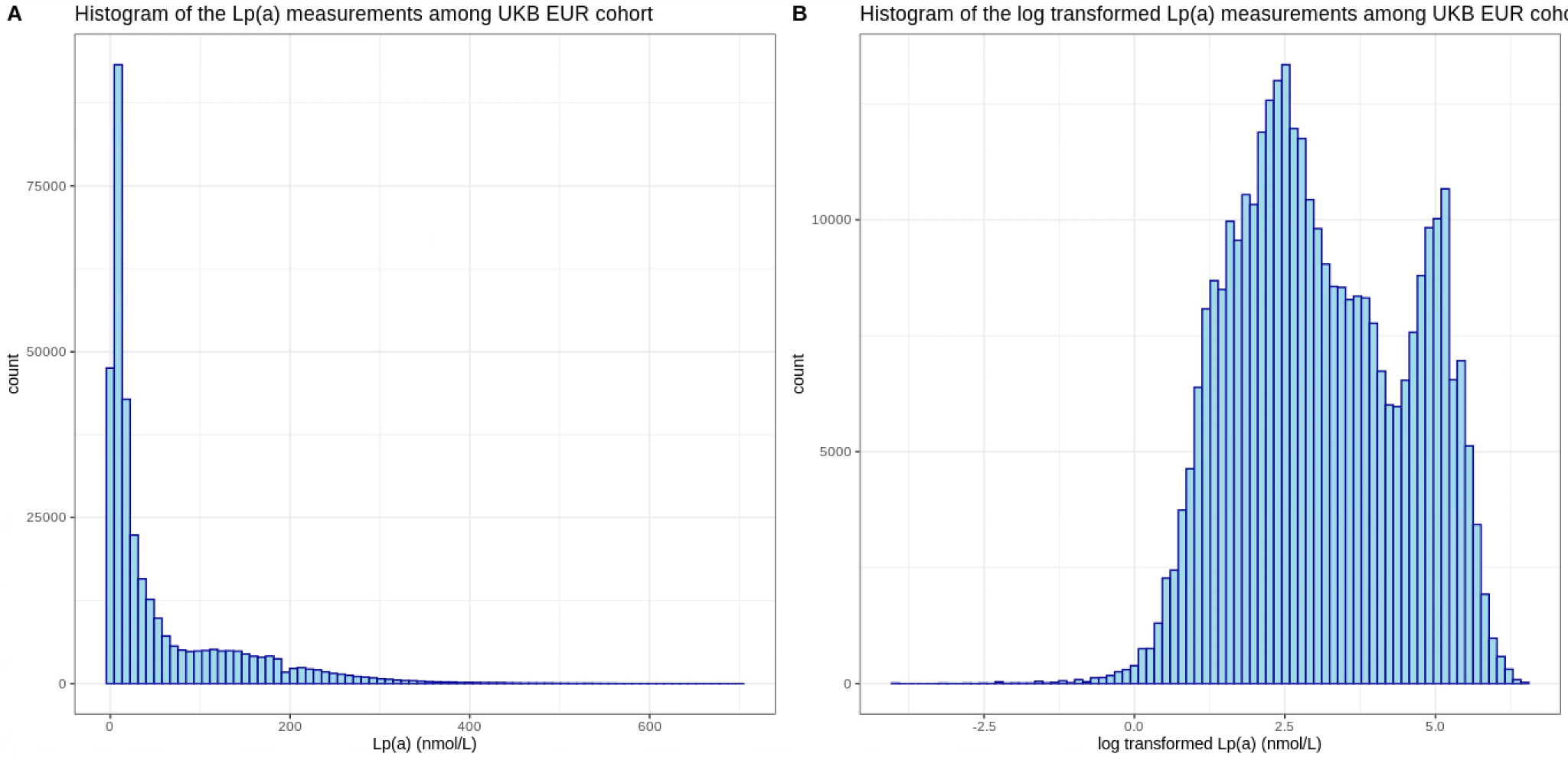

Supplementary Figure 6: Histogram of the Lp(a) distribution in the UKB EUR cohort. Panel A: raw Lp(a) levels; Panel B: log transformed Lp(a) levels.

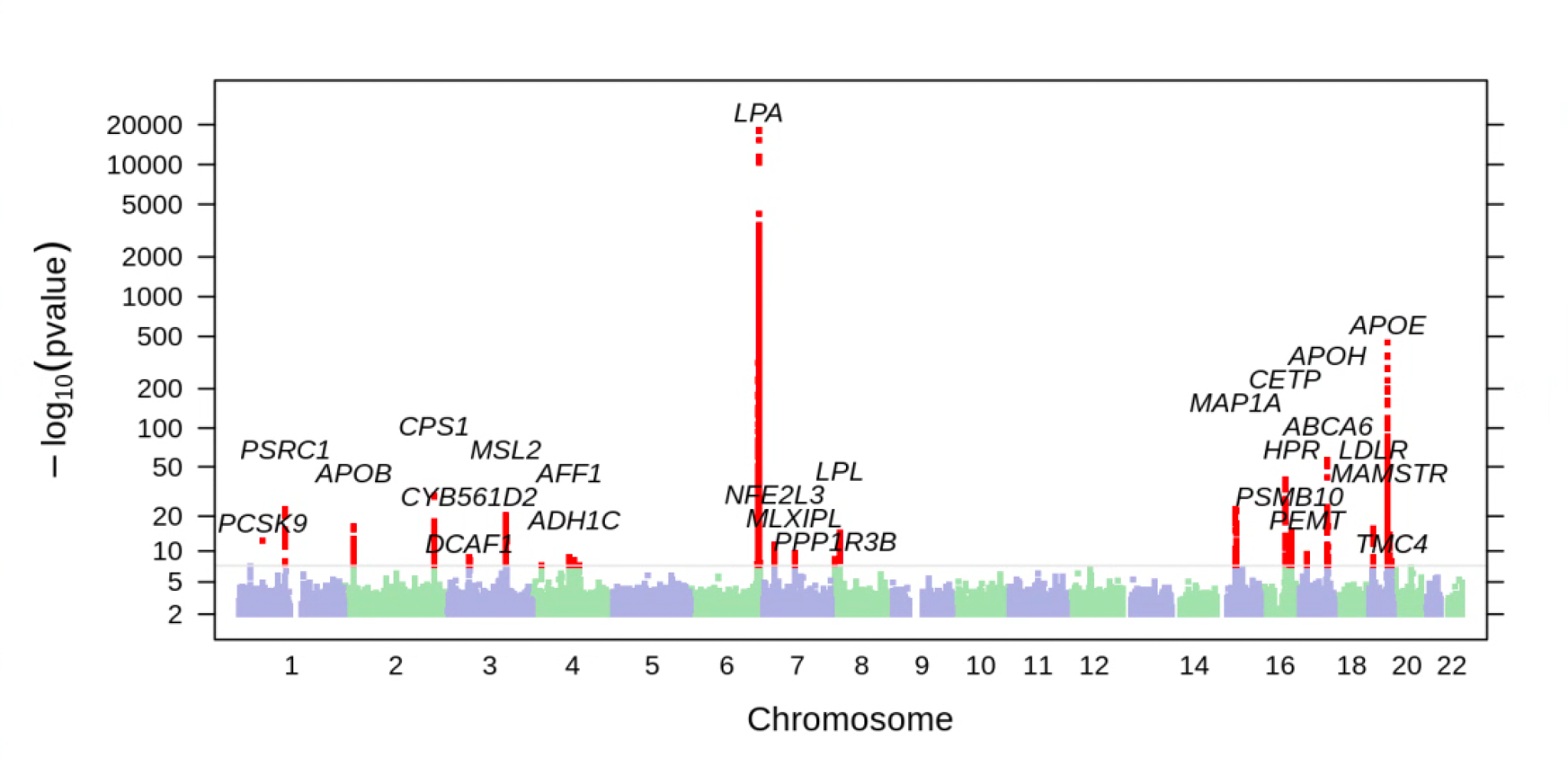

Supplementary Figure 7.1: Manhattan plot showing results from the log transformed Lp(a) GWAS in UKB training cohort. Top-associated variants at each locus are labeled with their nearest gene context.

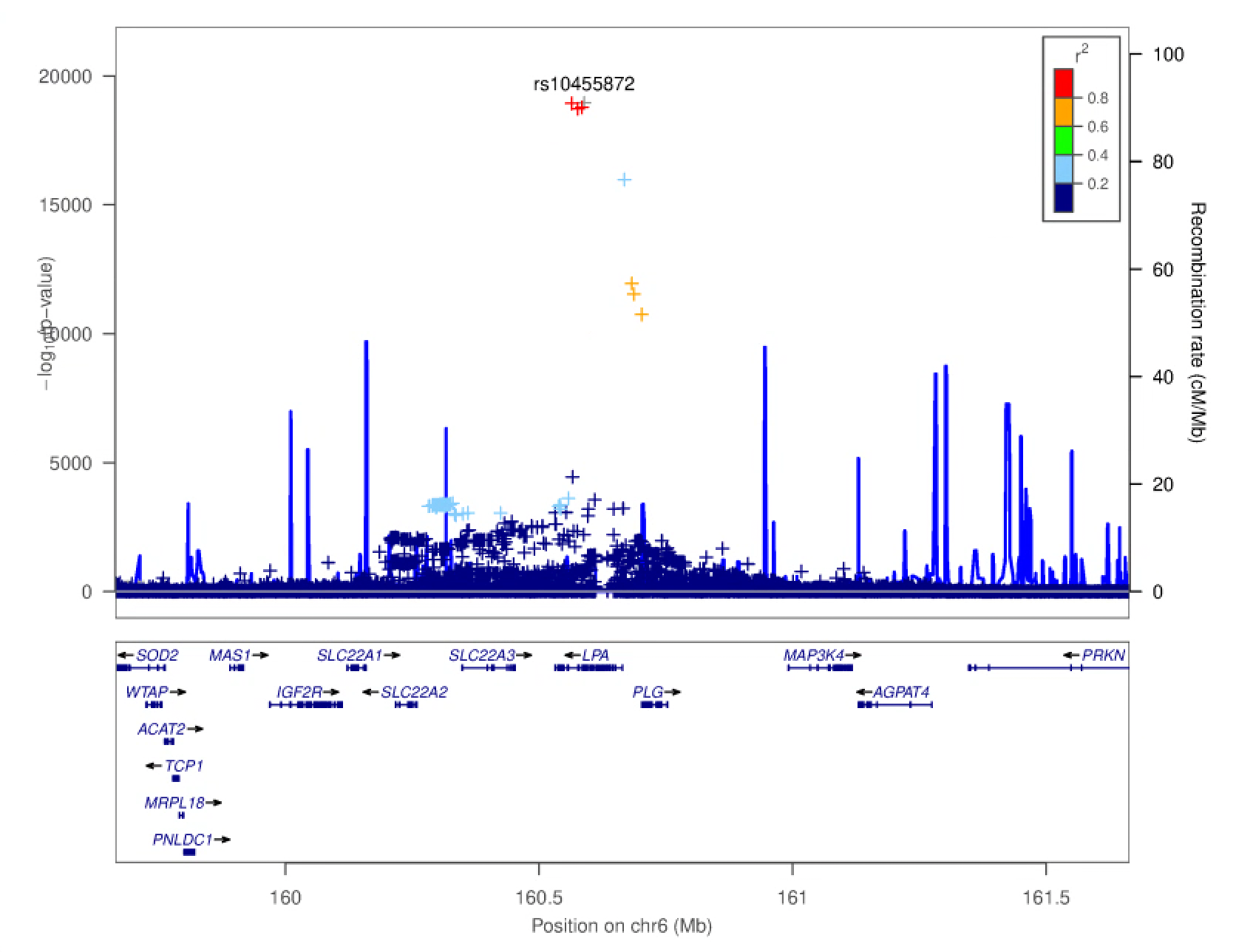

Supplementary Figure 7.2: LocusZoom plot of the log transformed Lp(a) GWAS test statistics versus position in the vicinity of the *LPA* gene (GRCh38: chr6: 159,664,259 - 161,664,259). In the plot, a ‘+’ indicates an imputed non-coding variant, whereas a ‘x’ indicates an imputed protein-altering variant.
