## Supplementary Figures for "Evaluating genetically-predicted causal effects of lipoprotein(a) in human diseases: a phenome-wide Mendelian randomization study": Supplementary_Figure_8.docx

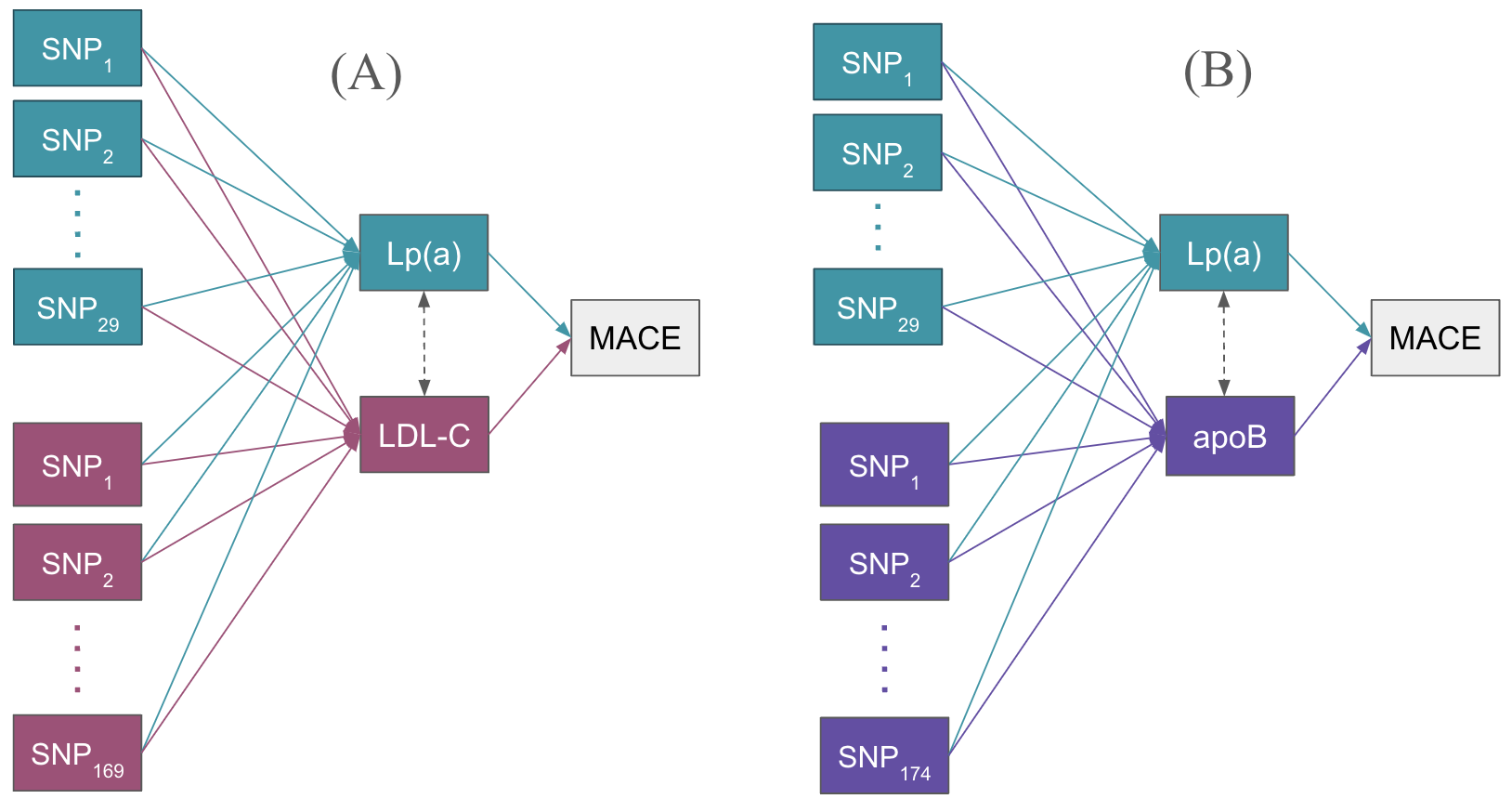

Supplementary Figure 8: **GRS instrument selection scheme for multivariable MR.** (A) Illustration of SNPs used in multivariable MR framework to obtain genetically-predicted independent causal effects of Lp(a) accounting for LDL-C. SNP_1_ to SNP_29_ enclosed in the green box represent the 29 genetic instruments used in the *LPA* GRS for univariable MR. SNP_1_ to SNP_169_ enclosed in the red box represent the 169 genetic instruments used in the LDL-C GRS for univariable MR. The green arrows denote the SNP weights for each genetic instrument extracted from the Lp(a) genetic association summary statistics based on the UKB data. The red arrows denote the SNP weights for each genetic instrument extracted from the LDL-C GWAS summary statistics based on the UKB data. (B) Illustration of SNPs used in multivariable MR framework to obtain genetically-predicted independent causal effects of Lp(a) accounting for apoB. SNP_1_ to SNP_29_ enclosed in the green box represent the 29 genetic instruments used in the *LPA* GRS for univariable MR. SNP_1_ to SNP_174_ enclosed in the purple box represent the 174 genetic instruments used in the apoB GRS for univariable MR. The green arrows denote the SNP weights for each genetic instrument extracted from the Lp(a) genetic association summary statistics based on the UKB data. The purple arrows denote the SNP weights for each genetic instrument extracted from the apoB GWAS summary statistics based on the UKB data.
