## Supplementary Figures for "Evaluating genetically-predicted causal effects of lipoprotein(a) in human diseases: a phenome-wide Mendelian randomization study": Supplementary_Figure_9.docx

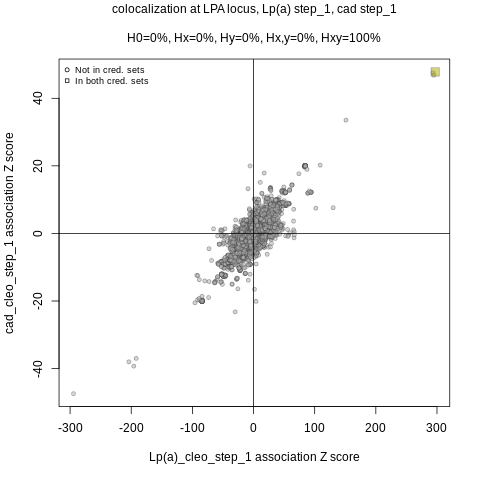

Supplementary Figure 9.1: Colocalization scatter plot of Lp(a) vs cad in the *LPA* locus. The x-axis denotes the z-scores from Lp(a) CLEO step 1 summary statistics; y-axis denotes the z-scores from cad CLEO step 1 summary statistics.

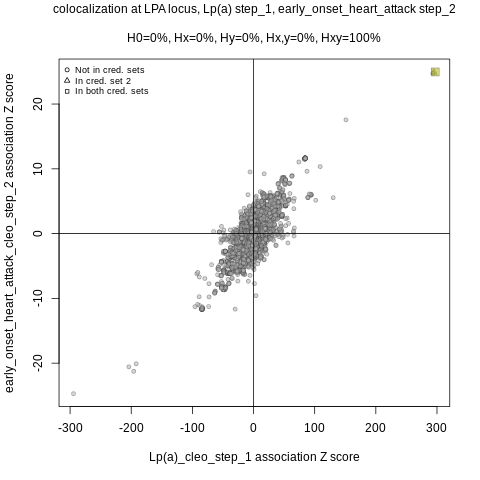

Supplementary Figure 9.2: Colocalization scatter plot of Lp(a) vs early_onset_heart_attack in the *LPA* locus. The x-axis denotes the z-scores from Lp(a) CLEO step 1 summary statistics; y-axis denotes the z-scores from early_onset_heart_attack CLEO step 2 summary statistics.

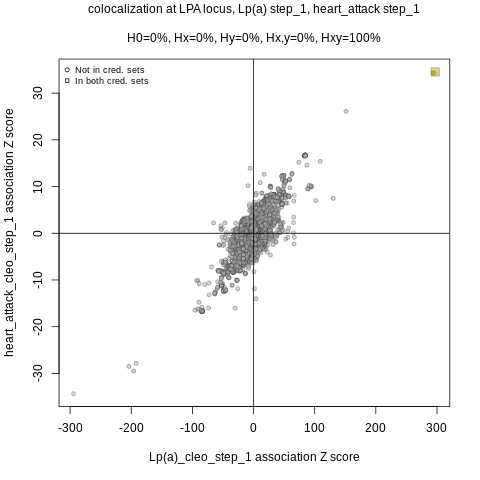

Supplementary Figure 9.3: Colocalization scatter plot of Lp(a) vs heart_attack in the *LPA* locus. The x-axis denotes the z-scores from Lp(a) CLEO step 1 summary statistics; y-axis denotes the z-scores from heart_attack CLEO step 2 summary statistics.

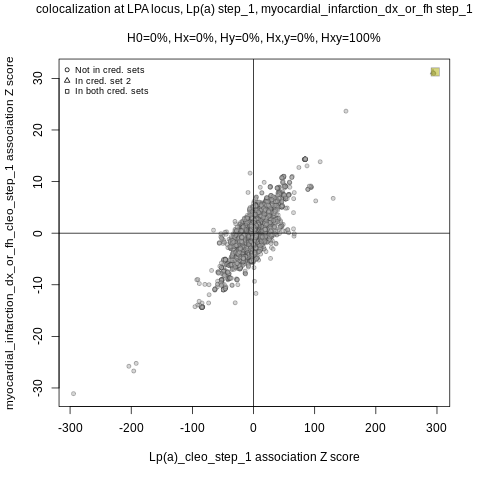

Supplementary Figure 9.4: Colocalization scatter plot of Lp(a) vs myocardial_infarction_dx_or_fh in the *LPA* locus. The x-axis denotes the z-scores from Lp(a) CLEO step 1 summary statistics; y-axis denotes the z-scores from myocardial_infarction_dx_or_fh CLEO step 1 summary statistics.

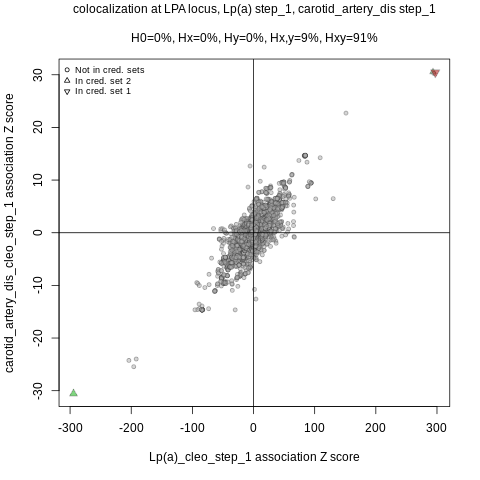

Supplementary Figure 9.5: Colocalization scatter plot of Lp(a) vs carotid_artery_dis in the *LPA* locus. The x-axis denotes the z-scores from Lp(a) CLEO step 1 summary statistics; y-axis denotes the z-scores from carotid_artery_dis CLEO step 1 summary statistics.

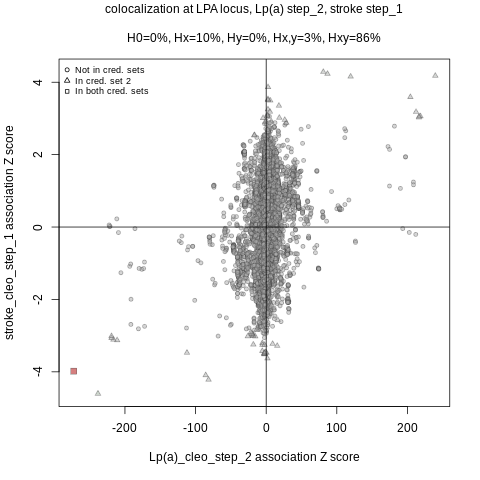

Supplementary Figure 9.6: Colocalization scatter plot of Lp(a) vs stoke in the *LPA* locus. The x-axis denotes the z-scores from Lp(a) CLEO step 2 summary statistics; y-axis denotes the z-scores from stroke CLEO step 1 summary statistics.

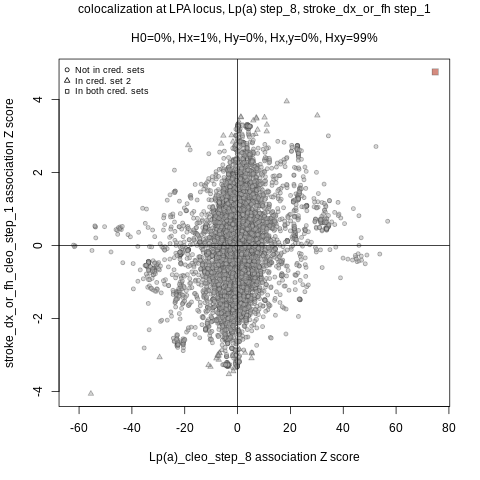

Supplementary Figure 9.7: Colocalization scatter plot of Lp(a) vs stroke_dx_or_fh in the *LPA* locus. The x-axis denotes the z-scores from Lp(a) CLEO step 8 summary statistics; y-axis denotes the z-scores from stroke_dx_or_fh CLEO step 1 summary statistics.

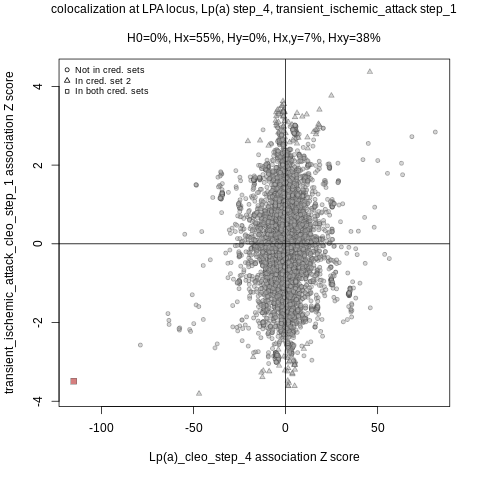

Supplementary Figure 9.8: Colocalization scatter plot of Lp(a) vs transient_ischemic_attack in the *LPA* locus. The x-axis denotes the z-scores from Lp(a) CLEO step 1 summary statistics; y-axis denotes the z-scores from transient_ischemic_attack CLEO step 1 summary statistics.

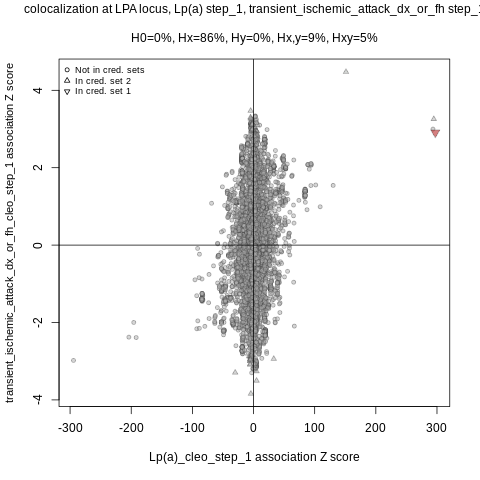

Supplementary Figure 9.9: Colocalization scatter plot of Lp(a) vs transient_ischemic_attack_dx_or_fh in the *LPA* locus. The x-axis denotes the z-scores from Lp(a) CLEO step 1 summary statistics; y-axis denotes the z-scores from transient_ischemic_attack_dx_or_fh CLEO step 1 summary statistics.

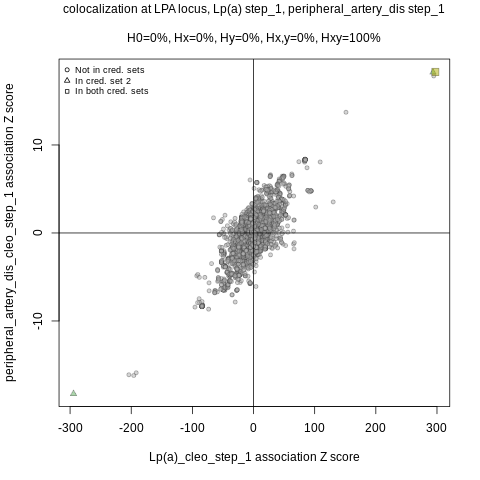

Supplementary Figure 9.10: Colocalization scatter plot of Lp(a) vs peripheral_artery_dis in the *LPA* locus. The x-axis denotes the z-scores from Lp(a) CLEO step 1 summary statistics; y-axis denotes the z-scores from peripheral_artery_dis CLEO step 1 summary statistics.

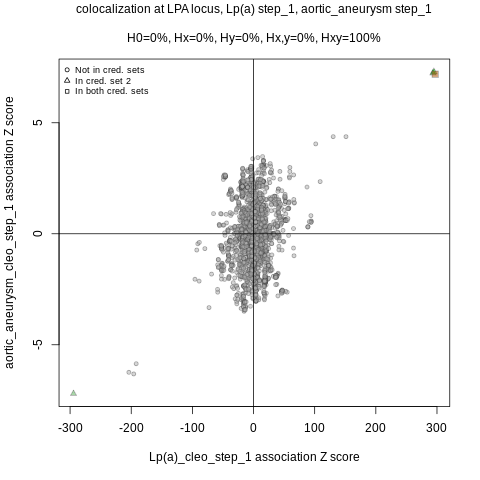

Supplementary Figure 9.11: Colocalization scatter plot of Lp(a) vs aortic_aneurysm in the *LPA* locus. The x-axis denotes the z-scores from Lp(a) CLEO step 1 summary statistics; y-axis denotes the z-scores from aortic_aneurysm CLEO step 1 summary statistics.

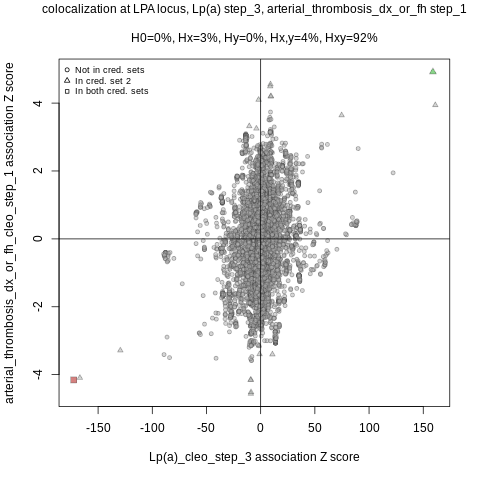

Supplementary Figure 9.12: Colocalization scatter plot of Lp(a) vs arterial_thrombosis_dx_or_fh in the *LPA* locus. The x-axis denotes the z-scores from Lp(a) CLEO step 1 summary statistics; the y-axis denotes the z-scores from arterial_thrombosis_dx_or_fh CLEO step 1 summary statistics.

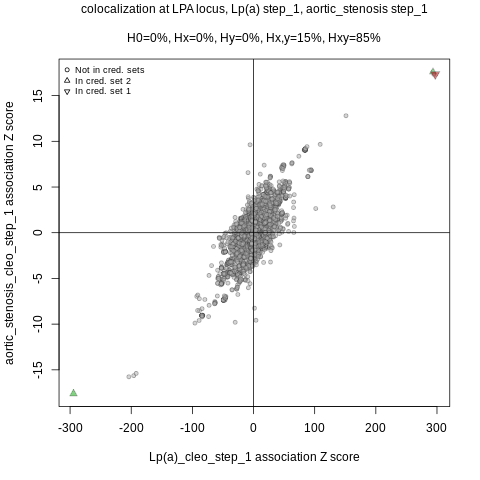

Supplementary Figure 9.13: Colocalization scatter plot of Lp(a) vs aortic_stenosis in the *LPA* locus. The x-axis denotes the z-scores from Lp(a) CLEO step 1 summary statistics; y-axis denotes the z-scores from aortic_stenosis CLEO step 1 summary statistics.

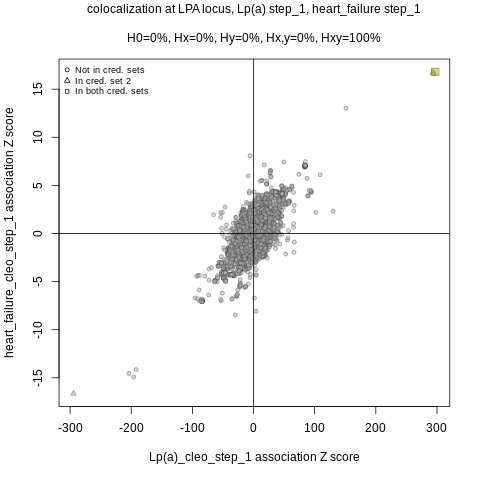

Supplementary Figure 9.14: Colocalization scatter plot of Lp(a) vs heart_failure in the *LPA* locus. The x-axis denotes the z-scores from Lp(a) CLEO step 1 summary statistics; y-axis denotes the z-scores from heart_failure CLEO step 1 summary statistics.

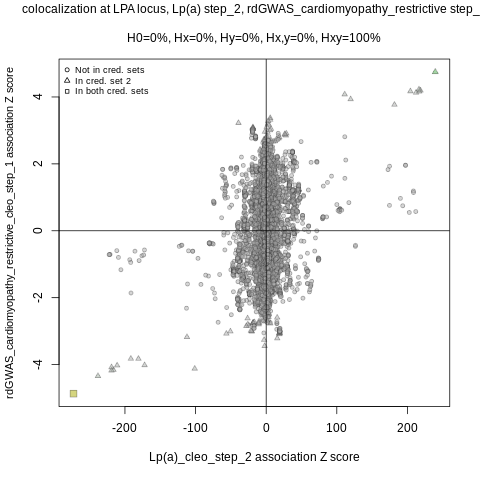

Supplementary Figure 9.15: Colocalization scatter plot of Lp(a) vs rdGWAS_cardiomyopathy_restrictive in the *LPA* locus. The x-axis denotes the z-scores from Lp(a) CLEO step 2 summary statistics; y-axis denotes the z-scores from rdGWAS_cardiomyopathy_restrictive CLEO step 1 summary statistics.

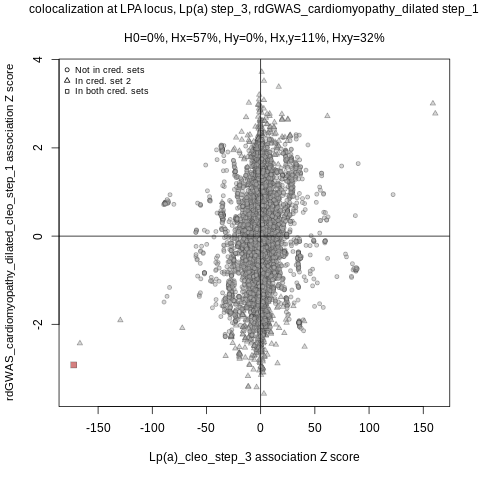

Supplementary Figure 9.16: Colocalization scatter plot of Lp(a) vs rdGWAS_cardiomyopathy_dilated in the *LPA* locus. The x-axis denotes the z-scores from Lp(a) CLEO step 3 summary statistics; y-axis denotes the z-scores from rdGWAS_cardiomyopathy_dilated CLEO step 1 summary statistics.

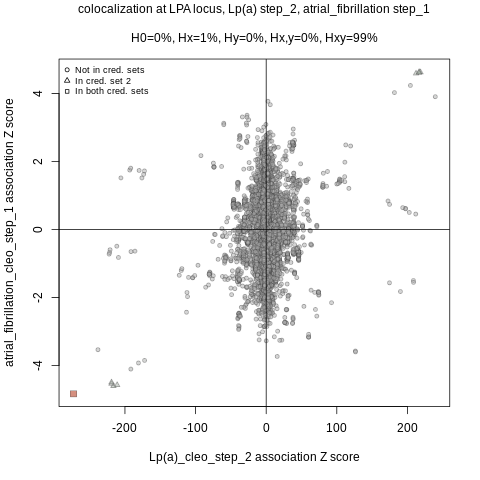

Supplementary Figure 9.17: Colocalization scatter plot of Lp(a) vs atrial_fibrillation in the *LPA* locus. The x-axis denotes the z-scores from Lp(a) CLEO step 2 summary statistics; y-axis denotes the z-scores from atrial_fibrillation CLEO step 1 summary statistics.

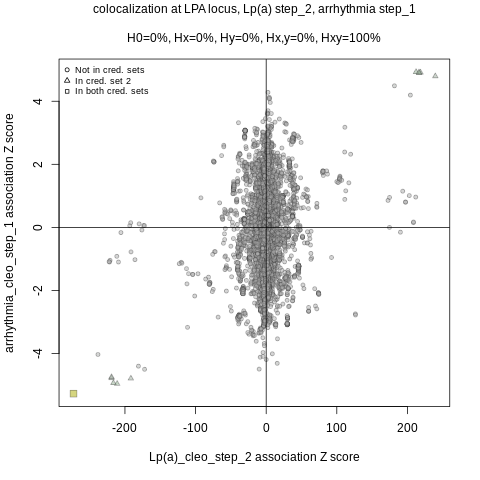

Supplementary Figure 9.18: Colocalization scatter plot of Lp(a) vs atrial_fibrillation in the *LPA* locus. The x-axis denotes the z-scores from Lp(a) CLEO step 2 summary statistics; y-axis denotes the z-scores from arrhythmia CLEO step 1 summary statistics.

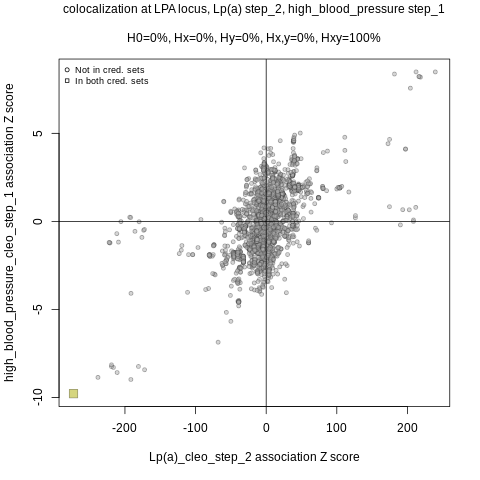

Supplementary Figure 9.19: Colocalization scatter plot of Lp(a) vs high_blood_pressure in the *LPA* locus. The x-axis denotes the z-scores from Lp(a) CLEO step 2 summary statistics; y-axis denotes the z-scores from high_blood_pressure CLEO step 1 summary statistics.

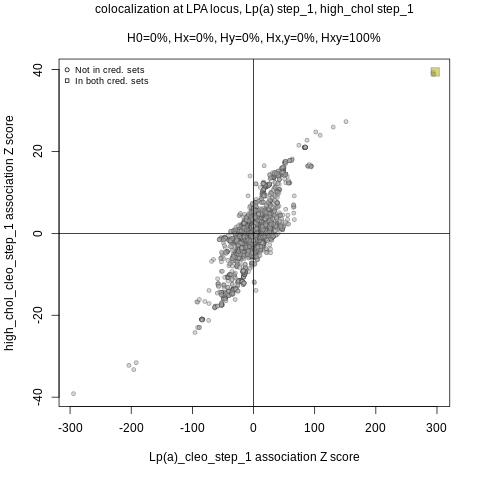

Supplementary Figure 9.20: Colocalization scatter plot of Lp(a) vs high_chol in the *LPA* locus. The x-axis denotes the z-scores from Lp(a) CLEO step 1 summary statistics; y-axis denotes the z-scores from high_chol CLEO step 1 summary statistics.

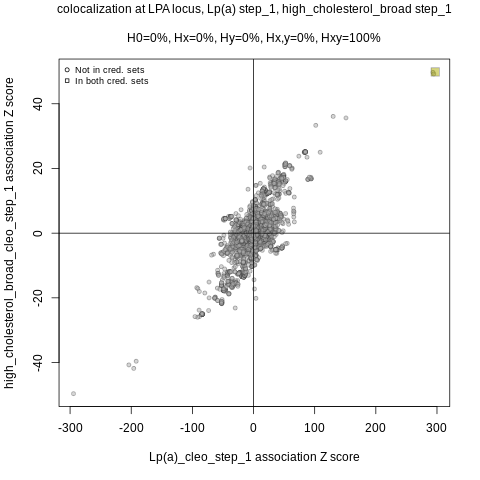

Supplementary Figure 9.21: Colocalization scatter plot of Lp(a) vs high_cholesterol_broad in the *LPA* locus. The x-axis denotes the z-scores from Lp(a) CLEO step 1 summary statistics; y-axis denotes the z-scores from high_cholesterol_broad CLEO step 1 summary statistics.

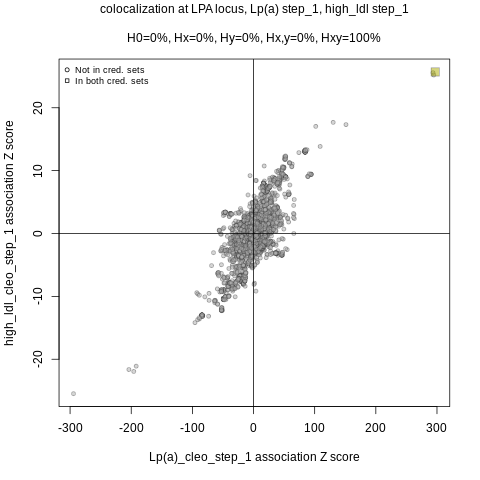

Supplementary Figure 9.22: Colocalization scatter plot of Lp(a) vs high_ldl in the *LPA* locus. The x-axis denotes the z-scores from Lp(a) CLEO step 1 summary statistics; y-axis denotes the z-scores from high_ldl CLEO step 1 summary statistics.

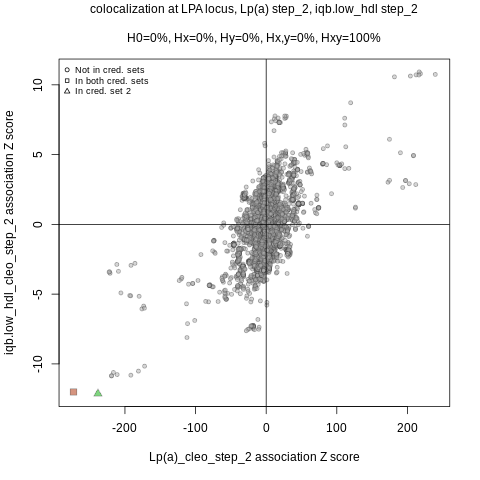

Supplementary Figure 9.23: Colocalization scatter plot of Lp(a) vs iqb.low_hdl in the *LPA* locus. The x-axis denotes the z-scores from Lp(a) CLEO step 2 summary statistics; y-axis denotes the z-scores from iqb.low_hdl CLEO step 2 summary statistics.

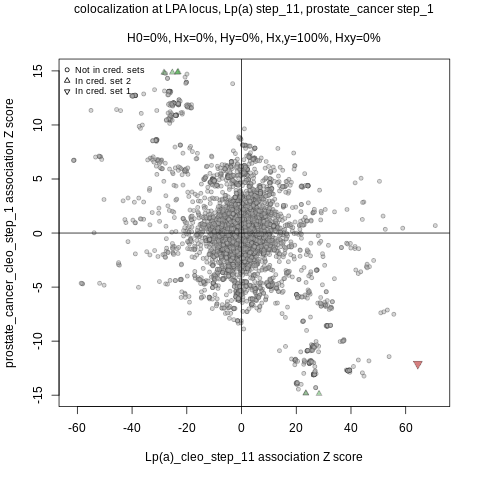

Supplementary Figure 9.24: Colocalization scatter plot of Lp(a) vs prostate_cancer in the *LPA* locus. The x-axis denotes the z-scores from Lp(a) CLEO step 11 summary statistics; y-axis denotes the z-scores from prostate_cancer CLEO step 1 summary statistics.

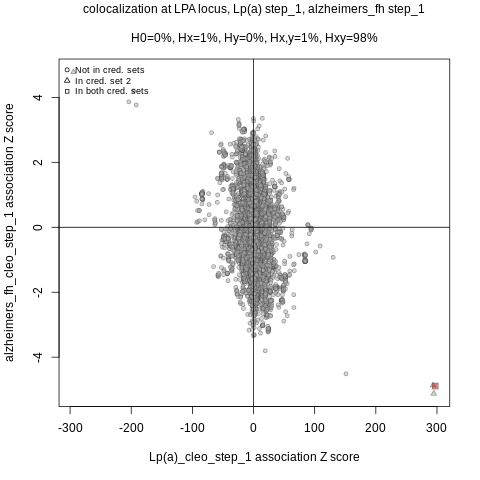

Supplementary Figure 9.25: Colocalization scatter plot of Lp(a) vs alzheimers_fh in the *LPA* locus. The x-axis denotes the z-scores from Lp(a) CLEO step 1 summary statistics; y-axis denotes the z-scores from alzheimers_fh CLEO step 1 summary statistics.
