## Supplementary Figures for "Evaluating genetically-predicted causal effects of lipoprotein(a) in human diseases: a phenome-wide Mendelian randomization study": Supplementary_Figure_11.docx

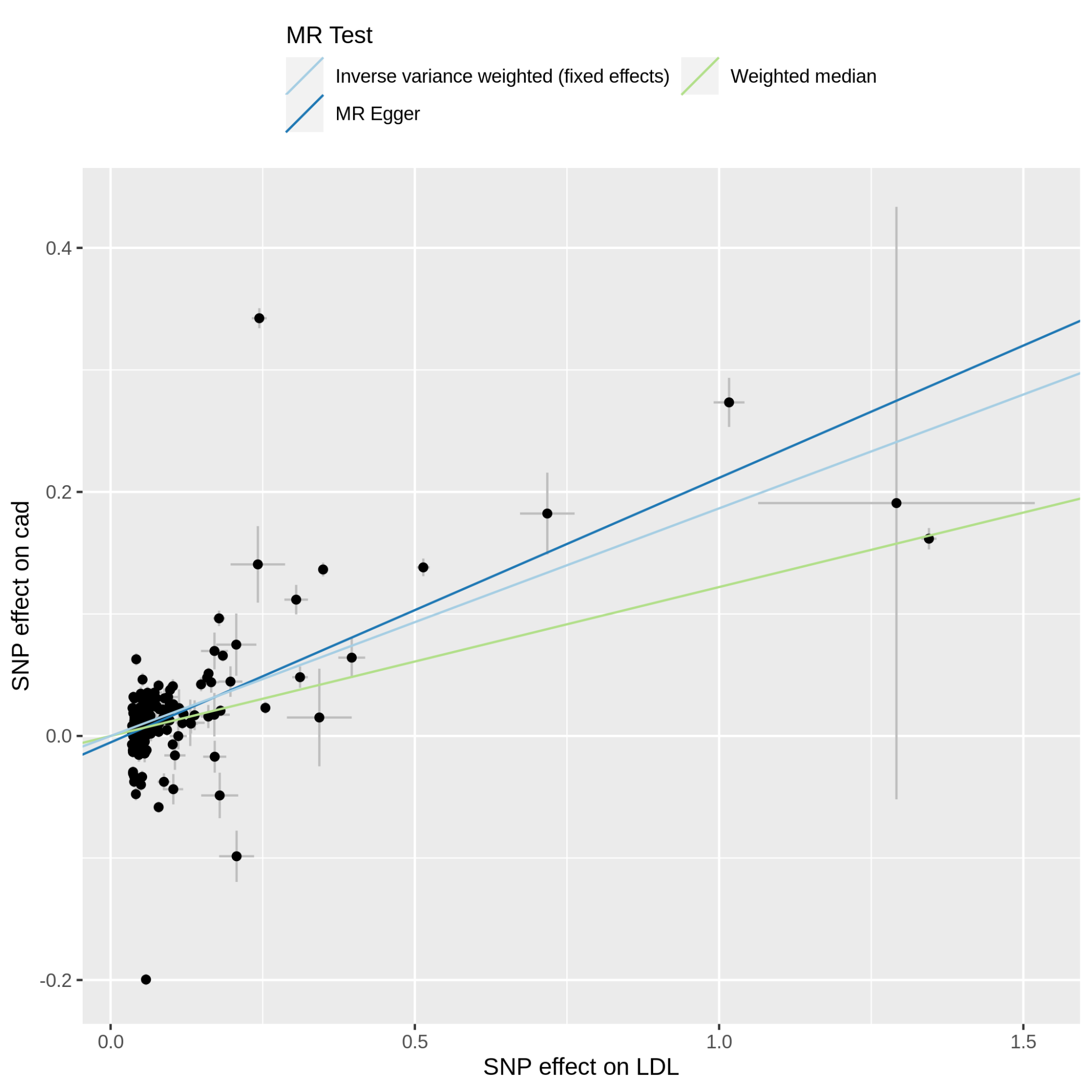

Supplementary Figure 11.1.1: MR scatter plot of LDL-C vs cad, based on the full set of LDL-C genetic instruments. The dot and horizontal bar along the x-axis denotes the GWAS marginal effect size estimate and 95% CI for LDL-C from the UKB cohort; the dot and vertical bar along the y-axis denotes the GWAS marginal effect size estimate and 95% CI for cad from the 23andMe cohort. The colored lines represent the 2SMR (IVW, MR-Egger, weighted median) estimates of genetically instrumented LDL-C onto cad.

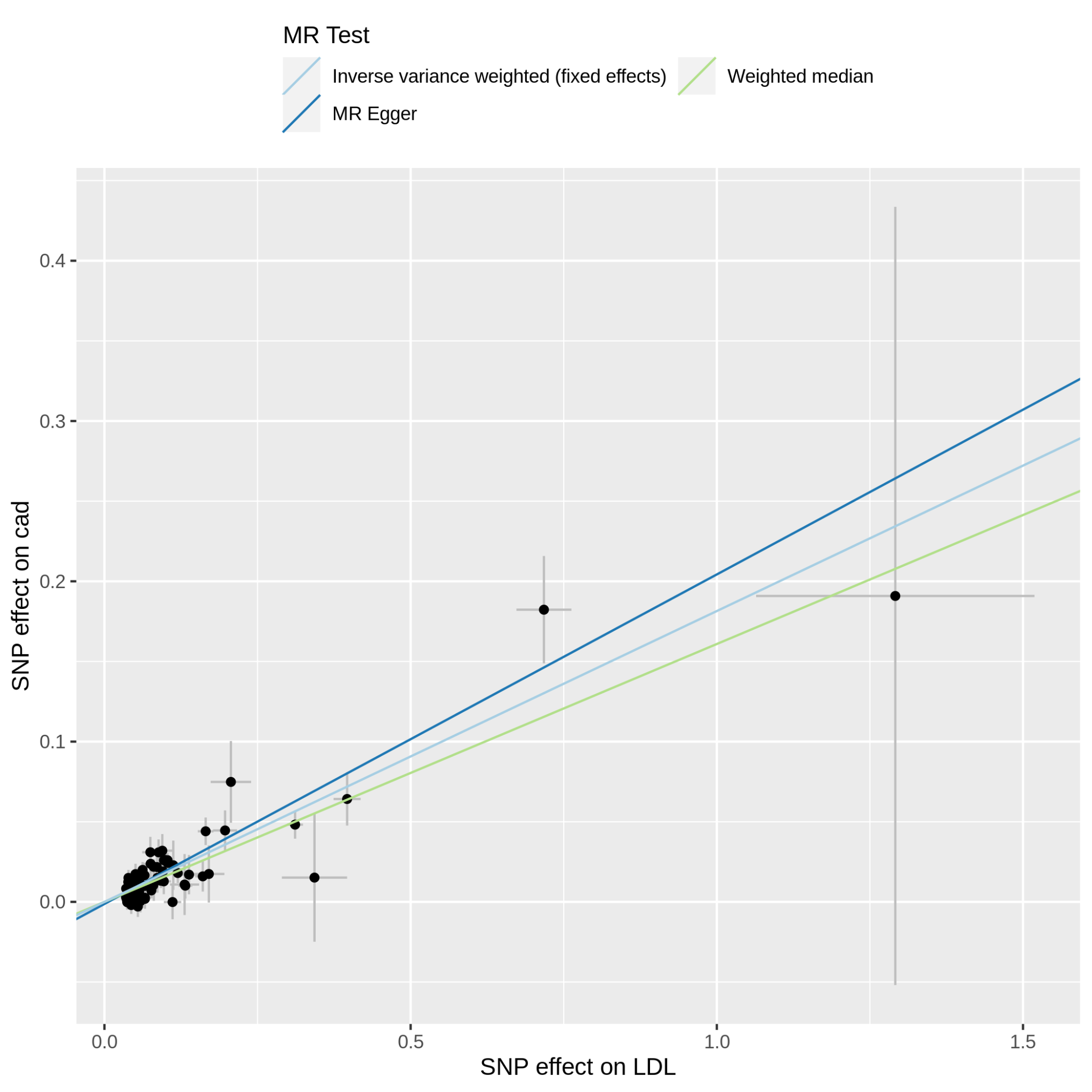

Supplementary Figure 11.1.2: MR scatter plot of LDL-C vs cad, based on the reduced set of LDL-C genetic instruments (after heterogeneity and Steiger filtering). The dot and horizontal bar along the x-axis denotes the GWAS marginal effect size estimate and 95% CI for LDL-C from the UKB cohort; the dot and vertical bar along the y-axis denotes the GWAS marginal effect size estimate and 95% CI for cad from the 23andMe cohort. The colored lines represent the 2SMR (IVW, MR-Egger, weighted median) estimates of genetically instrumented LDL-C onto cad.

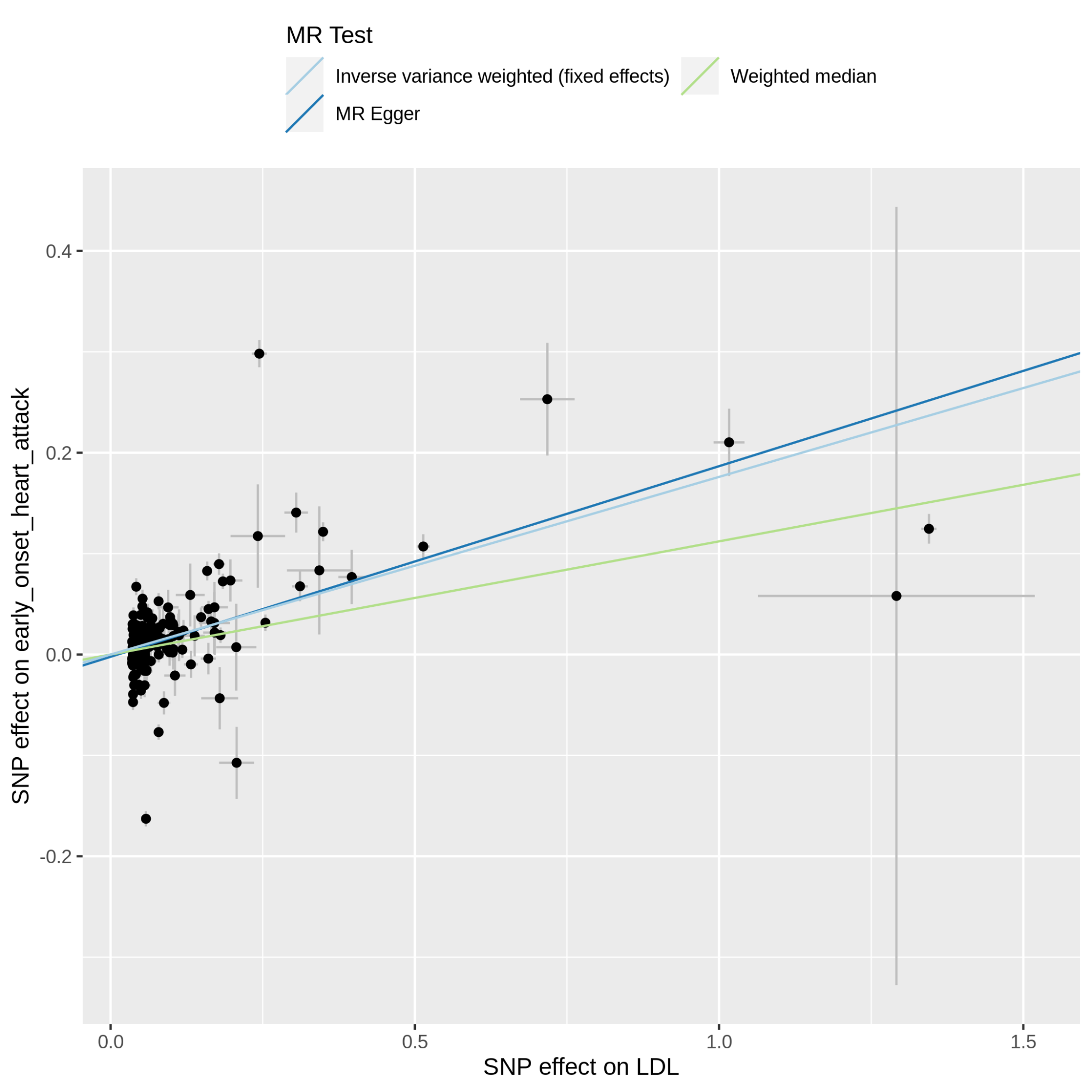

Supplementary Figure 11.2.1: MR scatter plot of LDL-C vs early_onset_heart_attack, based on the full set of LDL-C genetic instruments. The dot and horizontal bar along the x-axis denotes the GWAS marginal effect size estimate and 95% CI for LDL-C from the UKB cohort; the dot and vertical bar along the y-axis denotes the GWAS marginal effect size estimate and 95% CI for early_onset_heart_attack from the 23andMe cohort. The colored lines represent the 2SMR (IVW, MR-Egger, weighted median) estimates of genetically instrumented LDL-C onto early_onset_heart_attack.

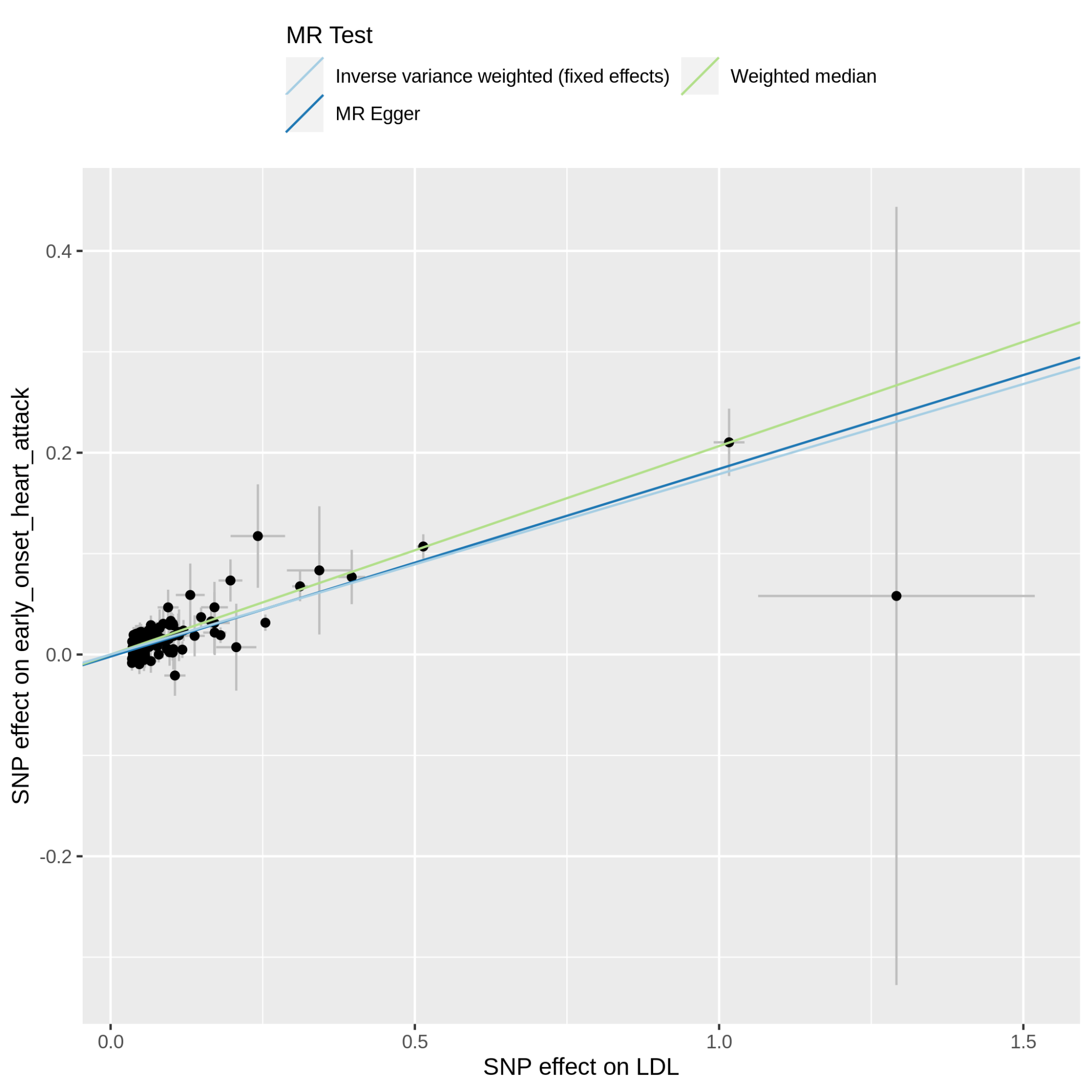

Supplementary Figure 11.2.2: MR scatter plot of LDL-C vs early_onset_heart_attack, based on the reduced set of LDL-C genetic instruments (after heterogeneity and Steiger filtering). The dot and horizontal bar along the x-axis denotes the GWAS marginal effect size estimate and 95% CI for LDL-C from the UKB cohort; the dot and vertical bar along the y-axis denotes the GWAS marginal effect size estimate and 95% CI for early_onset_heart_attack from the 23andMe cohort. The colored lines represent the 2SMR (IVW, MR-Egger, weighted median) estimates of genetically instrumented LDL-C onto early_onset_heart_attack.

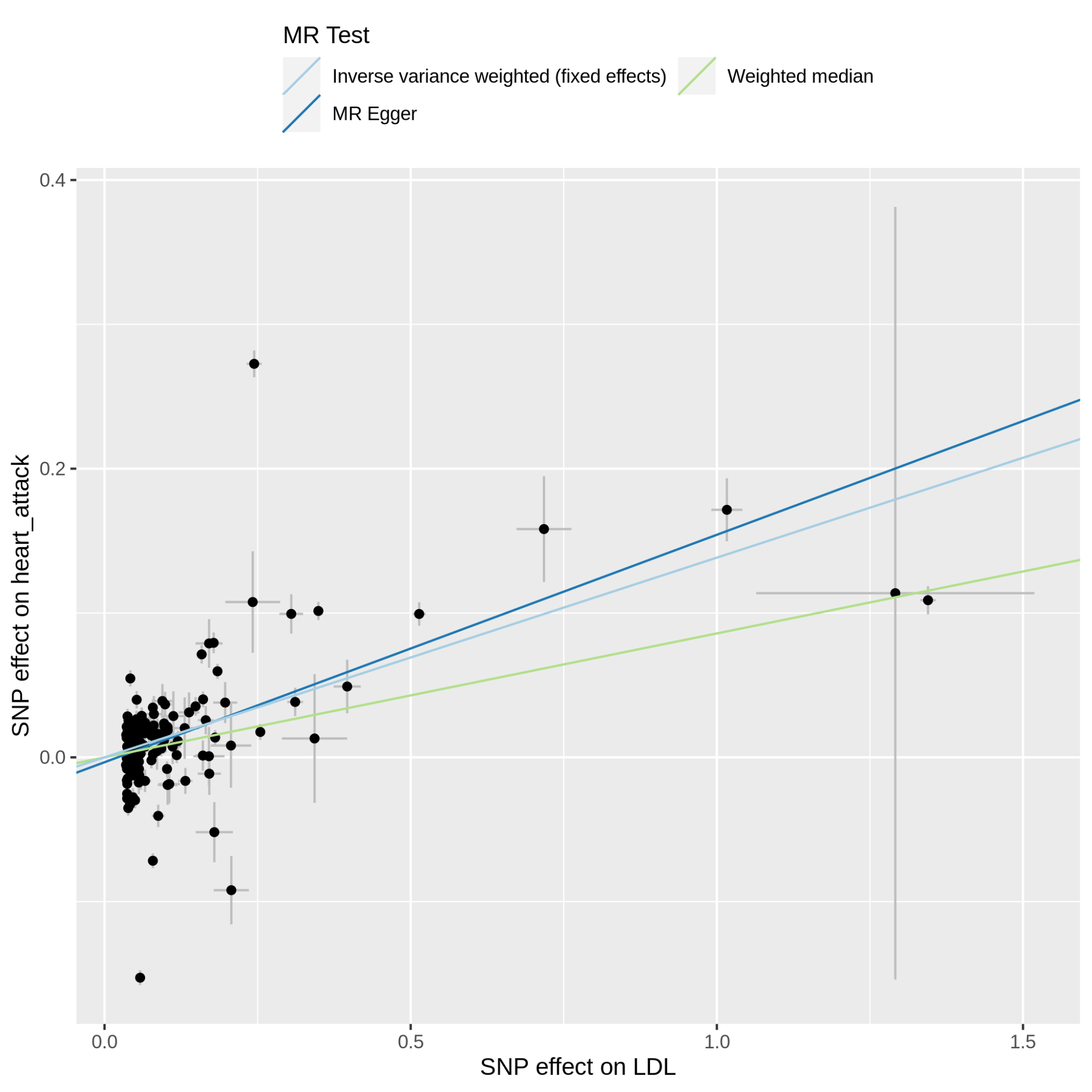

Supplementary Figure 11.3.1: MR scatter plot of LDL-C vs heart_attack, based on the full set of LDL-C genetic instruments. The dot and horizontal bar along the x-axis denotes the GWAS marginal effect size estimate and 95% CI for LDL-C from the UKB cohort; the dot and vertical bar along the y-axis denotes the GWAS marginal effect size estimate and 95% CI for heart_attack from the 23andMe cohort. The colored lines represent the 2SMR (IVW, MR-Egger, weighted median) estimates of genetically instrumented LDL-C onto heart_attack.

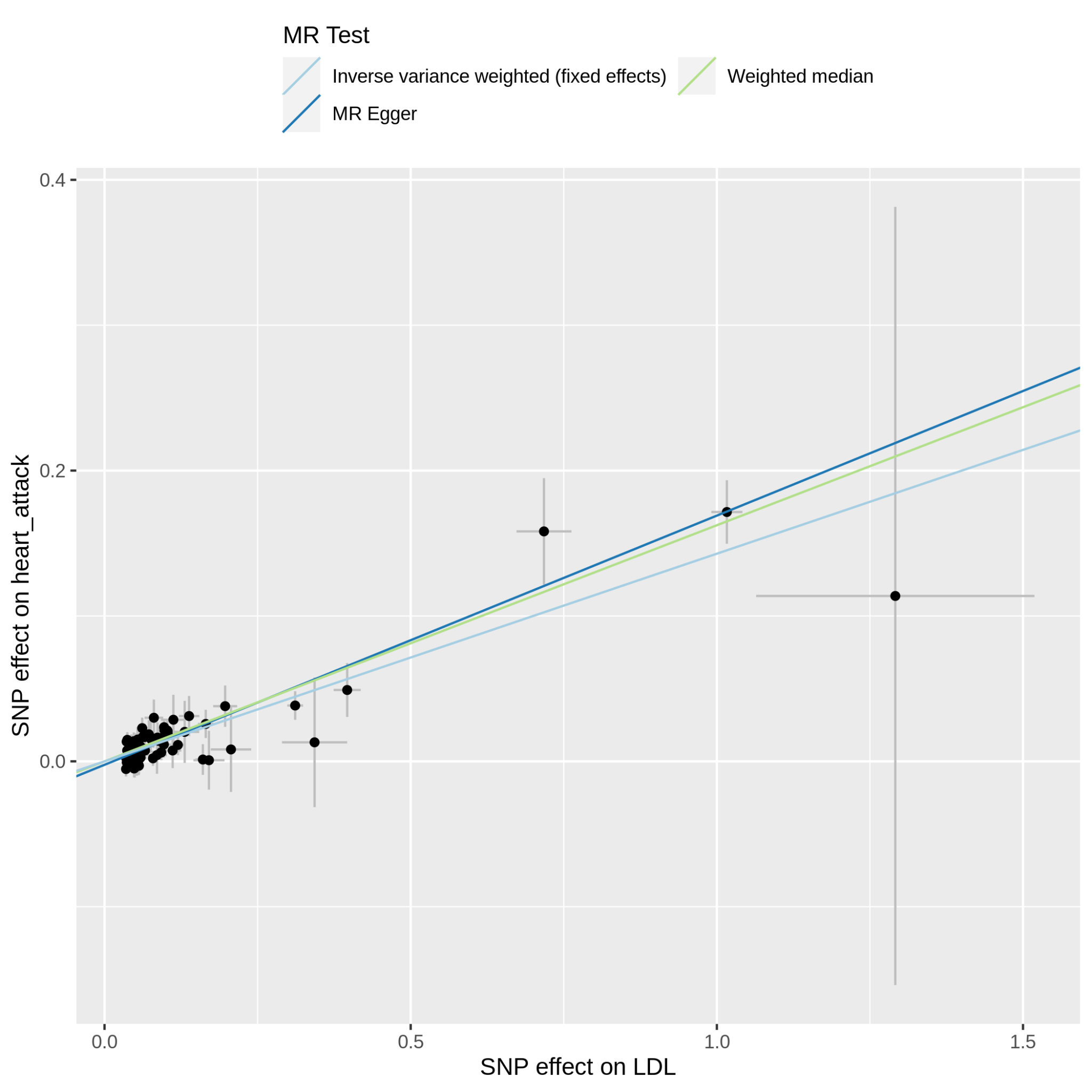

Supplementary Figure 11.3.2: MR scatter plot of LDL-C vs heart_attack, based on the reduced set of LDL-C genetic instruments (after heterogeneity and Steiger filtering). The dot and horizontal bar along the x-axis denotes the GWAS marginal effect size estimate and 95% CI for LDL-C from the UKB cohort; the dot and vertical bar along the y-axis denotes the GWAS marginal effect size estimate and 95% CI for heart_attack from the 23andMe cohort. The colored lines represent the 2SMR (IVW, MR-Egger, weighted median) estimates of genetically instrumented LDL-C onto heart_attack.

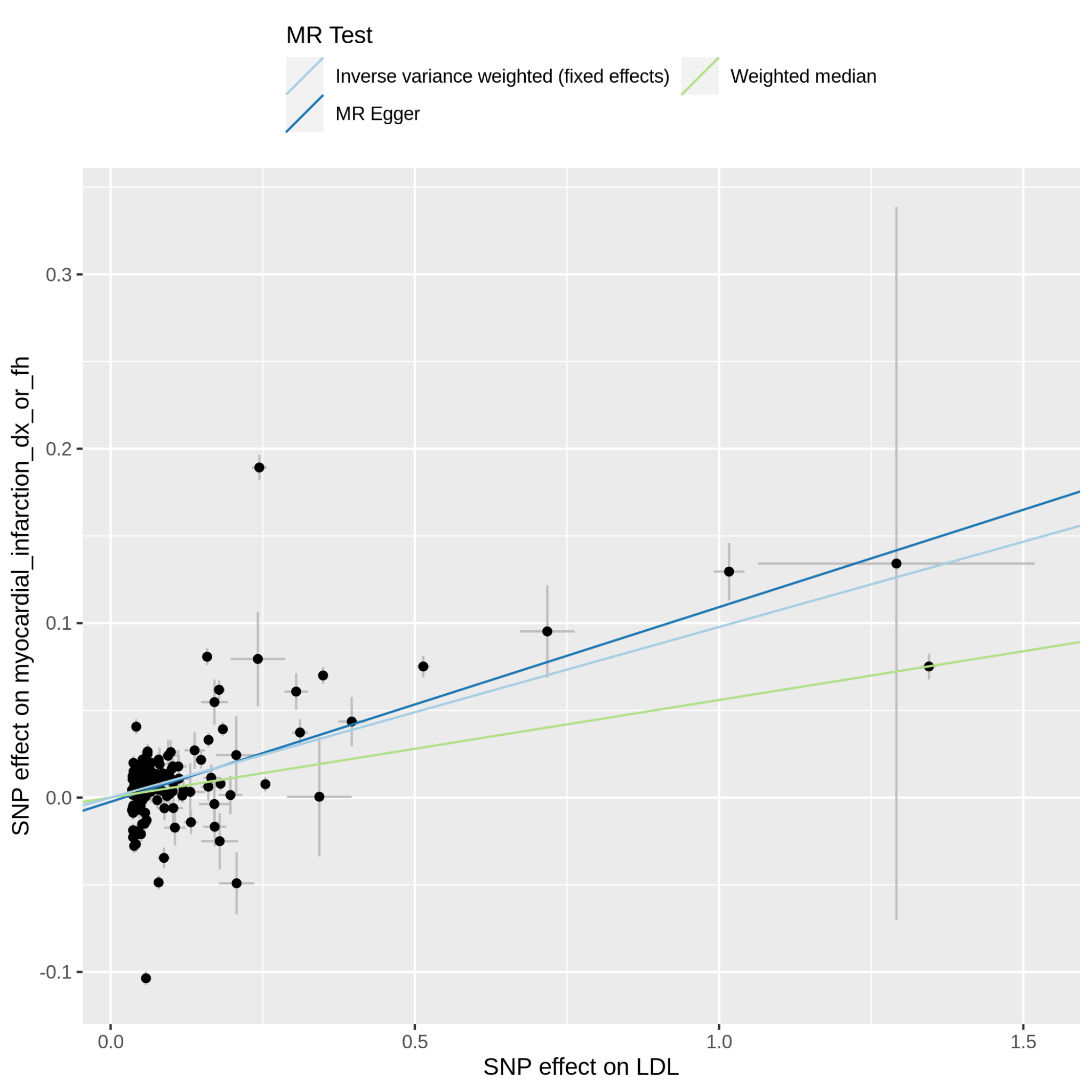

Supplementary Figure 11.4.1: MR scatter plot of LDL-C vs myocardial_infarction_dx_or_fh, based on the full set of LDL-C genetic instruments. The dot and horizontal bar along the x-axis denotes the GWAS marginal effect size estimate and 95% CI for LDL-C from the UKB cohort; the dot and vertical bar along the y-axis denotes the GWAS marginal effect size estimate and 95% CI for myocardial_infarction_dx_or_fh from the 23andMe cohort. The colored lines represent the 2SMR (IVW, MR-Egger, weighted median) estimates of genetically instrumented LDL-C onto myocardial_infarction_dx_or_fh.

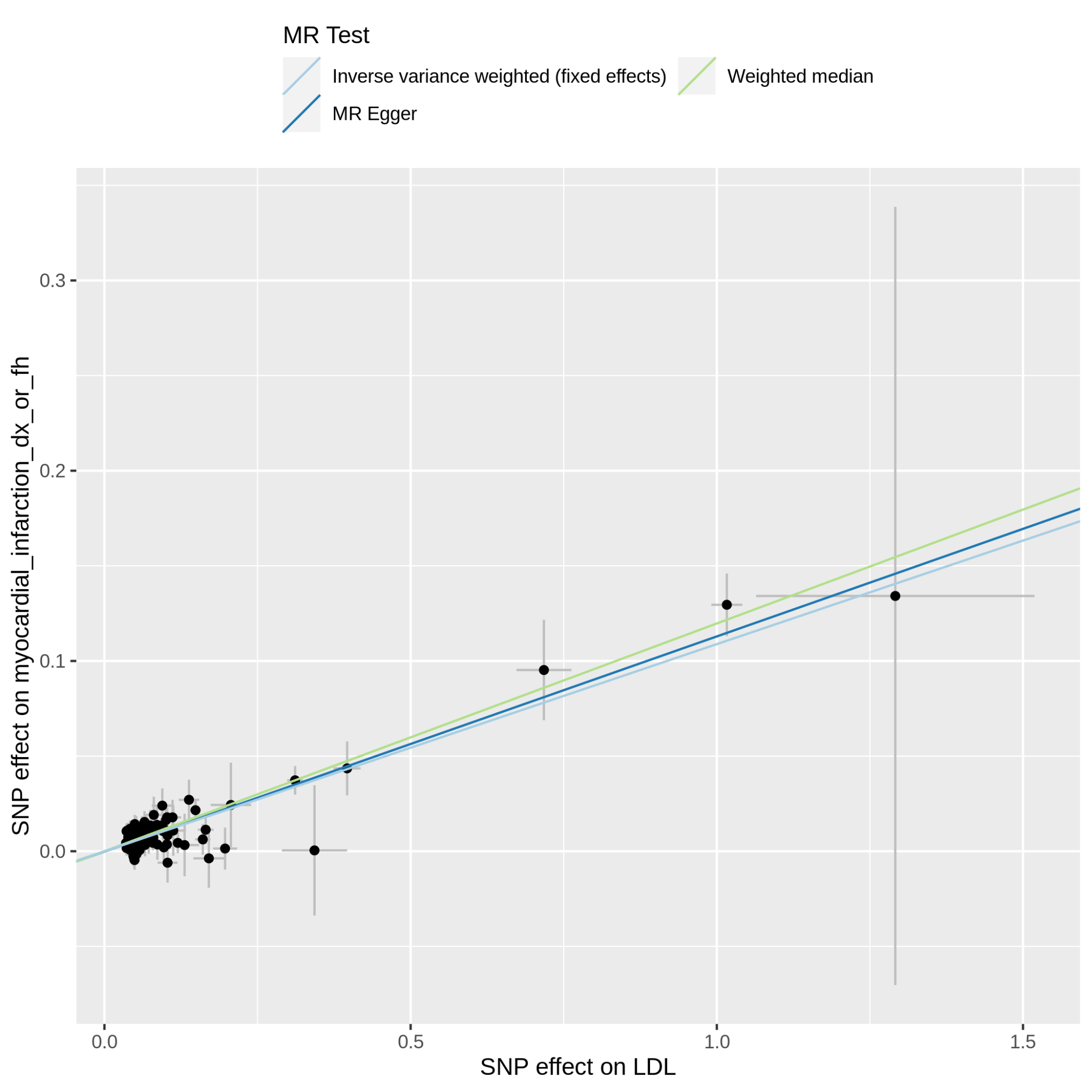

Supplementary Figure 11.4.2: MR scatter plot of LDL-C vs myocardial_infarction_dx_or_fh, based on the reduced set of LDL-C genetic instruments (after heterogeneity and Steiger filtering). The dot and horizontal bar along the x-axis denotes the GWAS marginal effect size estimate and 95% CI for LDL-C from the UKB cohort; the dot and vertical bar along the y-axis denotes the GWAS marginal effect size estimate and 95% CI for myocardial_infarction_dx_or_fh from the 23andMe cohort. The colored lines represent the 2SMR (IVW, MR-Egger, weighted median) estimates of genetically instrumented LDL-C onto myocardial_infarction_dx_or_fh.

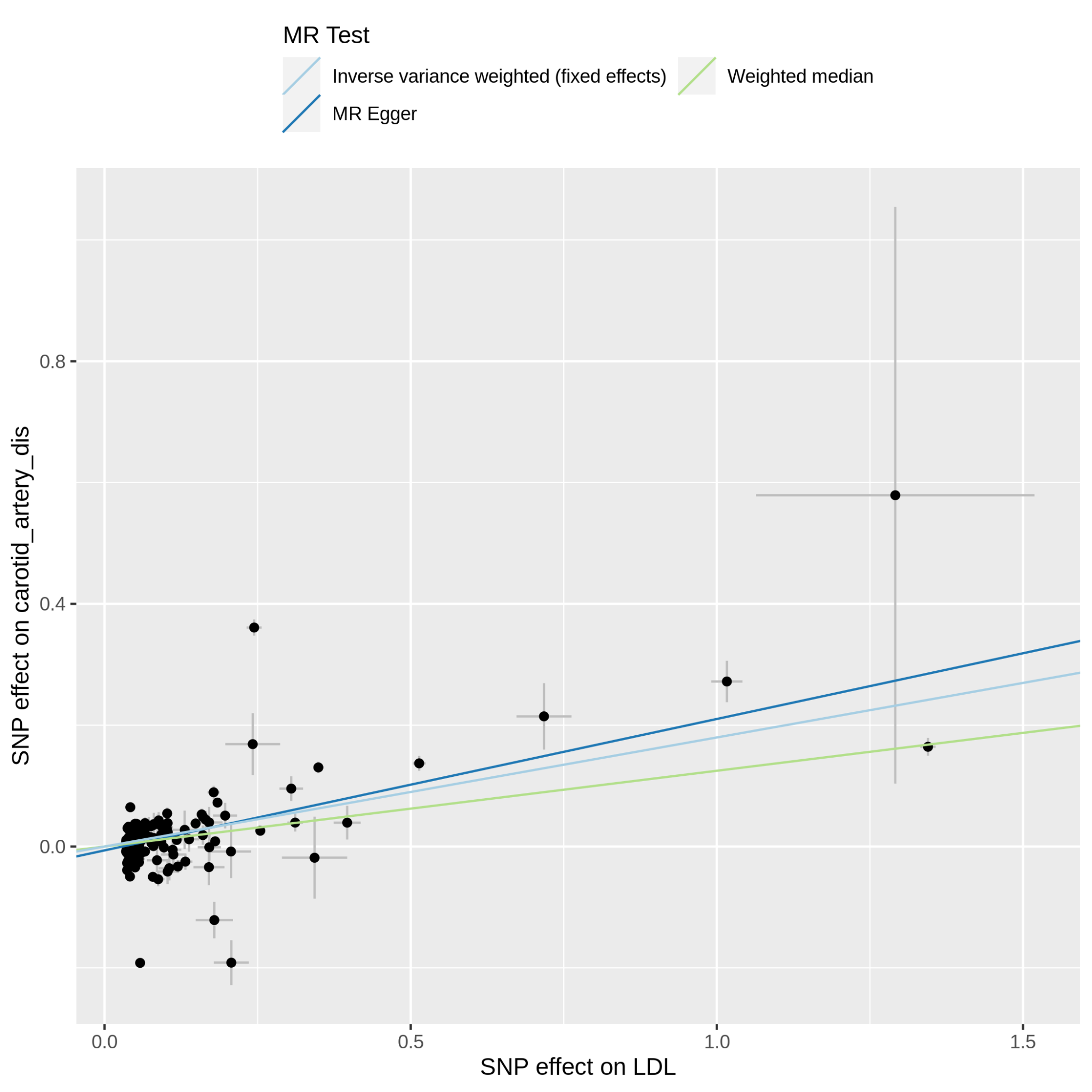

Supplementary Figure 11.5.1: MR scatter plot of LDL-C vs carotid_artery_dis, based on the full set of LDL-C genetic instruments. The dot and horizontal bar along the x-axis denotes the GWAS marginal effect size estimate and 95% CI for LDL-C from the UKB cohort; the dot and vertical bar along the y-axis denotes the GWAS marginal effect size estimate and 95% CI for carotid_artery_dis from the 23andMe cohort. The colored lines represent the 2SMR (IVW, MR-Egger, weighted median) estimates of genetically instrumented LDL-C onto carotid_artery_dis.

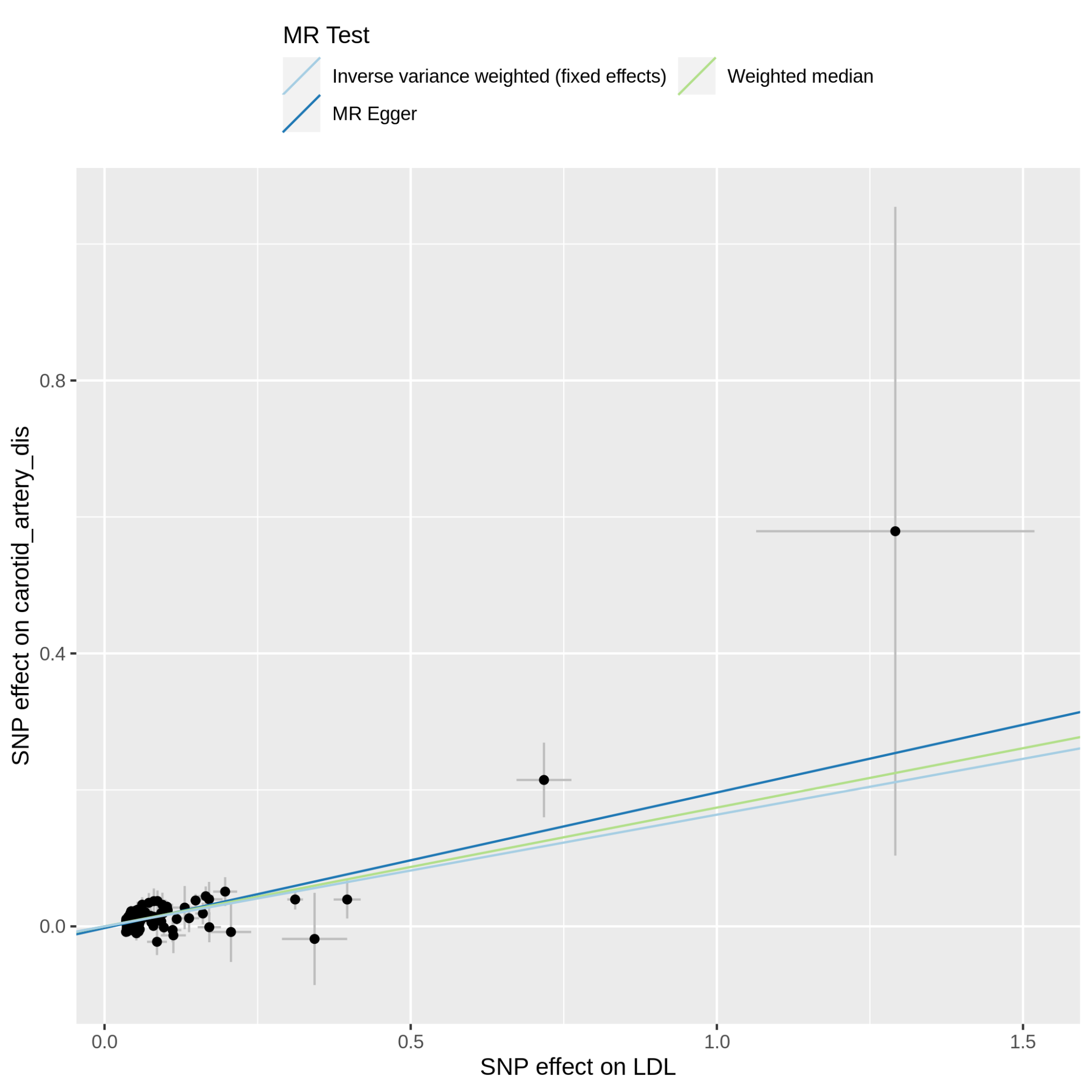

Supplementary Figure 11.5.2: MR scatter plot of LDL-C vs carotid_artery_dis, based on the reduced set of LDL-C genetic instruments (after heterogeneity and Steiger filtering). The dot and horizontal bar along the x-axis denotes the GWAS marginal effect size estimate and 95% CI for LDL-C from the UKB cohort; the dot and vertical bar along the y-axis denotes the GWAS marginal effect size estimate and 95% CI for carotid_artery_dis from the 23andMe cohort. The colored lines represent the 2SMR (IVW, MR-Egger, weighted median) estimates of genetically instrumented LDL-C onto carotid_artery_dis.

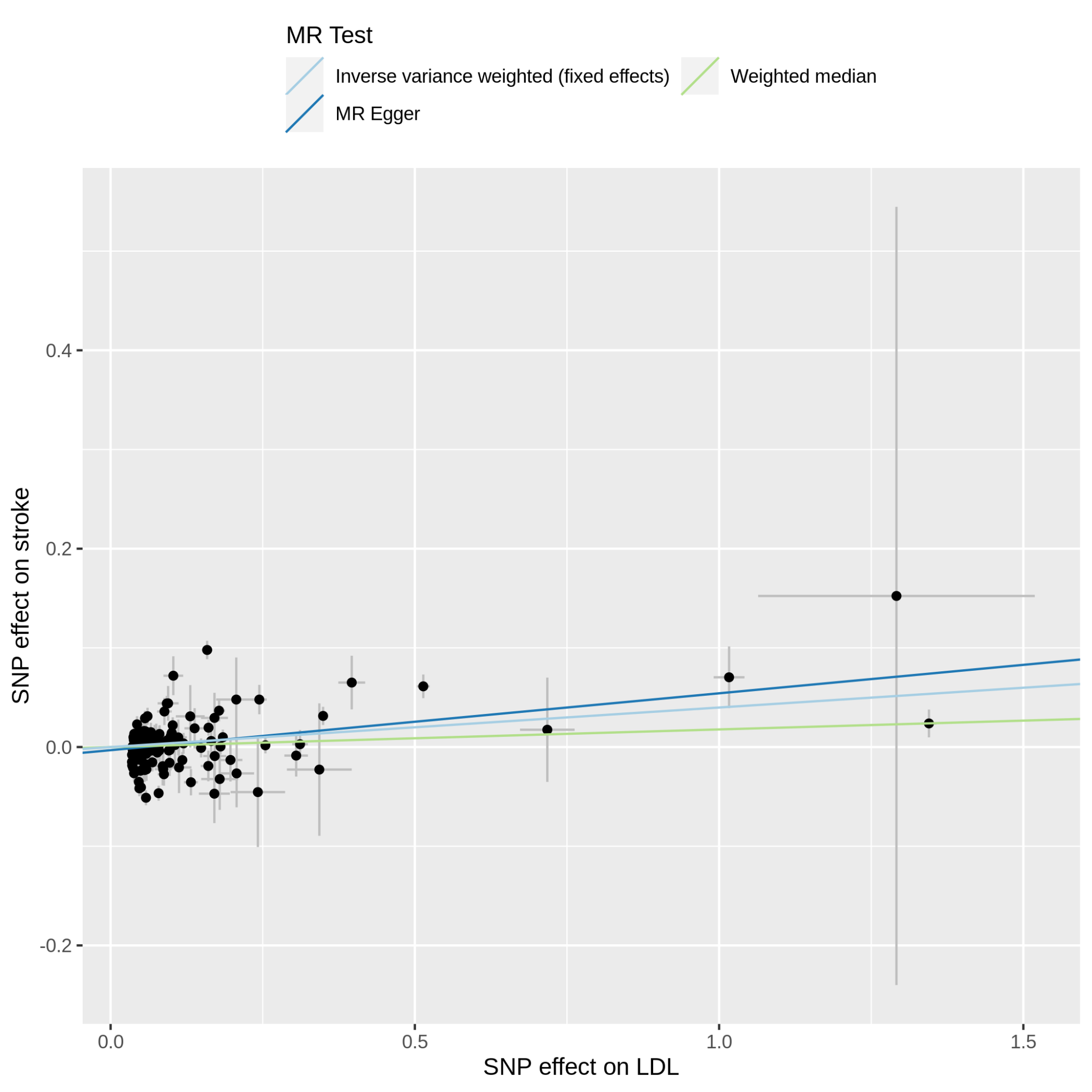

Supplementary Figure 11.6.1: MR scatter plot of LDL-C vs stroke, based on the full set of LDL-C genetic instruments. The dot and horizontal bar along the x-axis denotes the GWAS marginal effect size estimate and 95% CI for LDL-C from the UKB cohort; the dot and vertical bar along the y-axis denotes the GWAS marginal effect size estimate and 95% CI for stroke from the 23andMe cohort. The colored lines represent the 2SMR (IVW, MR-Egger, weighted median) estimates of genetically instrumented LDL-C onto stroke.

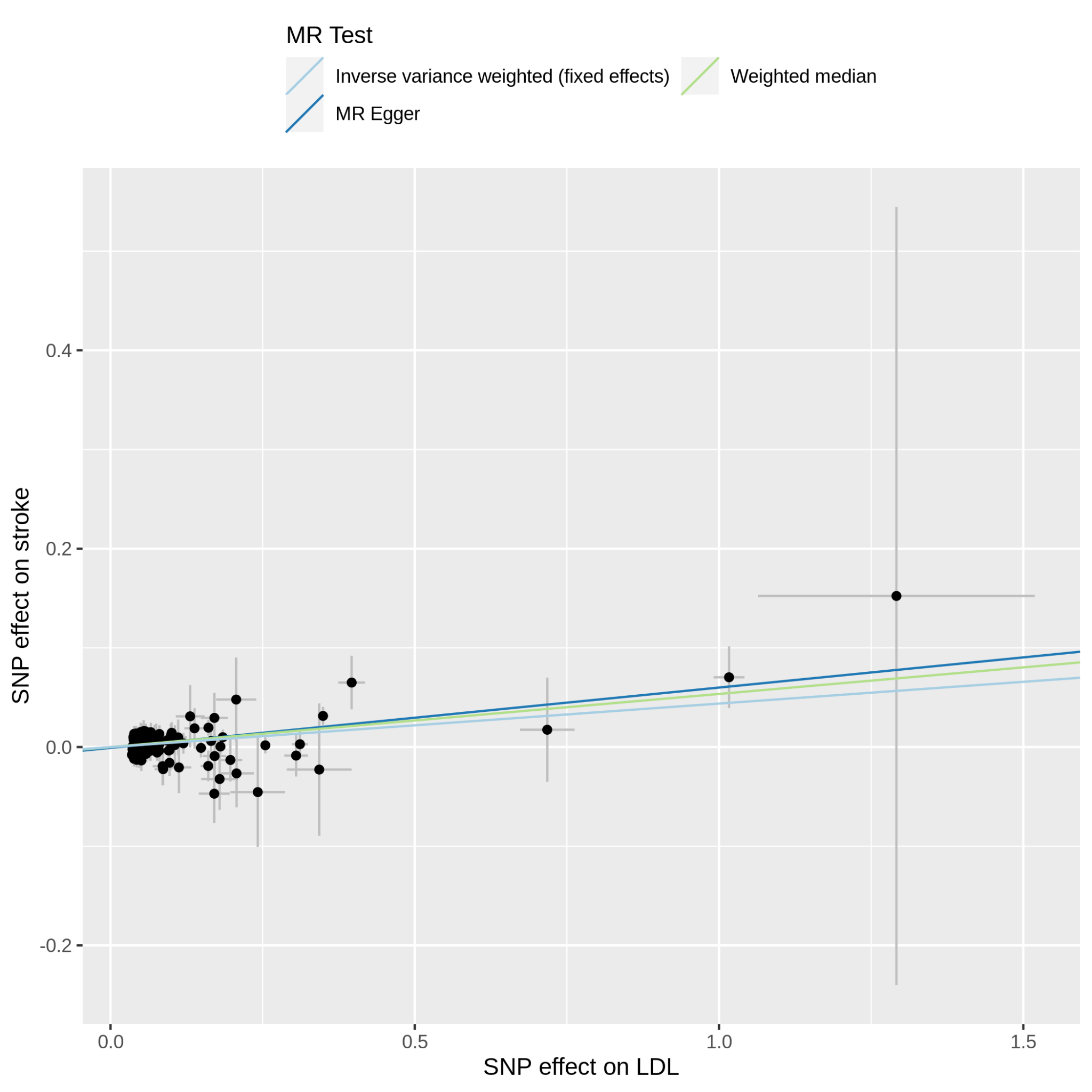

Supplementary Figure 11.6.2: MR scatter plot of LDL-C vs stroke, based on the reduced set of LDL-C genetic instruments (after heterogeneity and Steiger filtering). The dot and horizontal bar along the x-axis denotes the GWAS marginal effect size estimate and 95% CI for LDL-C from the UKB cohort; the dot and vertical bar along the y-axis denotes the GWAS marginal effect size estimate and 95% CI for stroke from the 23andMe cohort. The colored lines represent the 2SMR (IVW, MR-Egger, weighted median) estimates of genetically instrumented LDL-C onto stroke.

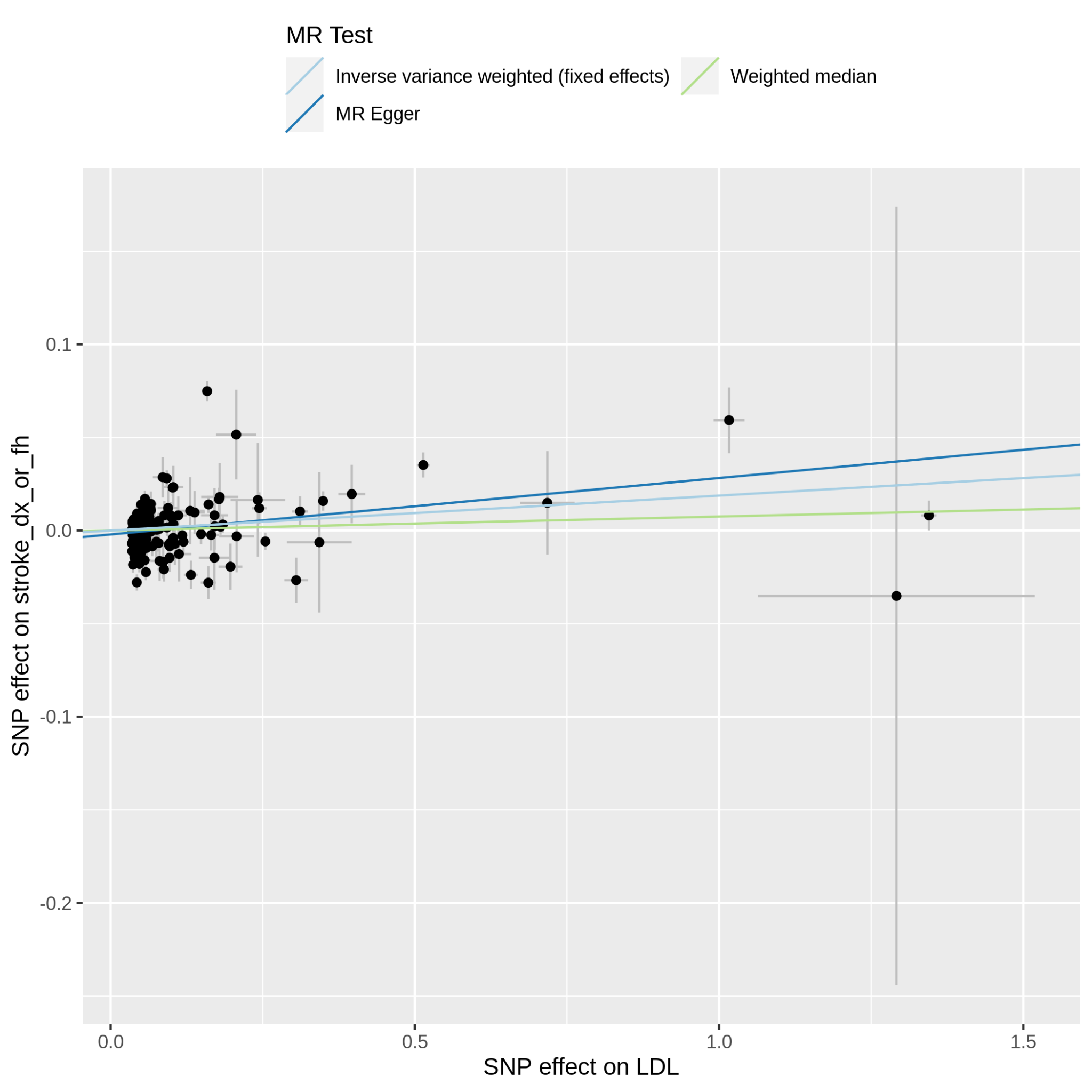

Supplementary Figure 11.7.1: MR scatter plot of LDL-C vs stroke_dx_or_fh, based on the full set of LDL-C genetic instruments. The dot and horizontal bar along the x-axis denotes the GWAS marginal effect size estimate and 95% CI for LDL-C from the UKB cohort; the dot and vertical bar along the y-axis denotes the GWAS marginal effect size estimate and 95% CI for stroke_dx_or_fh from the 23andMe cohort. The colored lines represent the 2SMR (IVW, MR-Egger, weighted median) estimates of genetically instrumented LDL-C onto stroke_dx_or_fh.

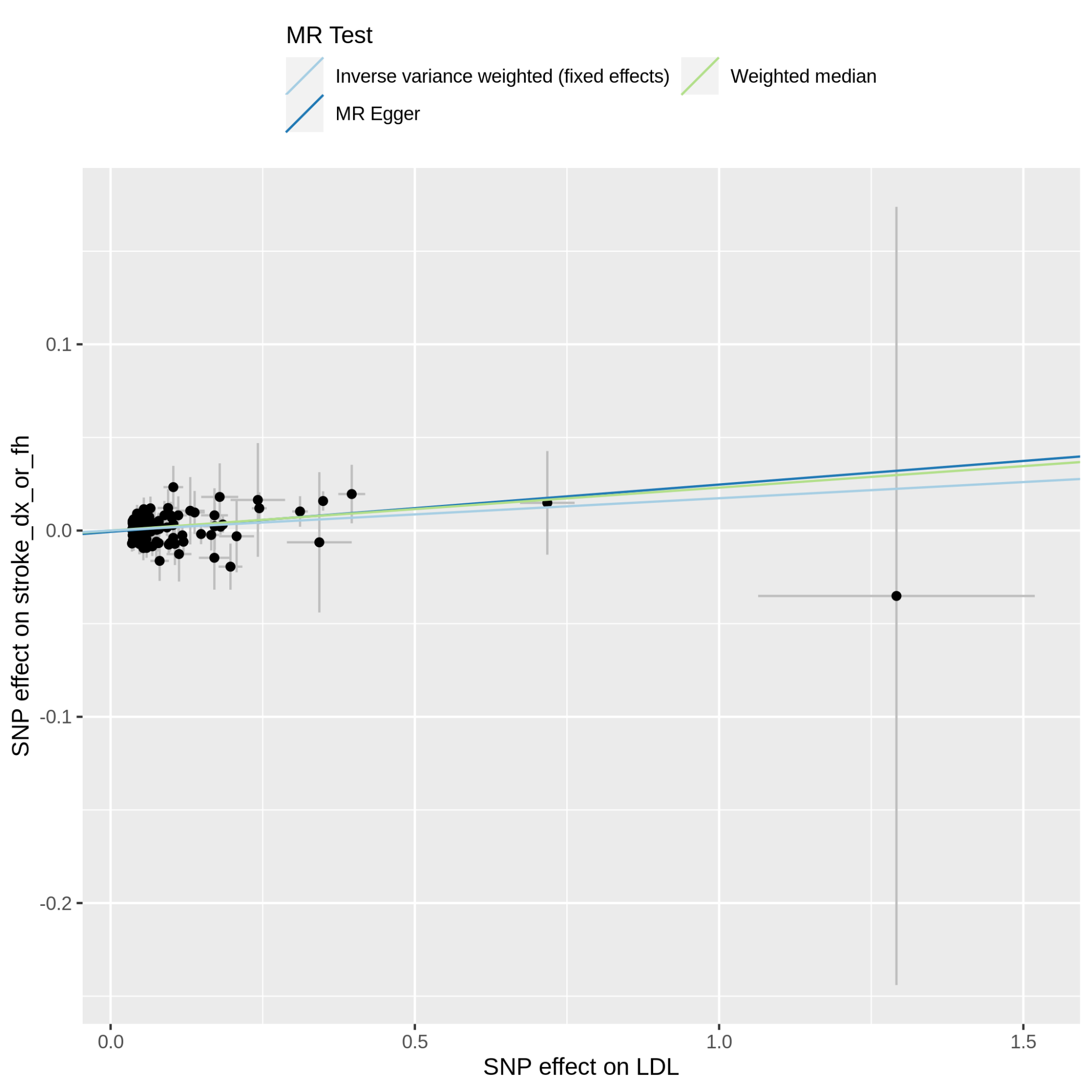

Supplementary Figure 11.7.2: MR scatter plot of LDL-C vs stroke_dx_or_fh, based on the reduced set of LDL-C genetic instruments (after heterogeneity and Steiger filtering). The dot and horizontal bar along the x-axis denotes the GWAS marginal effect size estimate and 95% CI for LDL-C from the UKB cohort; the dot and vertical bar along the y-axis denotes the GWAS marginal effect size estimate and 95% CI for stroke_dx_or_fh from the 23andMe cohort. The colored lines represent the 2SMR (IVW, MR-Egger, weighted median) estimates of genetically instrumented LDL-C onto stroke_dx_or_fh.

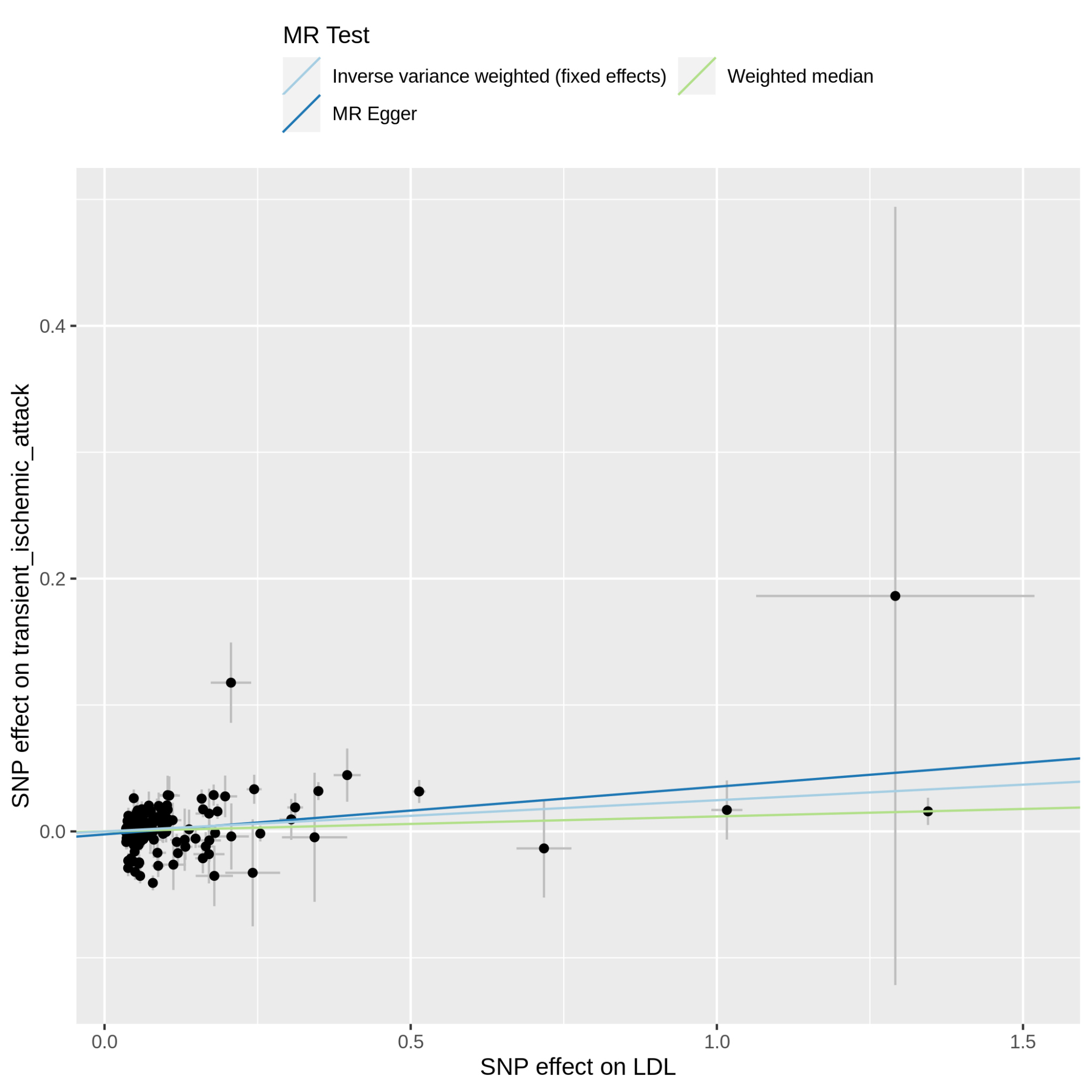

Supplementary Figure 11.8.1: MR scatter plot of LDL-C vs transient_ischemic_attack, based on the full set of LDL-C genetic instruments. The dot and horizontal bar along the x-axis denotes the GWAS marginal effect size estimate and 95% CI for LDL-C from the UKB cohort; the dot and vertical bar along the y-axis denotes the GWAS marginal effect size estimate and 95% CI for transient_ischemic_attack from the 23andMe cohort. The colored lines represent the 2SMR (IVW, MR-Egger, weighted median) estimates of genetically instrumented LDL-C onto transient_ischemic_attack.

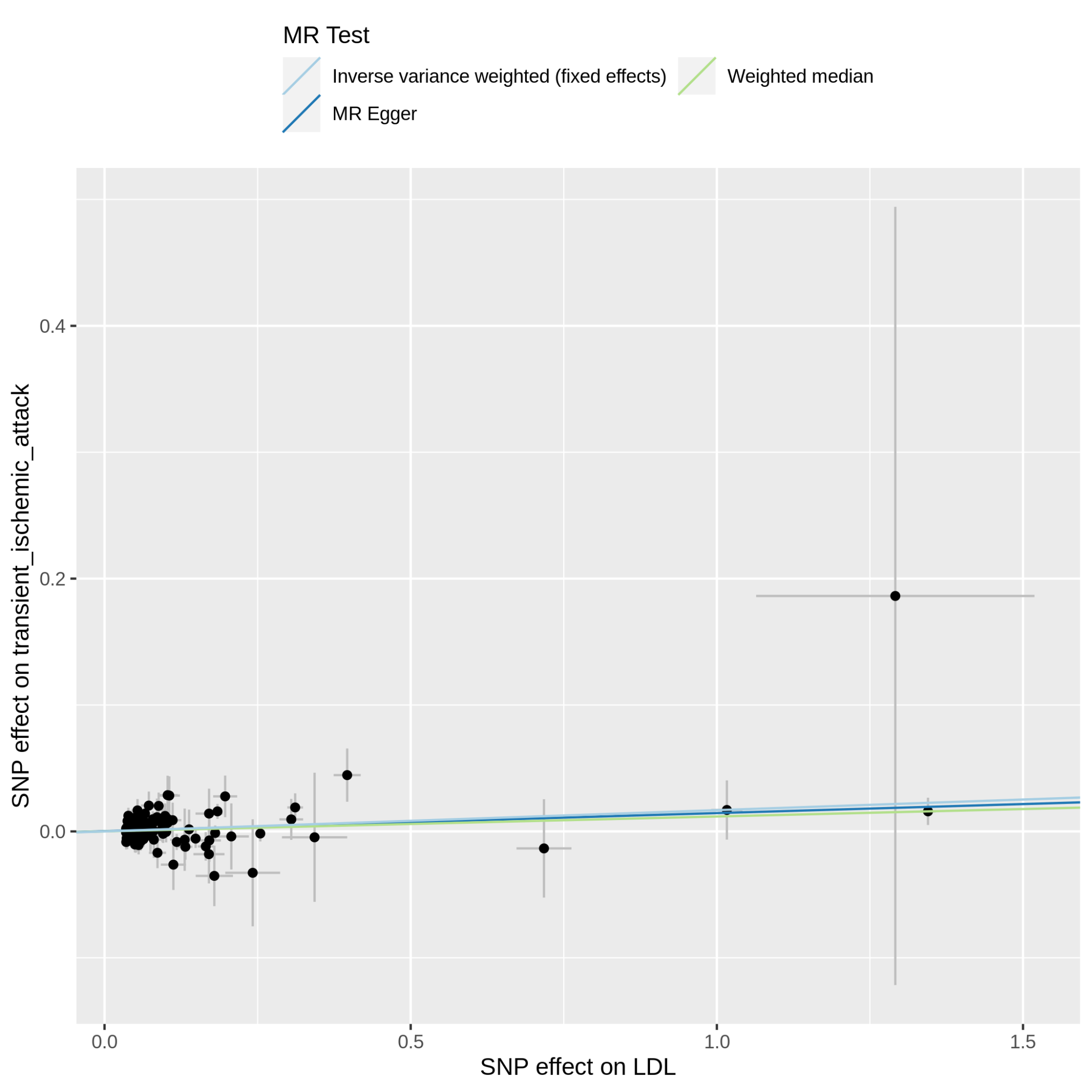

Supplementary Figure 11.8.2: MR scatter plot of LDL-C vs transient_ischemic_attack, based on the reduced set of LDL-C genetic instruments (after heterogeneity and Steiger filtering). The dot and horizontal bar along the x-axis denotes the GWAS marginal effect size estimate and 95% CI for LDL-C from the UKB cohort; the dot and vertical bar along the y-axis denotes the GWAS marginal effect size estimate and 95% CI for transient_ischemic_attack from the 23andMe cohort. The colored lines represent the 2SMR (IVW, MR-Egger, weighted median) estimates of genetically instrumented LDL-C onto transient_ischemic_attack.

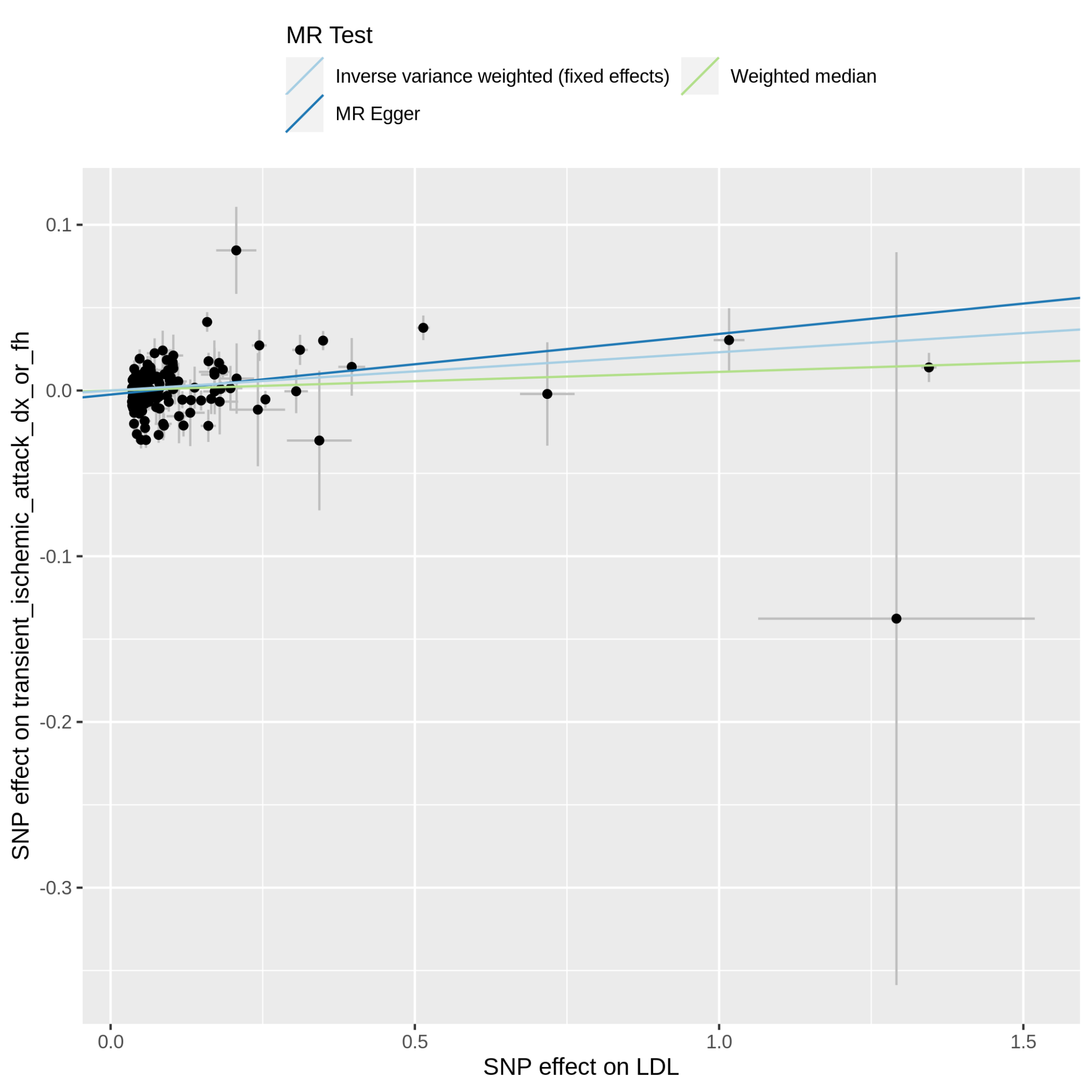

Supplementary Figure 11.9.1: MR scatter plot of LDL-C vs transient_ischemic_attack_dx_or_fh, based on the full set of LDL-C genetic instruments. The dot and horizontal bar along the x-axis denotes the GWAS marginal effect size estimate and 95% CI for LDL-C from the UKB cohort; the dot and vertical bar along the y-axis denotes the GWAS marginal effect size estimate and 95% CI for transient_ischemic_attack_dx_or_fh from the 23andMe cohort. The colored lines represent the 2SMR (IVW, MR-Egger, weighted median) estimates of genetically instrumented LDL onto transient_ischemic_attack_dx_or_fh.

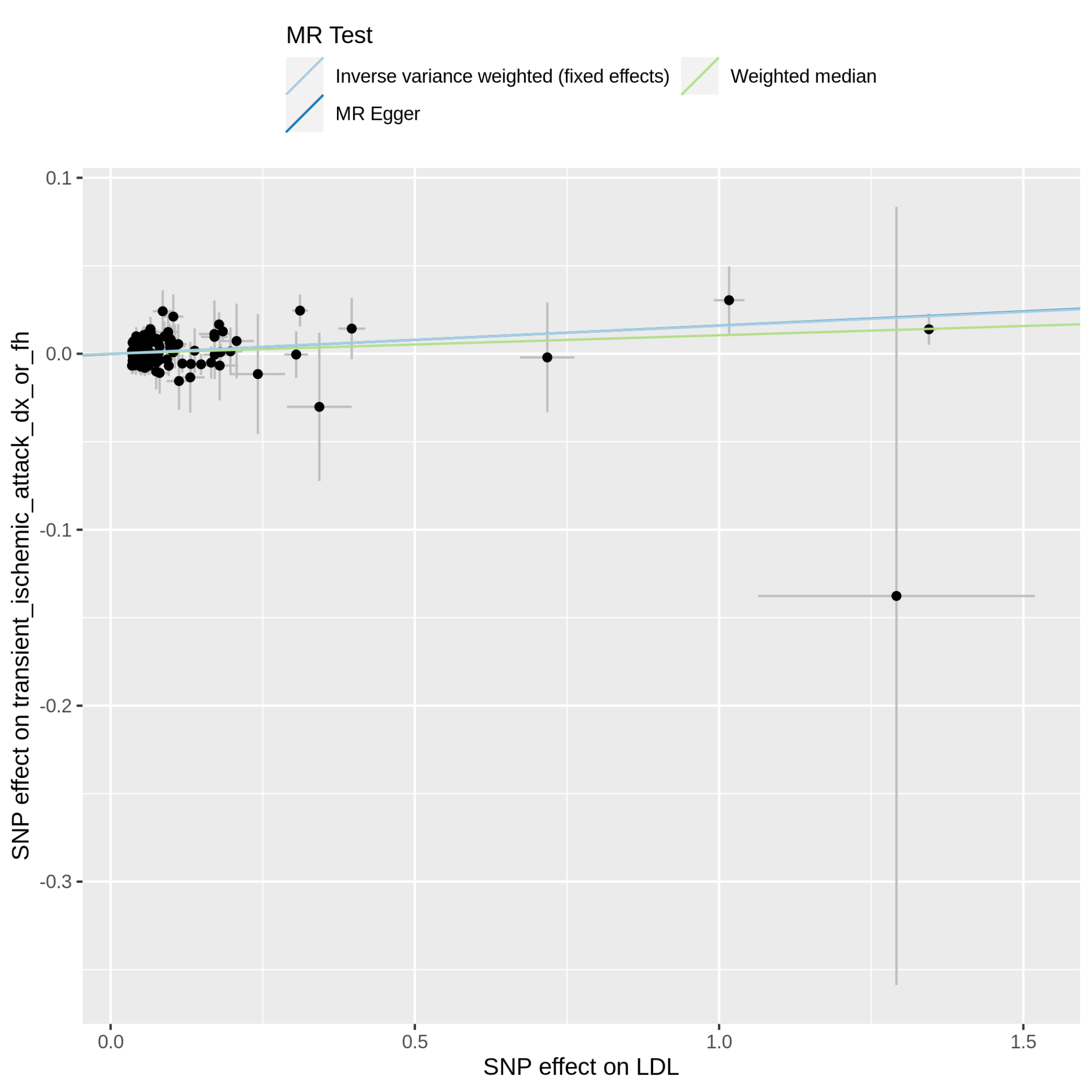

Supplementary Figure 11.9.2: MR scatter plot of LDL-C vs transient_ischemic_attack_dx_or_fh, based on the reduced set of LDL-C genetic instruments (after heterogeneity and Steiger filtering). The dot and horizontal bar along the x-axis denotes the GWAS marginal effect size estimate and 95% CI for LDL-C from the UKB cohort; the dot and vertical bar along the y-axis denotes the GWAS marginal effect size estimate and 95% CI for transient_ischemic_attack_dx_or_fh from the 23andMe cohort. The colored lines represent the 2SMR (IVW, MR-Egger, weighted median) estimates of genetically instrumented LDL-C onto transient_ischemic_attack_dx_or_fh.

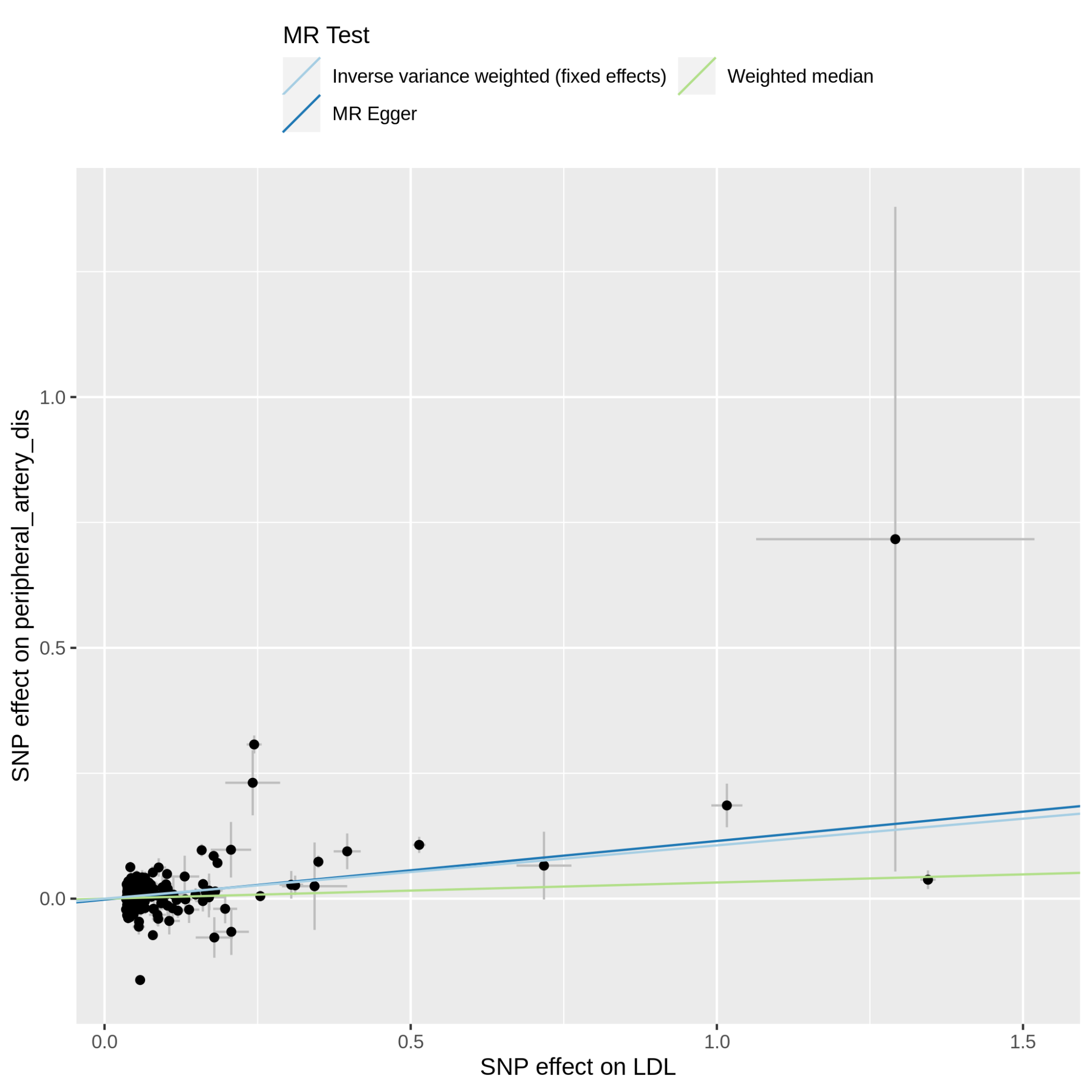

Supplementary Figure 11.10.1: MR scatter plot of LDL-C vs peripheral_artery_dis, based on the full set of LDL-C genetic instruments. The dot and horizontal bar along the x-axis denotes the GWAS marginal effect size estimate and 95% CI for LDL-C from the UKB cohort; the dot and vertical bar along the y-axis denotes the GWAS marginal effect size estimate and 95% CI for peripheral_artery_dis from the 23andMe cohort. The colored lines represent the 2SMR (IVW, MR-Egger, weighted median) estimates of genetically instrumented LDL-C onto peripheral_artery_dis.

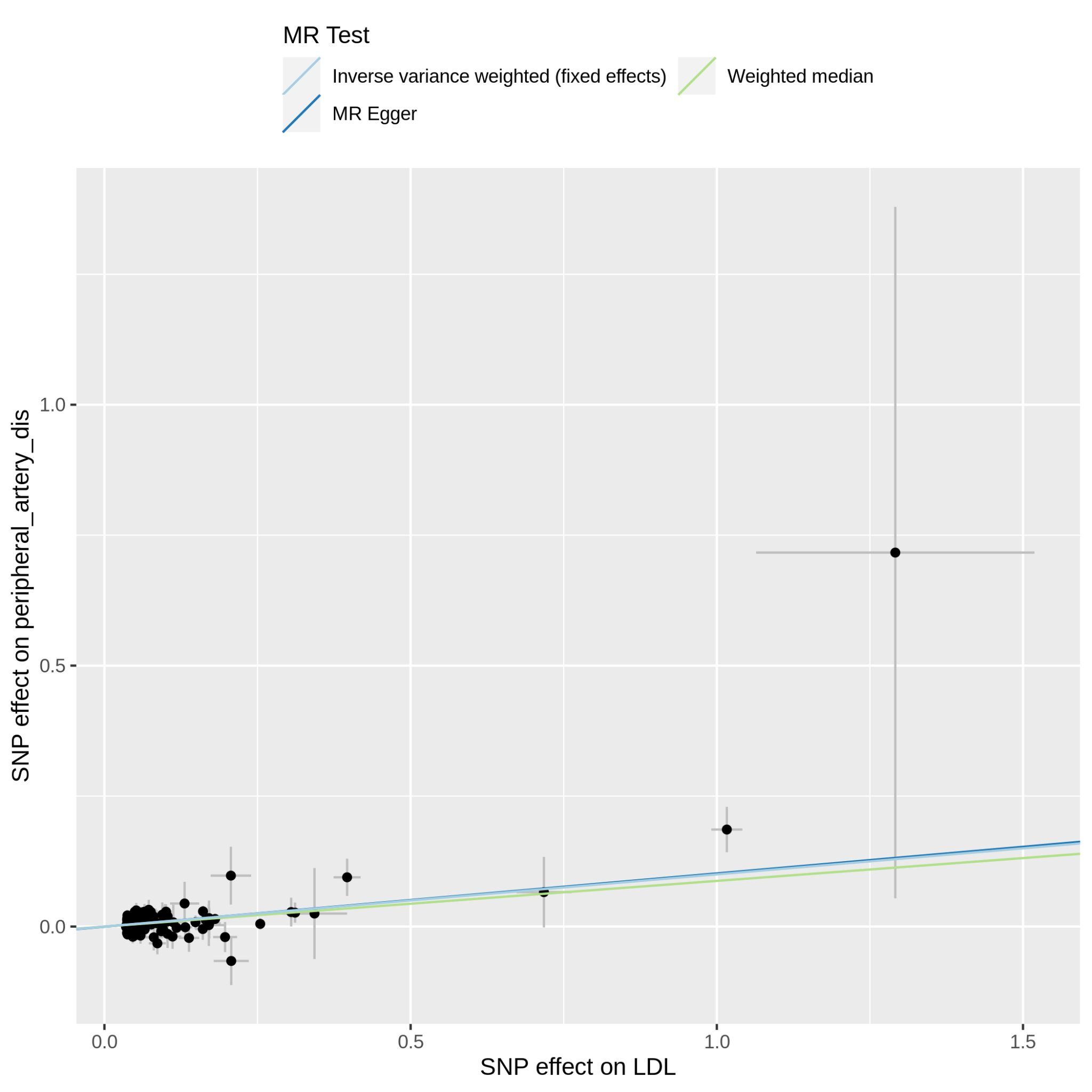

Supplementary Figure 11.10.2: MR scatter plot of LDL-C vs peripheral_artery_dis, based on the reduced set of LDL-C genetic instruments (after heterogeneity and Steiger filtering). The dot and horizontal bar along the x-axis denotes the GWAS marginal effect size estimate and 95% CI for LDL-C from the UKB cohort; the dot and vertical bar along the y-axis denotes the GWAS marginal effect size estimate and 95% CI for peripheral_artery_dis from the 23andMe cohort. The colored lines represent the 2SMR (IVW, MR-Egger, weighted median) estimates of genetically instrumented LDL-C onto peripheral_artery_dis.

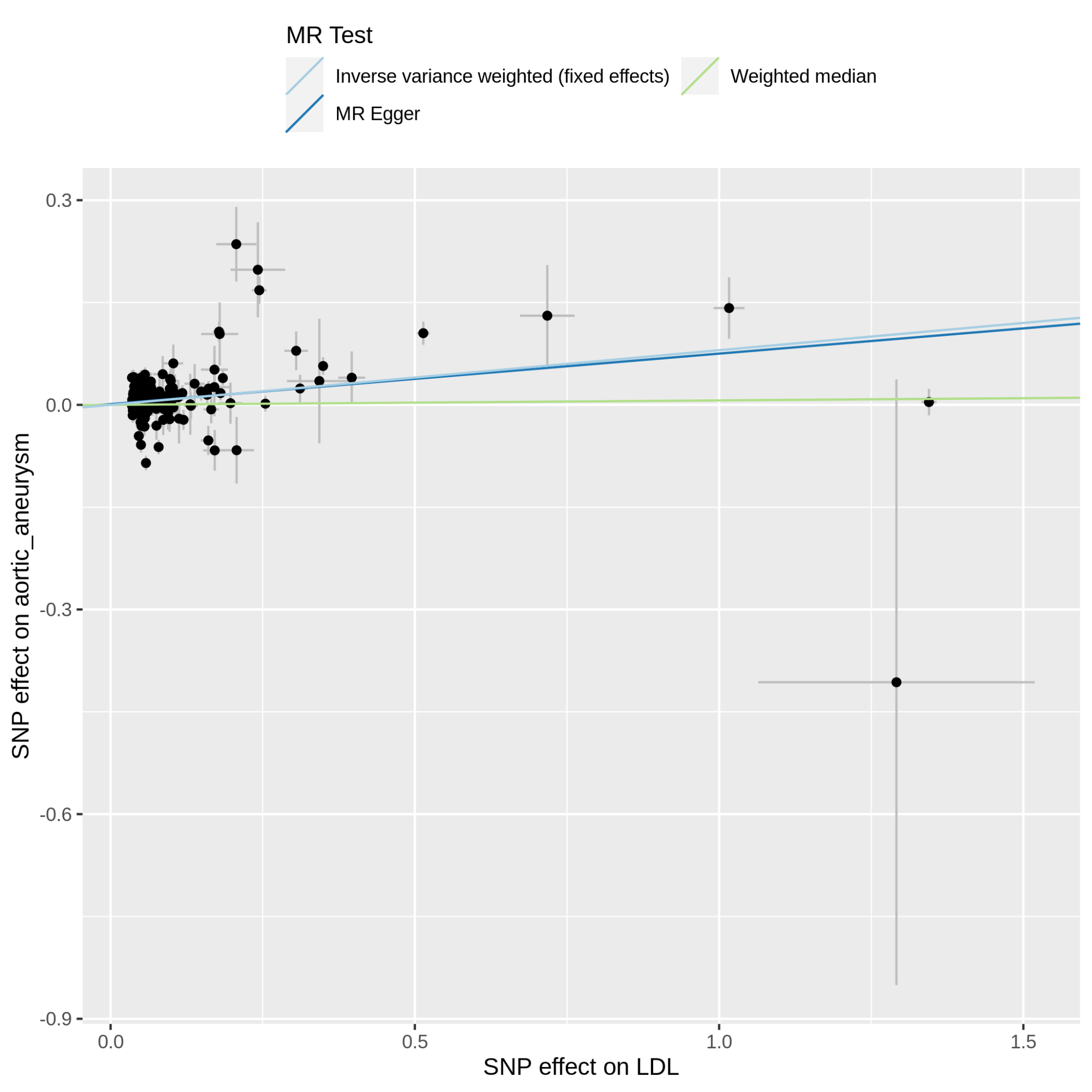

Supplementary Figure 11.11.1: MR scatter plot of LDL-C vs aortic_aneurysm, based on the full set of LDL-C genetic instruments. The dot and horizontal bar along the x-axis denotes the GWAS marginal effect size estimate and 95% CI for LDL-C from the UKB cohort; the dot and vertical bar along the y-axis denotes the GWAS marginal effect size estimate and 95% CI for aortic_aneurysm from the 23andMe cohort. The colored lines represent the 2SMR (IVW, MR-Egger, weighted median) estimates of genetically instrumented LDL-C onto aortic_aneurysm.

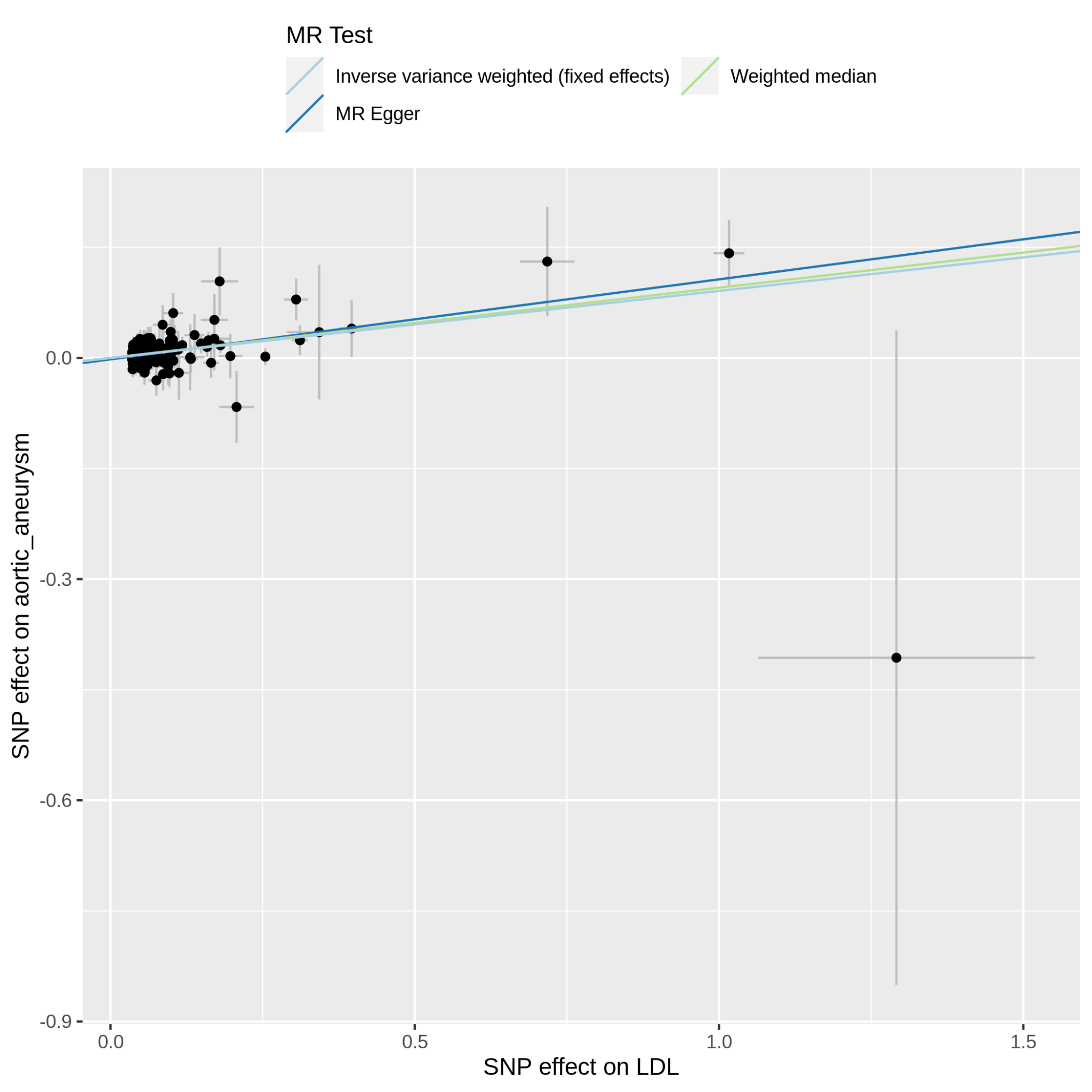

Supplementary Figure 11.11.2: MR scatter plot of LDL-C vs aortic_aneurysm, based on the reduced set of LDL-C genetic instruments (after heterogeneity and Steiger filtering). The dot and horizontal bar along the x-axis denotes the GWAS marginal effect size estimate and 95% CI for LDL-C from the UKB cohort; the dot and vertical bar along the y-axis denotes the GWAS marginal effect size estimate and 95% CI for aortic_aneurysm from the 23andMe cohort. The colored lines represent the 2SMR (IVW, MR-Egger, weighted median) estimates of genetically instrumented LDL-C onto aortic_aneurysm.

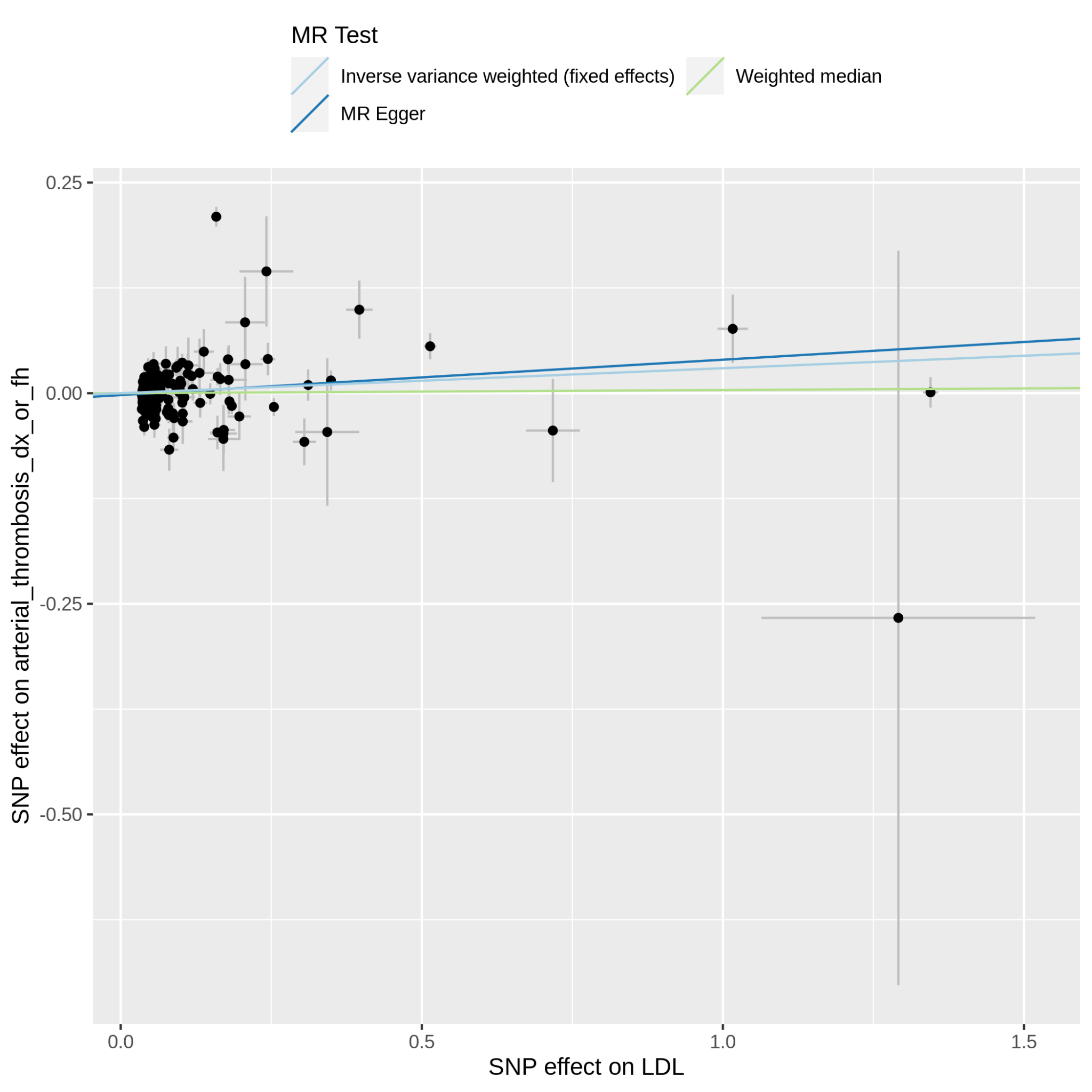

Supplementary Figure 11.12.1: MR scatter plot of LDL-C vs arterial_thrombosis_dx_or_fh, based on the full set of LDL-C genetic instruments. The dot and horizontal bar along the x-axis denotes the GWAS marginal effect size estimate and 95% CI for LDL-C from the UKB cohort; the dot and vertical bar along the y-axis denotes the GWAS marginal effect size estimate and 95% CI for arterial_thrombosis_dx_or_fh from the 23andMe cohort. The colored lines represent the 2SMR (IVW, MR-Egger, weighted median) estimates of genetically instrumented LDL-C onto arterial_thrombosis_dx_or_fh.

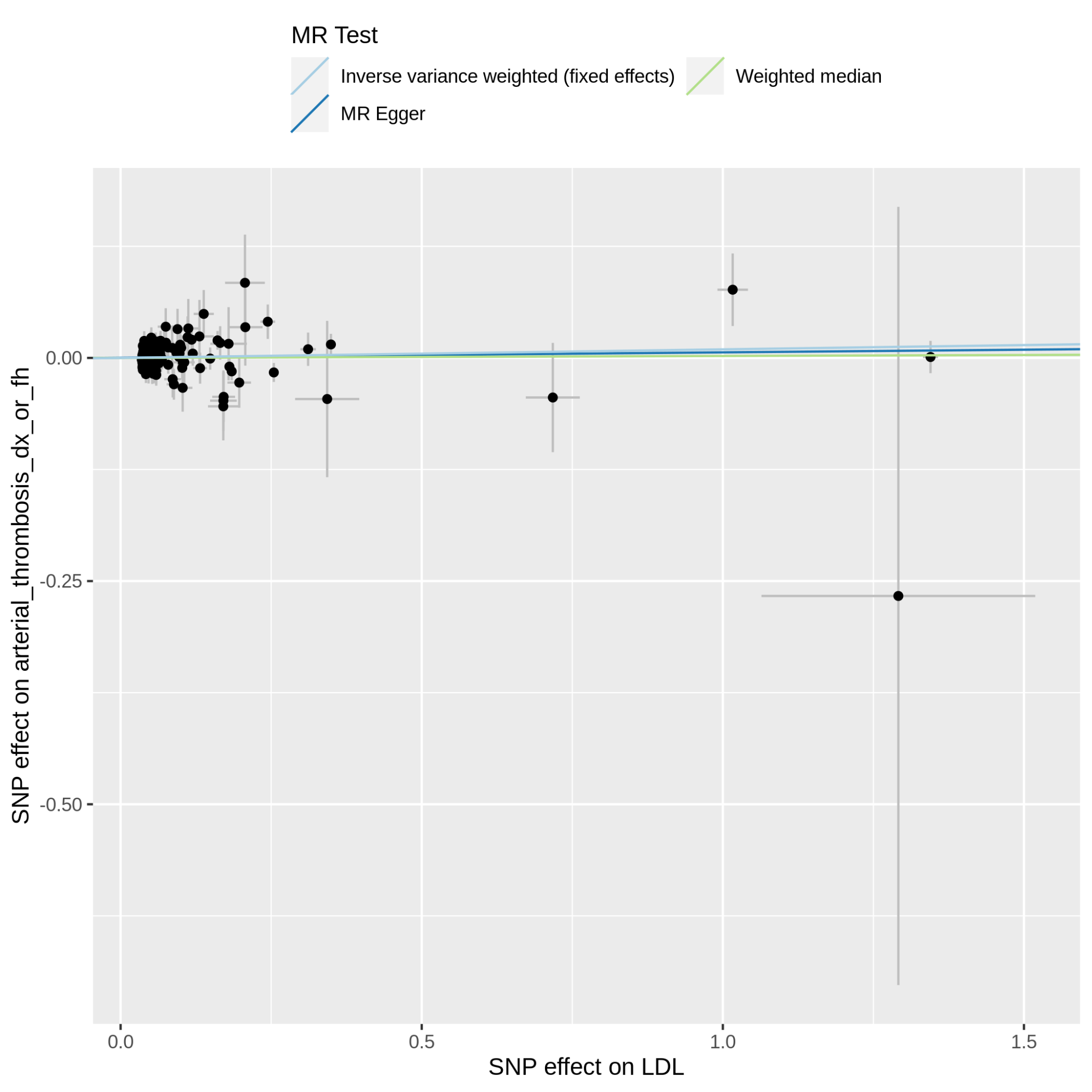

Supplementary Figure 11.12.2: MR scatter plot of LDL-C vs arterial_thrombosis_dx_or_fh, based on the reduced set of LDL-C genetic instruments (after heterogeneity and Steiger filtering). The dot and horizontal bar along the x-axis denotes the GWAS marginal effect size estimate and 95% CI for LDL-C from the UKB cohort; the dot and vertical bar along the y-axis denotes the GWAS marginal effect size estimate and 95% CI for arterial_thrombosis_dx_or_fh from the 23andMe cohort. The colored lines represent the 2SMR (IVW, MR-Egger, weighted median) estimates of genetically instrumented LDL-C onto arterial_thrombosis_dx_or_fh.

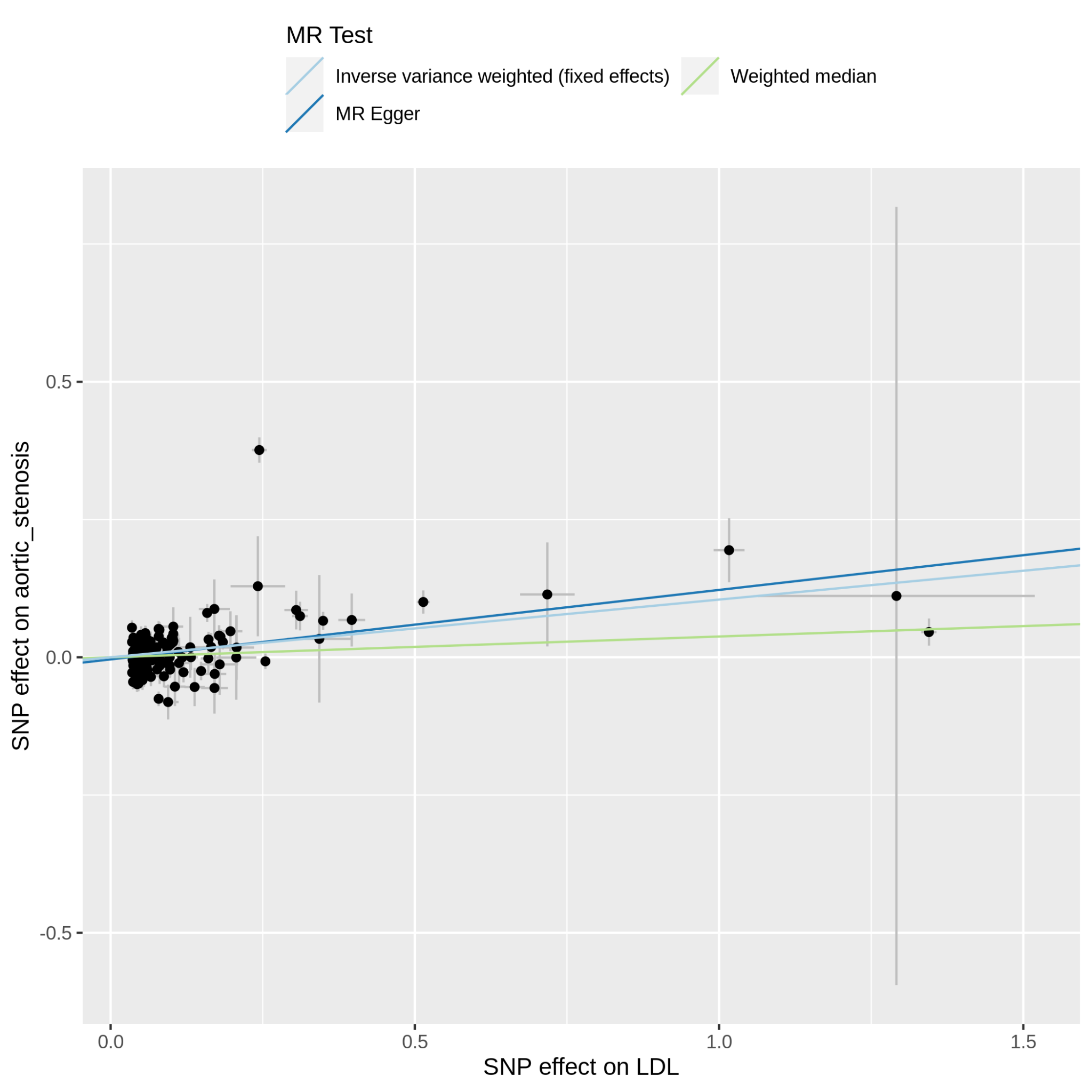

Supplementary Figure 11.13.1: MR scatter plot of LDL-C vs aortic_stenosis, based on the full set of LDL-C genetic instruments. The dot and horizontal bar along the x-axis denotes the GWAS marginal effect size estimate and 95% CI for LDL-C from the UKB cohort; the dot and vertical bar along the y-axis denotes the GWAS marginal effect size estimate and 95% CI for aortic_stenosis from the 23andMe cohort. The colored lines represent the 2SMR (IVW, MR-Egger, weighted median) estimates of genetically instrumented LDL-C onto aortic_stenosis.

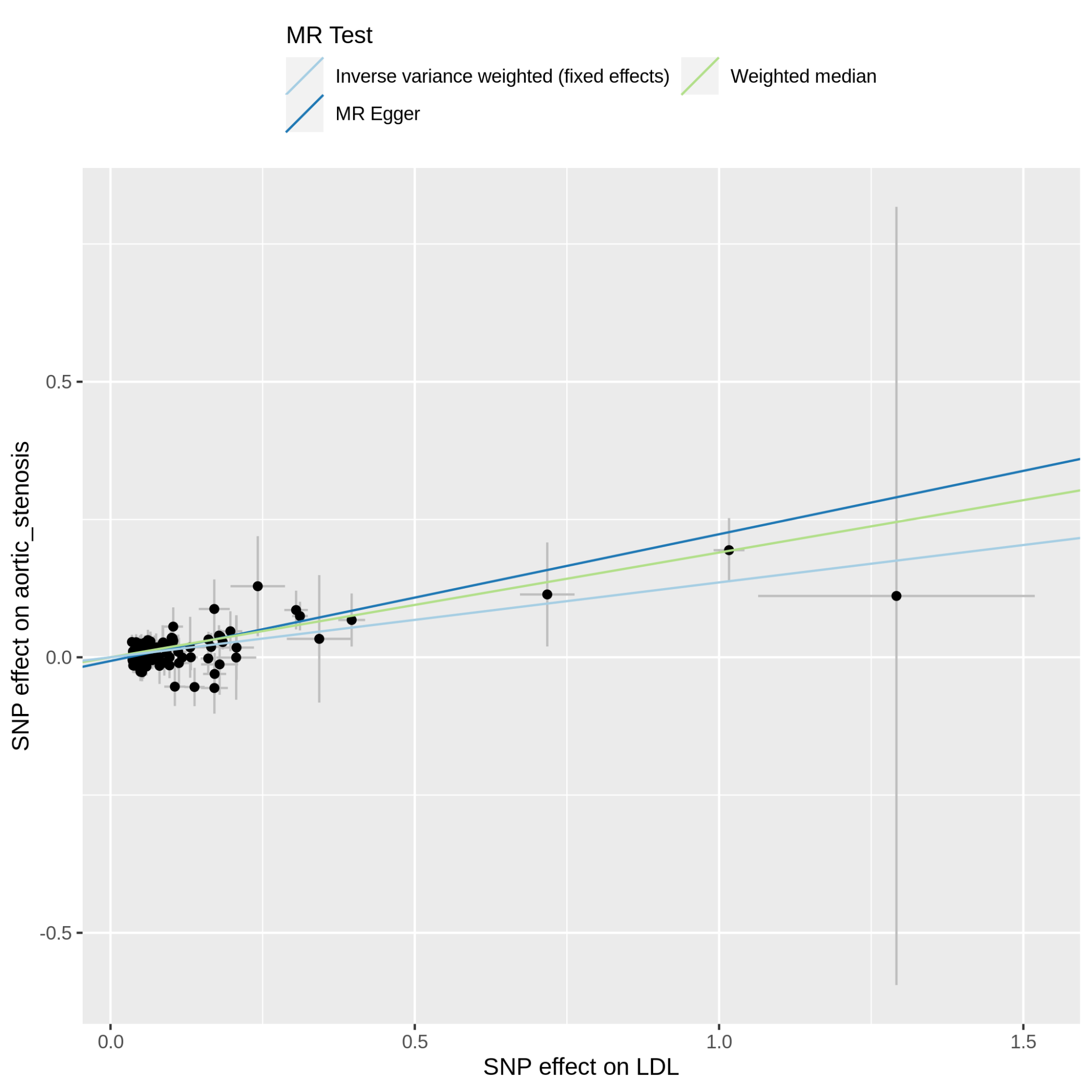

Supplementary Figure 11.13.2: MR scatter plot of LDL-C vs aortic_stenosis, based on the reduced set of LDL-C genetic instruments (after heterogeneity and Steiger filtering). The dot and horizontal bar along the x-axis denotes the GWAS marginal effect size estimate and 95% CI for LDL-C from the UKB cohort; the dot and vertical bar along the y-axis denotes the GWAS marginal effect size estimate and 95% CI for aortic_stenosis from the 23andMe cohort. The colored lines represent the 2SMR (IVW, MR-Egger, weighted median) estimates of genetically instrumented LDL-C onto aortic_stenosis.

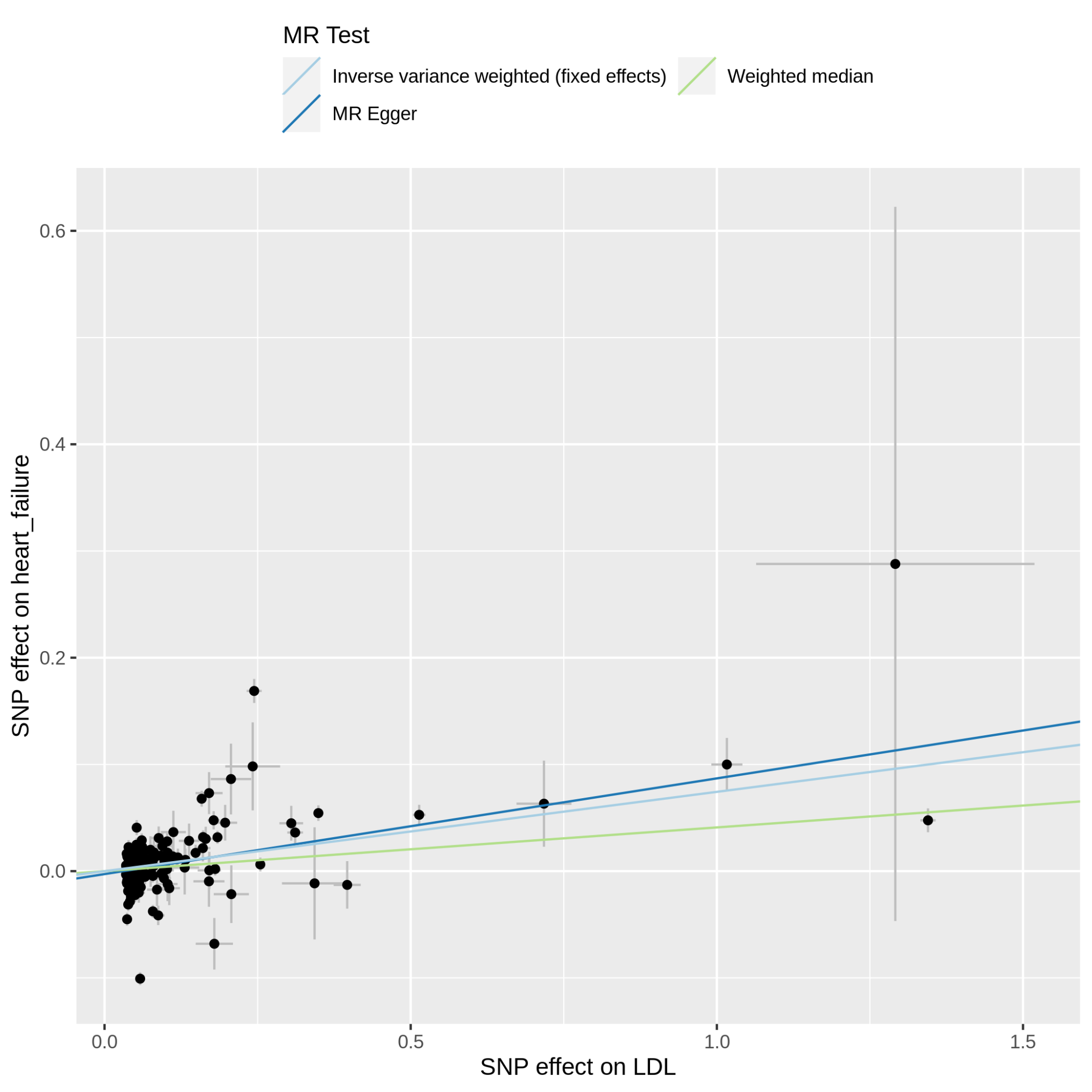

Supplementary Figure 11.14.1: MR scatter plot of LDL-C vs heart_failure, based on the full set of LDL-C genetic instruments. The dot and horizontal bar along the x-axis denotes the GWAS marginal effect size estimate and 95% CI for LDL-C from the UKB cohort; the dot and vertical bar along the y-axis denotes the GWAS marginal effect size estimate and 95% CI for heart_failure from the 23andMe cohort. The colored lines represent the 2SMR (IVW, MR-Egger, weighted median) estimates of genetically instrumented LDL-C onto heart_failure.

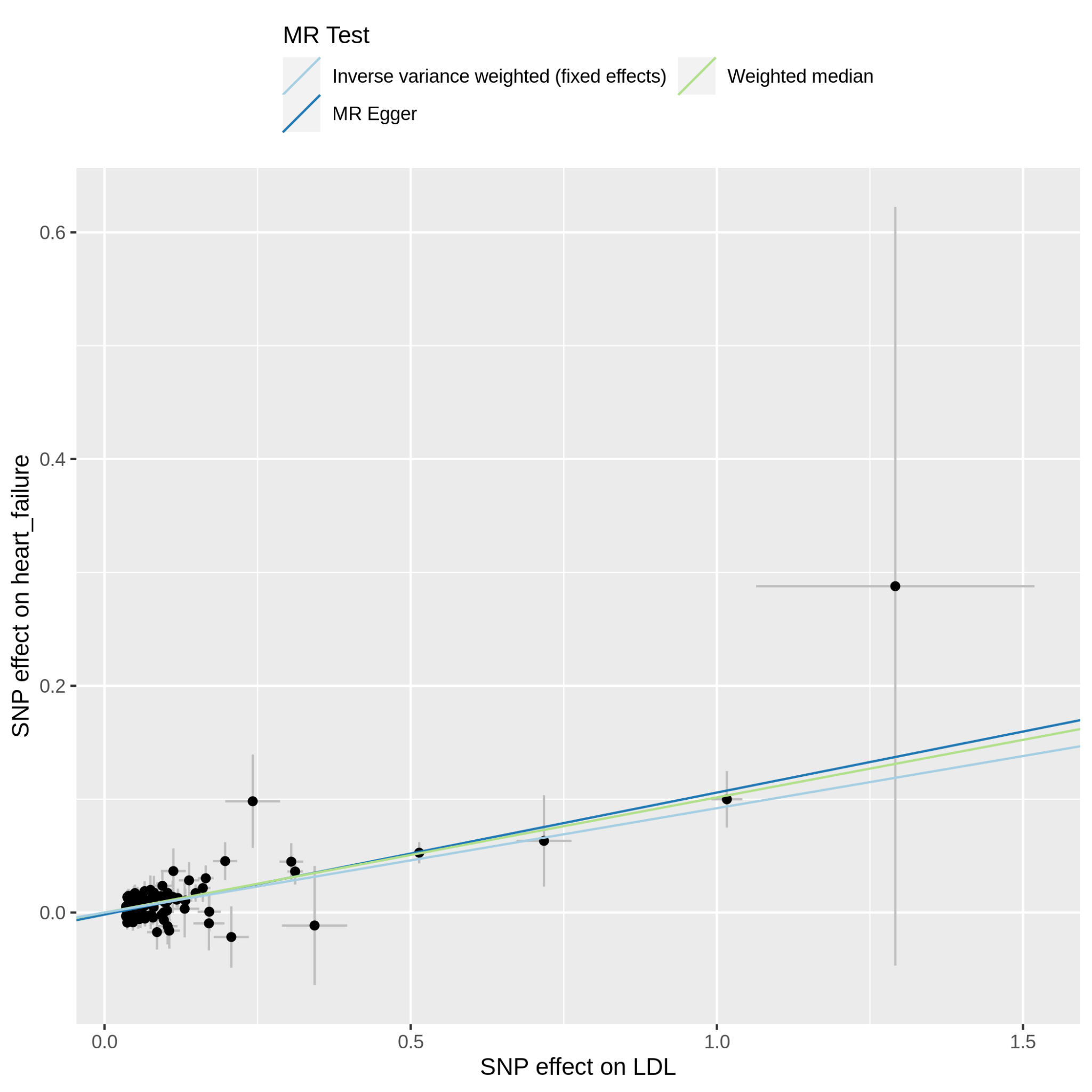

Supplementary Figure 11.14.2: MR scatter plot of LDL-C vs heart_failure, based on the reduced set of LDL-C genetic instruments (after heterogeneity and Steiger filtering). The dot and horizontal bar along the x-axis denotes the GWAS marginal effect size estimate and 95% CI for LDL-C from the UKB cohort; the dot and vertical bar along the y-axis denotes the GWAS marginal effect size estimate and 95% CI for heart_failure from the 23andMe cohort. The colored lines represent the 2SMR (IVW, MR-Egger, weighted median) estimates of genetically instrumented LDL-C onto heart_failure.

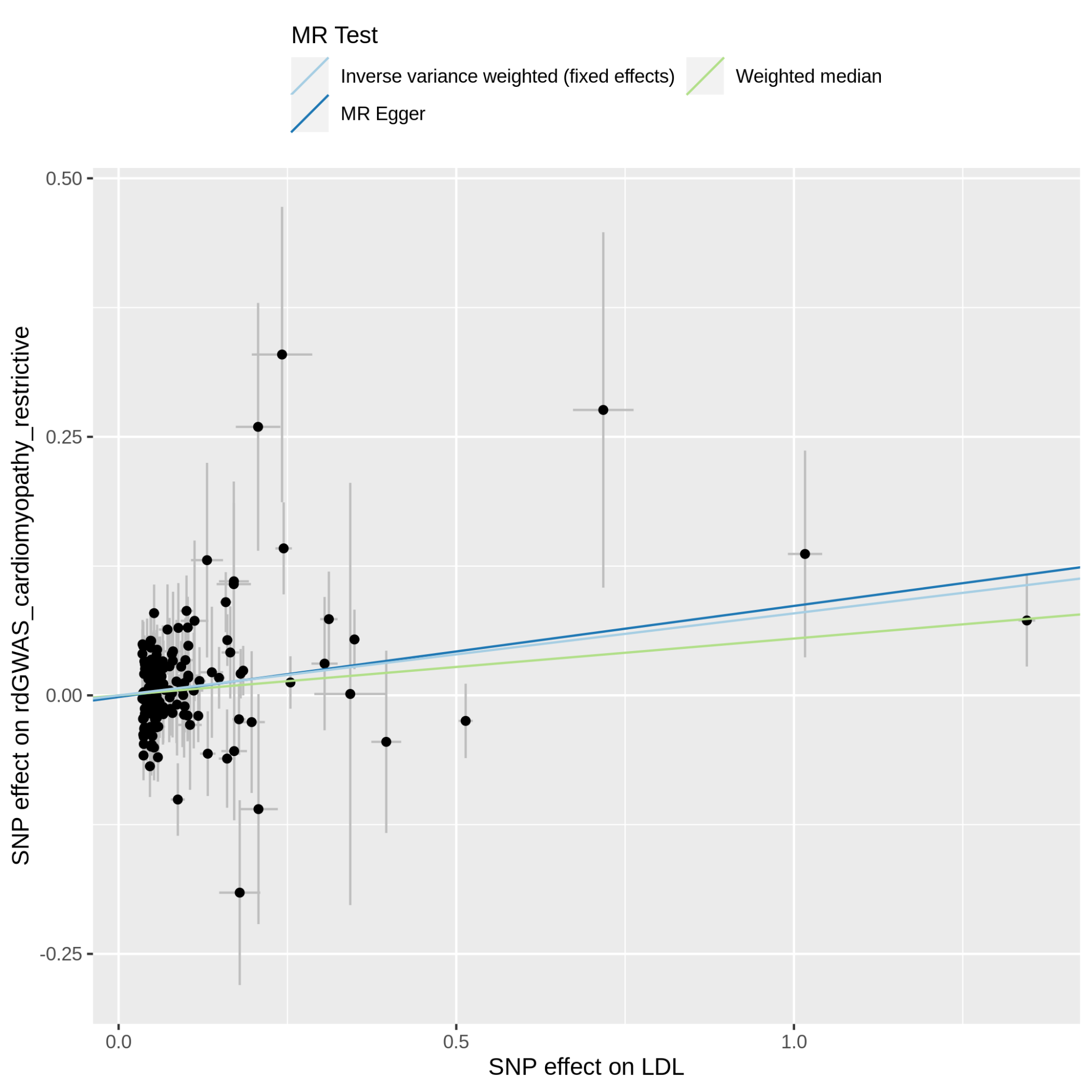

Supplementary Figure 11.15.1: MR scatter plot of LDL-C vs rdGWAS_cardiomyopathy_restrictive, based on the full set of LDL-C genetic instruments. The dot and horizontal bar along the x-axis denotes the GWAS marginal effect size estimate and 95% CI for LDL-C from the UKB cohort; the dot and vertical bar along the y-axis denotes the GWAS marginal effect size estimate and 95% CI for rdGWAS_cardiomyopathy_restrictive from the 23andMe cohort. The colored lines represent the 2SMR (IVW, MR-Egger, weighted median) estimates of genetically instrumented LDL onto rdGWAS_cardiomyopathy_restrictive.

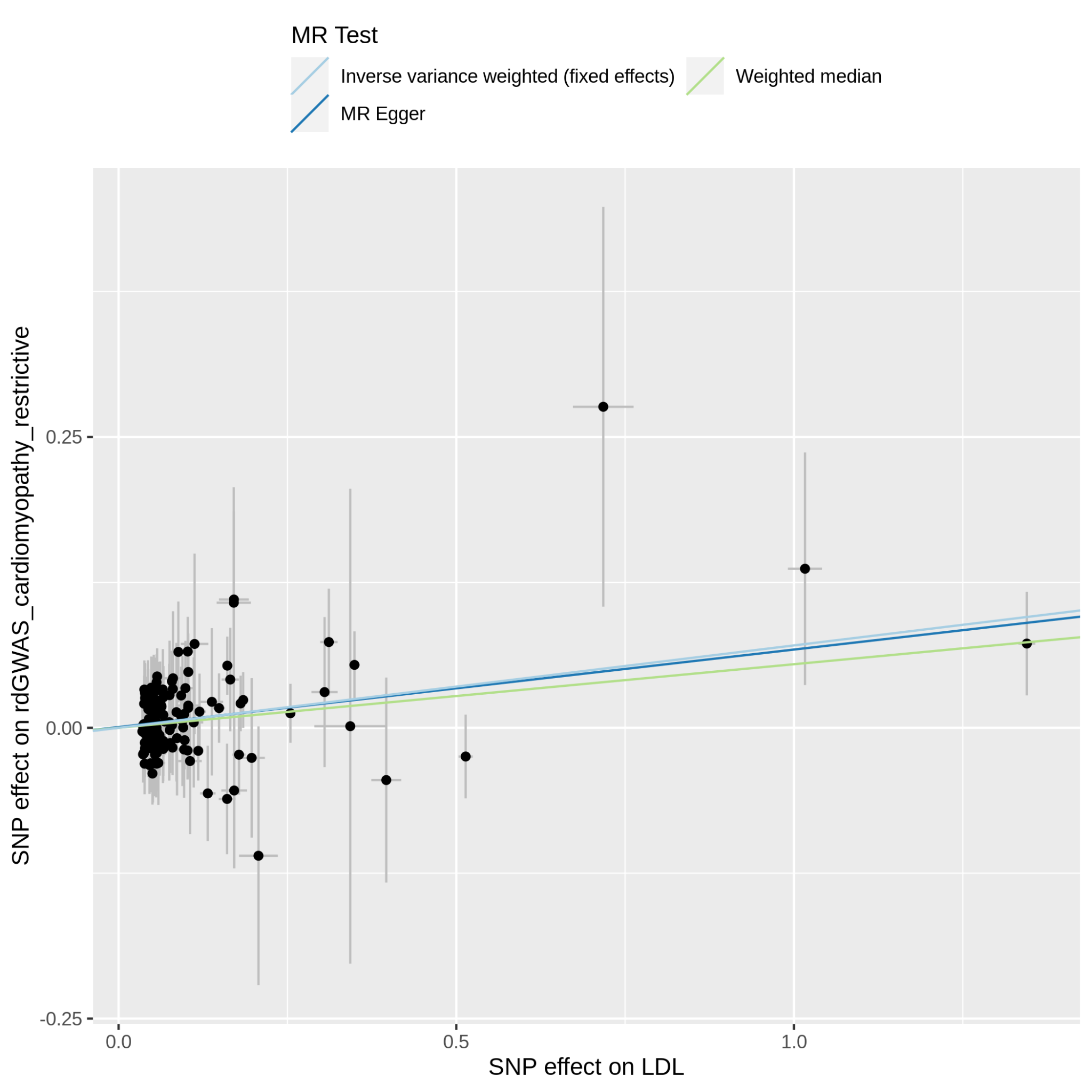

Supplementary Figure 11.15.2: MR scatter plot of LDL-C vs rdGWAS_cardiomyopathy_restrictive, based on the reduced set of LDL-C genetic instruments (after heterogeneity and Steiger filtering). The dot and horizontal bar along the x-axis denotes the GWAS marginal effect size estimate and 95% CI for LDL-C from the UKB cohort; the dot and vertical bar along the y-axis denotes the GWAS marginal effect size estimate and 95% CI for rdGWAS_cardiomyopathy_restrictive from the 23andMe cohort. The colored lines represent the 2SMR (IVW, MR-Egger, weighted median) estimates of genetically instrumented LDL-C onto rdGWAS_cardiomyopathy_restrictive.

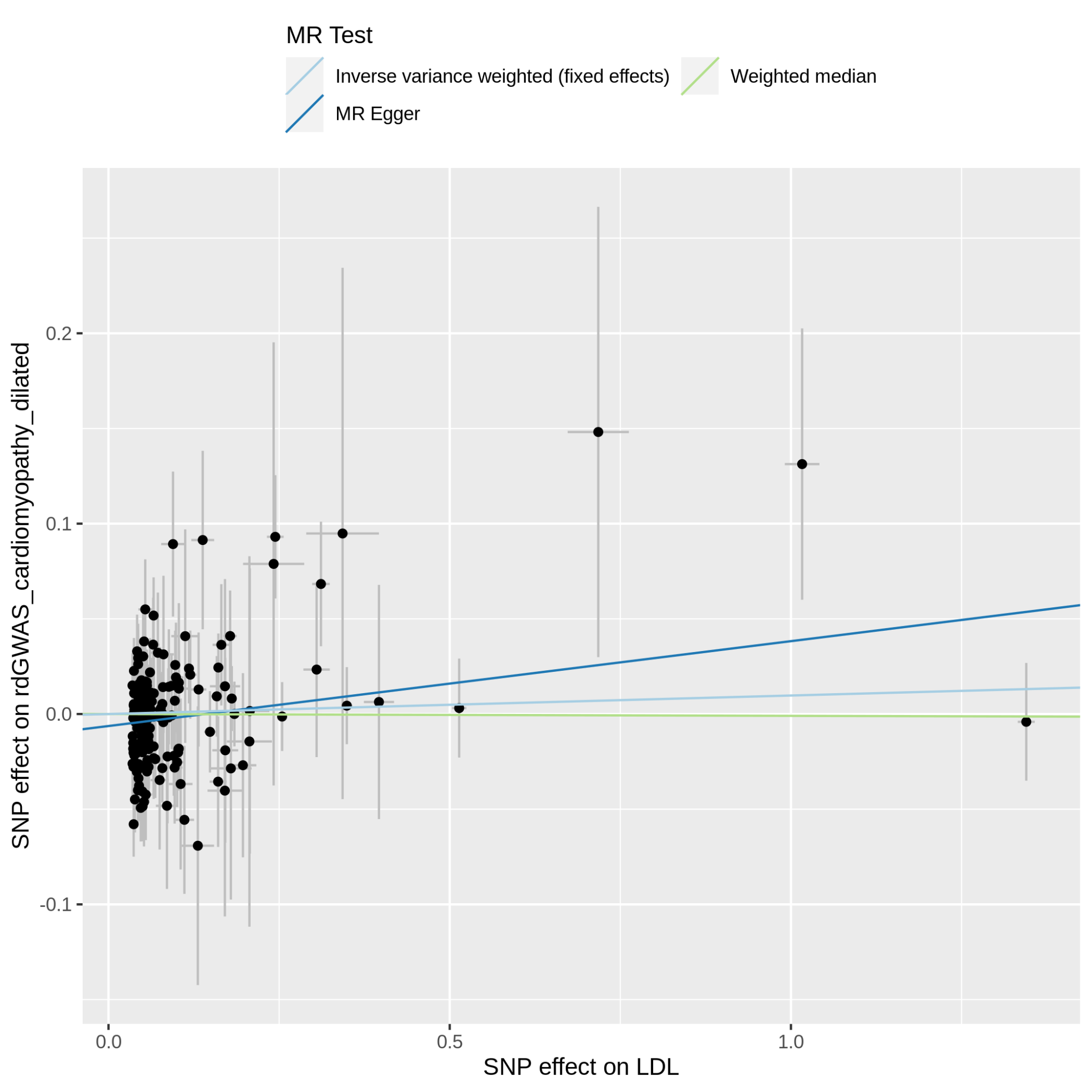

Supplementary Figure 11.16.1: MR scatter plot of LDL-C vs rdGWAS_cardiomyopathy_dilated, based on the full set of LDL-C genetic instruments. The dot and horizontal bar along the x-axis denotes the GWAS marginal effect size estimate and 95% CI for LDL-C from the UKB cohort; the dot and vertical bar along the y-axis denotes the GWAS marginal effect size estimate and 95% CI for rdGWAS_cardiomyopathy_dilated from the 23andMe cohort. The colored lines represent the 2SMR (IVW, MR-Egger, weighted median) estimates of genetically instrumented LDL-C onto rdGWAS_cardiomyopathy_dilated.

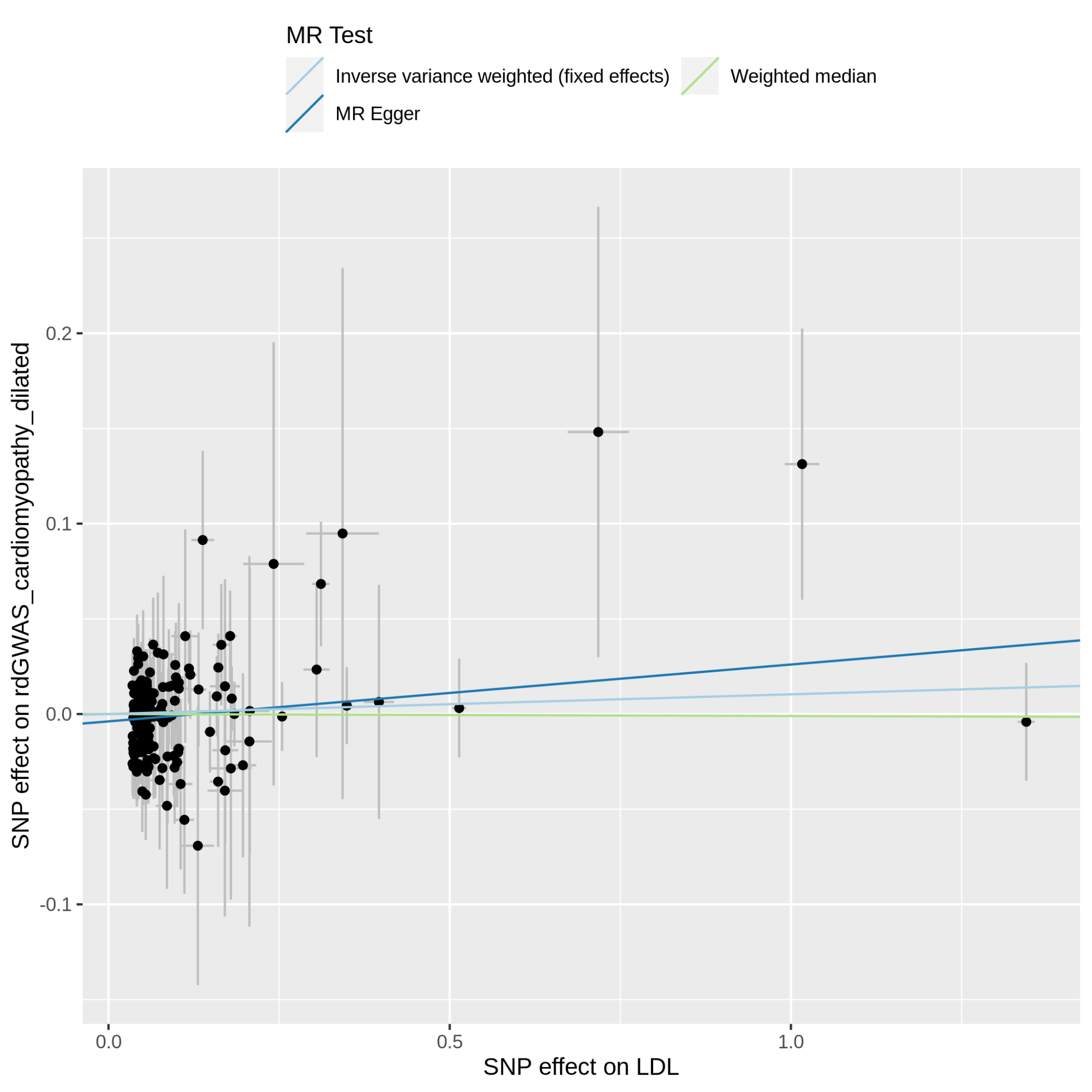

Supplementary Figure 11.16.2: MR scatter plot of LDL-C vs rdGWAS_cardiomyopathy_dilated, based on the reduced set of LDL-C genetic instruments (after heterogeneity and Steiger filtering). The dot and horizontal bar along the x-axis denotes the GWAS marginal effect size estimate and 95% CI for LDL-C from the UKB cohort; the dot and vertical bar along the y-axis denotes the GWAS marginal effect size estimate and 95% CI for rdGWAS_cardiomyopathy_dilated from the 23andMe cohort. The colored lines represent the 2SMR (IVW, MR-Egger, weighted median) estimates of genetically instrumented LDL-C onto rdGWAS_cardiomyopathy_dilated.

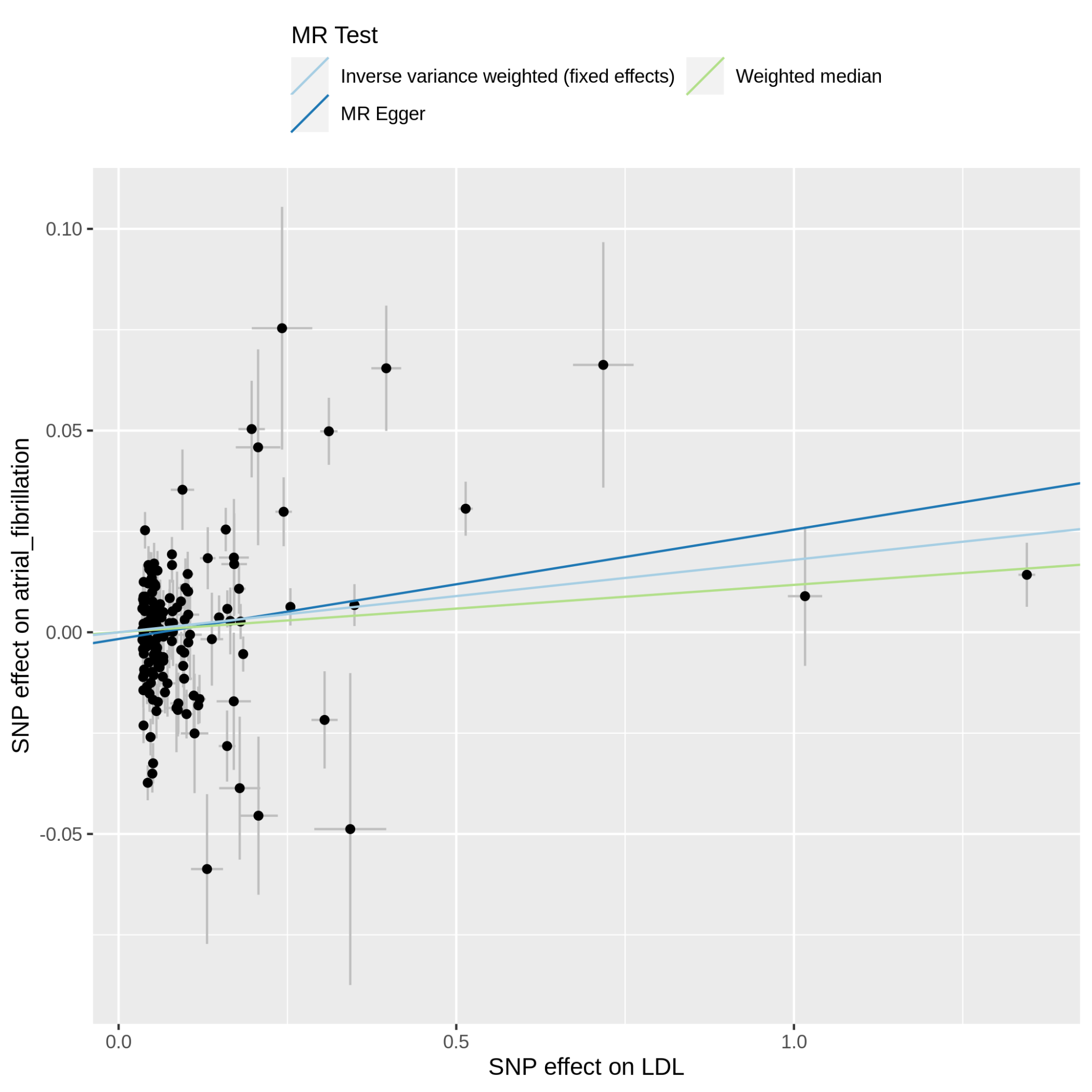

Supplementary Figure 11.17.1: MR scatter plot of LDL-C vs atrial_fibrillation, based on the full set of LDL-C genetic instruments. The dot and horizontal bar along the x-axis denotes the GWAS marginal effect size estimate and 95% CI for LDL-C from the UKB cohort; the dot and vertical bar along the y-axis denotes the GWAS marginal effect size estimate and 95% CI for atrial_fibrillation from the 23andMe cohort. The colored lines represent the 2SMR (IVW, MR-Egger, weighted median) estimates of genetically instrumented LDL-C onto atrial_fibrillation.

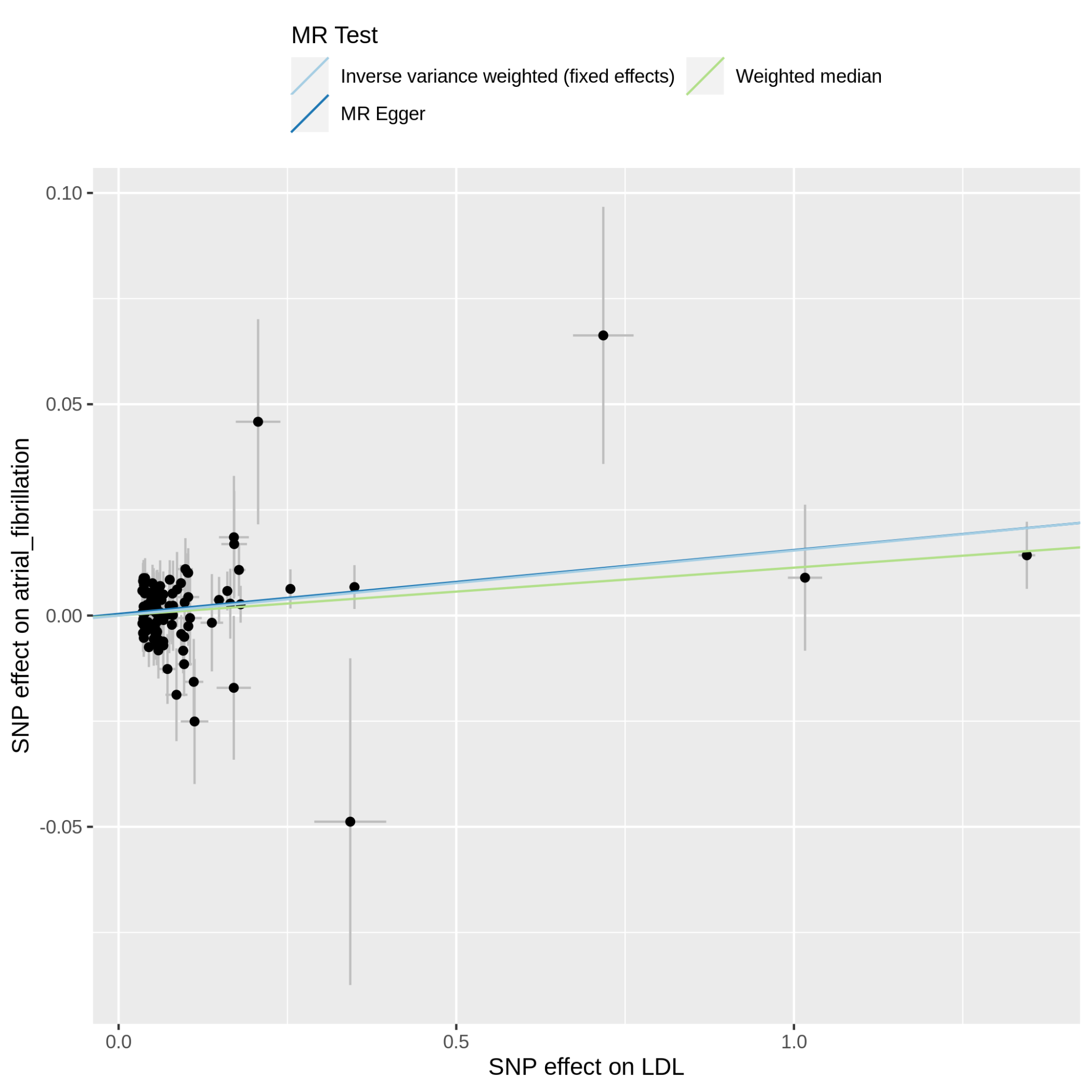

Supplementary Figure 11.17.2: MR scatter plot of LDL-C vs atrial_fibrillation, based on the reduced set of LDL-C genetic instruments (after heterogeneity and Steiger filtering). The dot and horizontal bar along the x-axis denotes the GWAS marginal effect size estimate and 95% CI for LDL-C from the UKB cohort; the dot and vertical bar along the y-axis denotes the GWAS marginal effect size estimate and 95% CI for atrial_fibrillation from the 23andMe cohort. The colored lines represent the 2SMR (IVW, MR-Egger, weighted median) estimates of genetically instrumented LDL-C onto atrial_fibrillation.

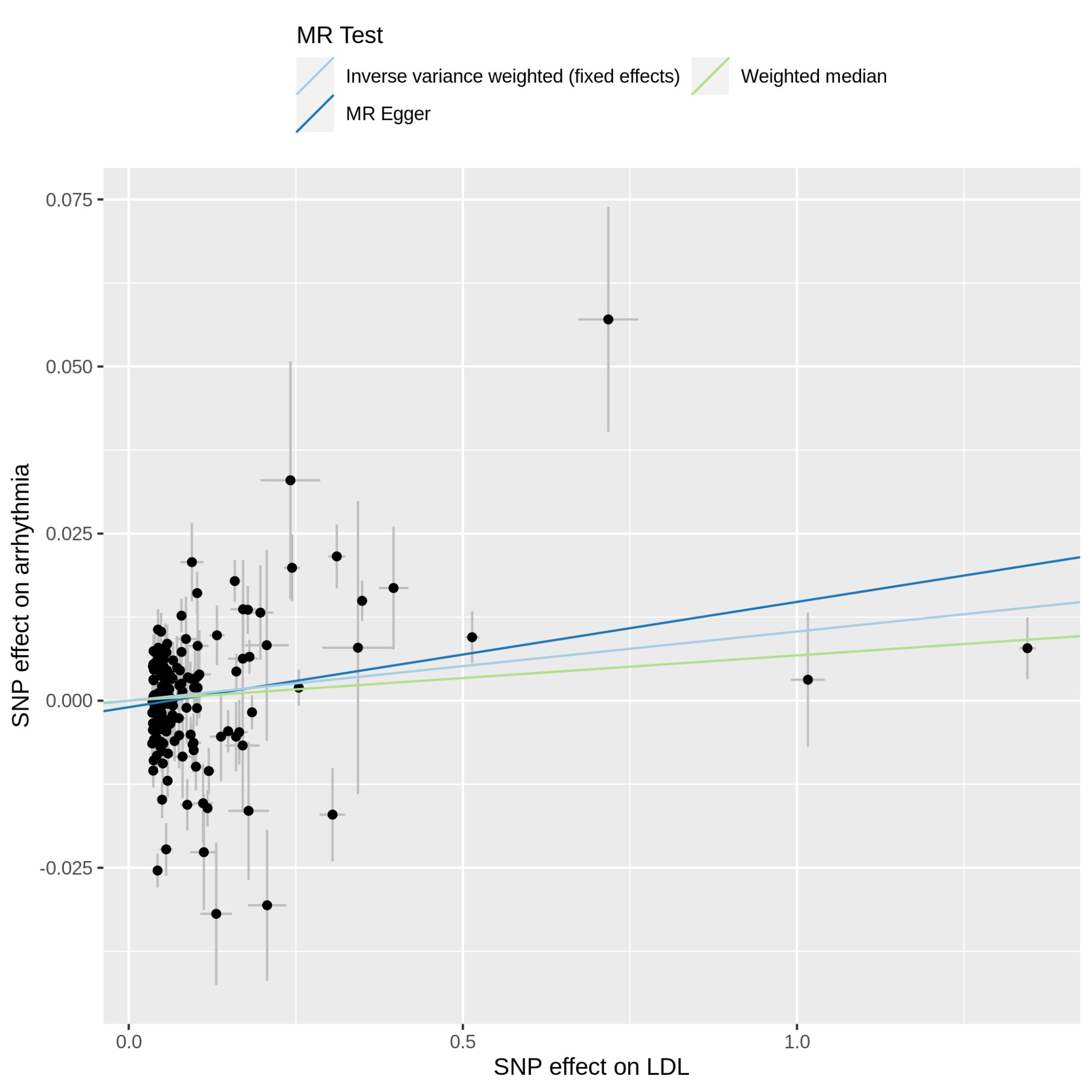

Supplementary Figure 11.18.1: MR scatter plot of LDL-C vs arrhythmia, based on the full set of LDL-C genetic instruments. The dot and horizontal bar along the x-axis denotes the GWAS marginal effect size estimate and 95% CI for LDL-C from the UKB cohort; the dot and vertical bar along the y-axis denotes the GWAS marginal effect size estimate and 95% CI for arrhythmia from the 23andMe cohort. The colored lines represent the 2SMR (IVW, MR-Egger, weighted median) estimates of genetically instrumented LDL-C onto arrhythmia.

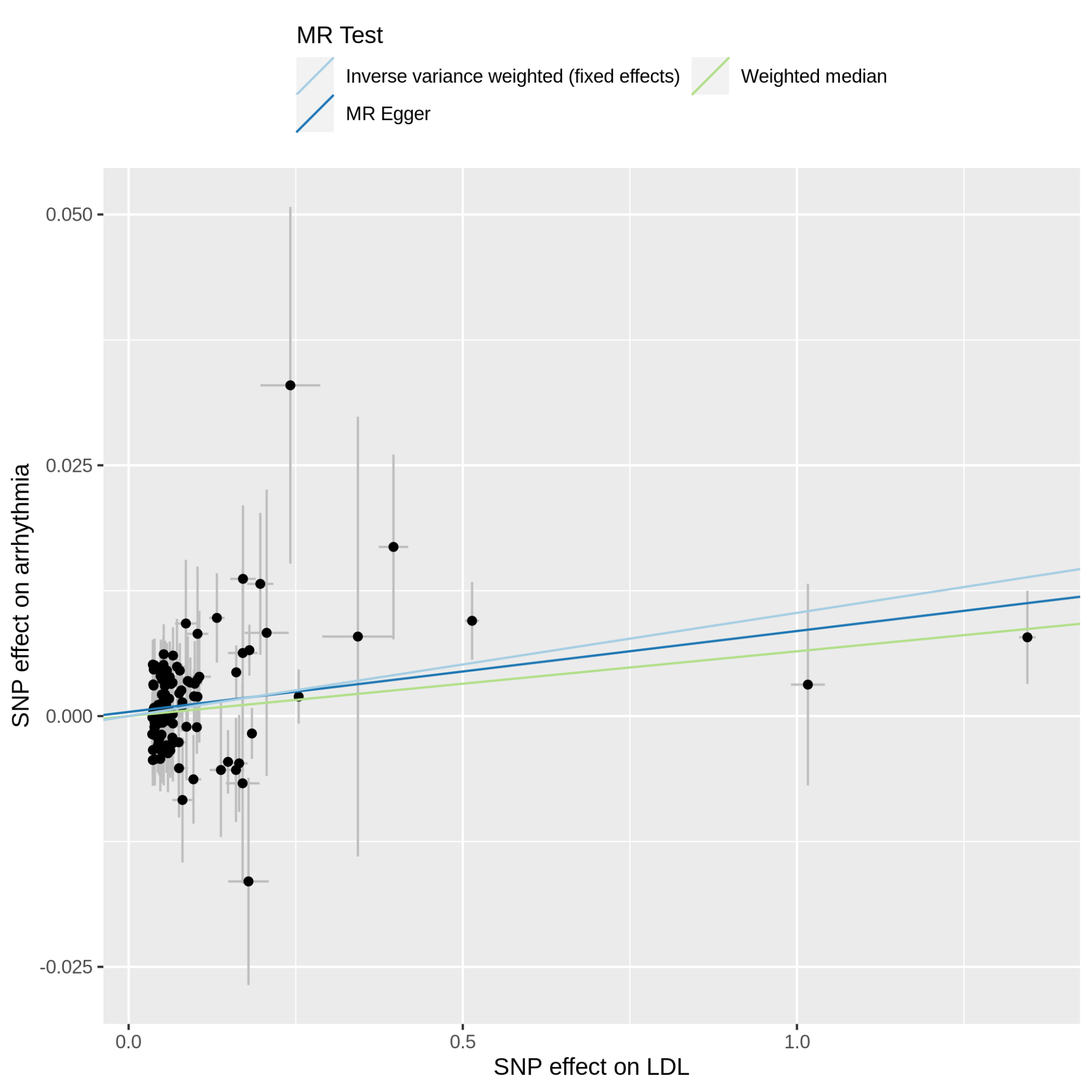

Supplementary Figure 11.18.2: MR scatter plot of LDL-C vs arrhythmia, based on the reduced set of LDL-C genetic instruments (after heterogeneity and Steiger filtering). The dot and horizontal bar along the x-axis denotes the GWAS marginal effect size estimate and 95% CI for LDL-C from the UKB cohort; the dot and vertical bar along the y-axis denotes the GWAS marginal effect size estimate and 95% CI for arrhythmia from the 23andMe cohort. The colored lines represent the 2SMR (IVW, MR-Egger, weighted median) estimates of genetically instrumented LDL-C onto arrhythmia.

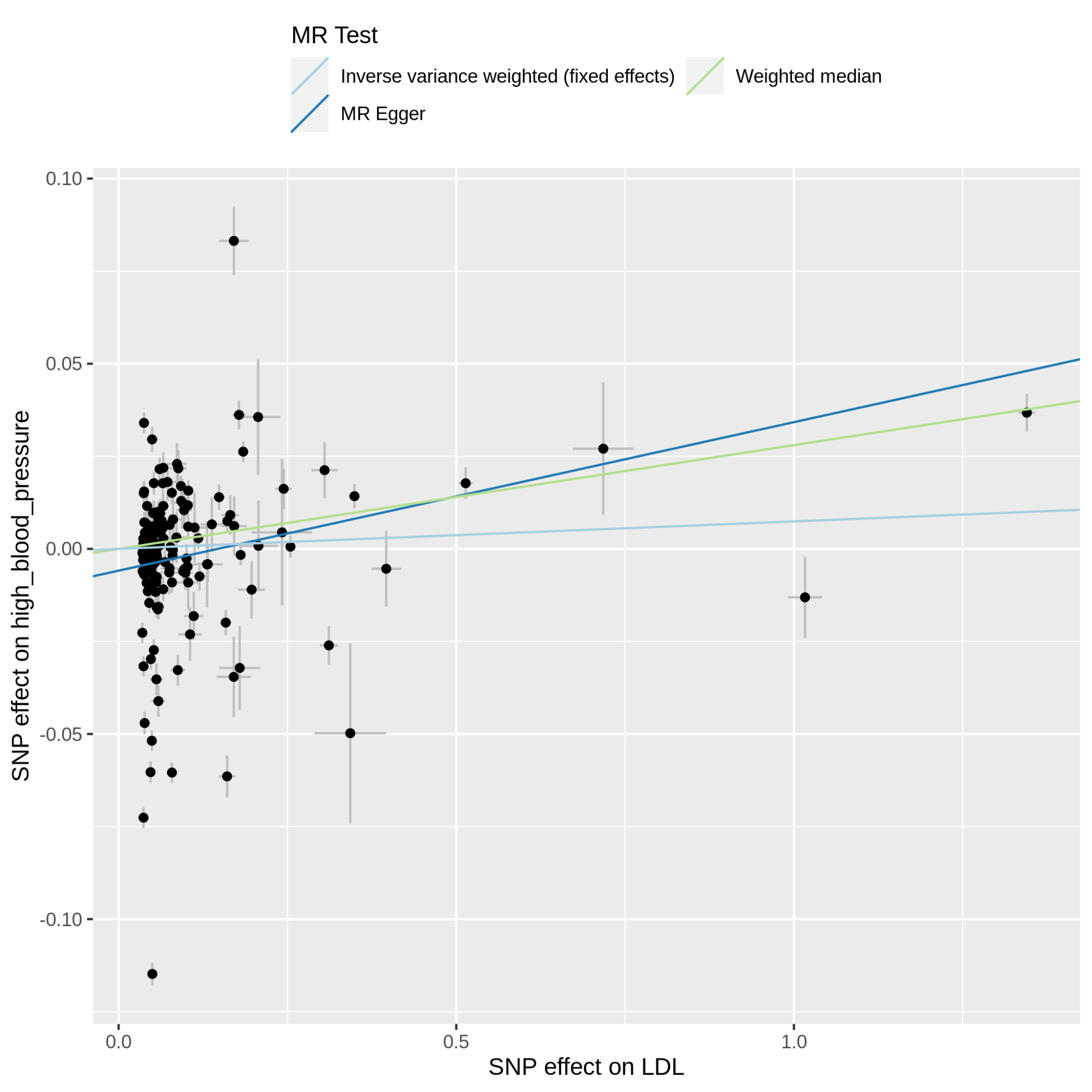

Supplementary Figure 11.19.1: MR scatter plot of LDL-C vs high_blood_pressure, based on the full set of LDL-C genetic instruments. The dot and horizontal bar along the x-axis denotes the GWAS marginal effect size estimate and 95% CI for LDL-C from the UKB cohort; the dot and vertical bar along the y-axis denotes the GWAS marginal effect size estimate and 95% CI for high_blood_pressure from the 23andMe cohort. The colored lines represent the 2SMR (IVW, MR-Egger, weighted median) estimates of genetically instrumented LDL-C onto high_blood_pressure.

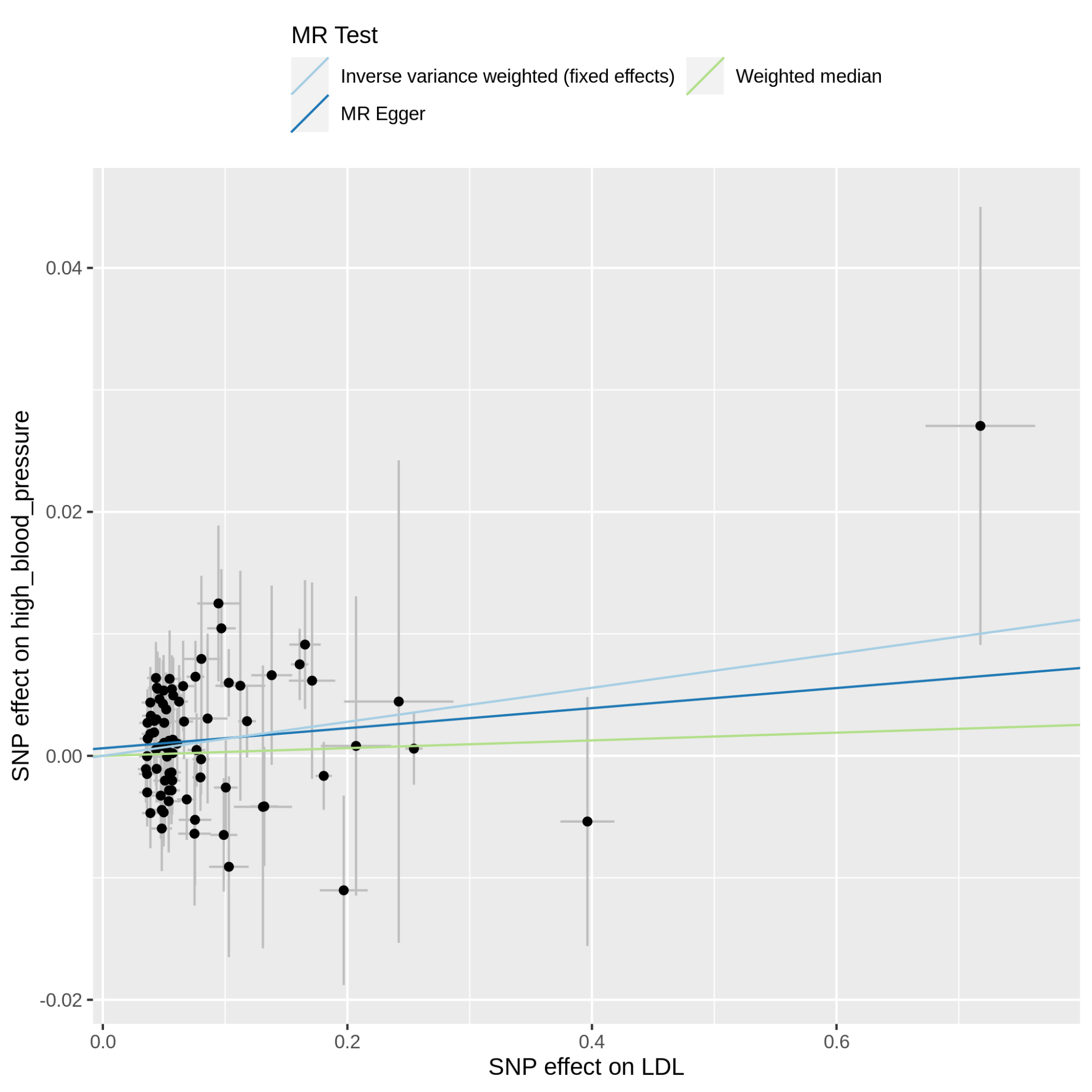

Supplementary Figure 11.19.2: MR scatter plot of LDL-C vs high_blood_pressure, based on the reduced set of LDL-C genetic instruments (after heterogeneity and Steiger filtering). The dot and horizontal bar along the x-axis denotes the GWAS marginal effect size estimate and 95% CI for LDL-C from the UKB cohort; the dot and vertical bar along the y-axis denotes the GWAS marginal effect size estimate and 95% CI for high_blood_pressure from the 23andMe cohort. The colored lines represent the 2SMR (IVW, MR-Egger, weighted median) estimates of genetically instrumented LDL-C onto high_blood_pressure.

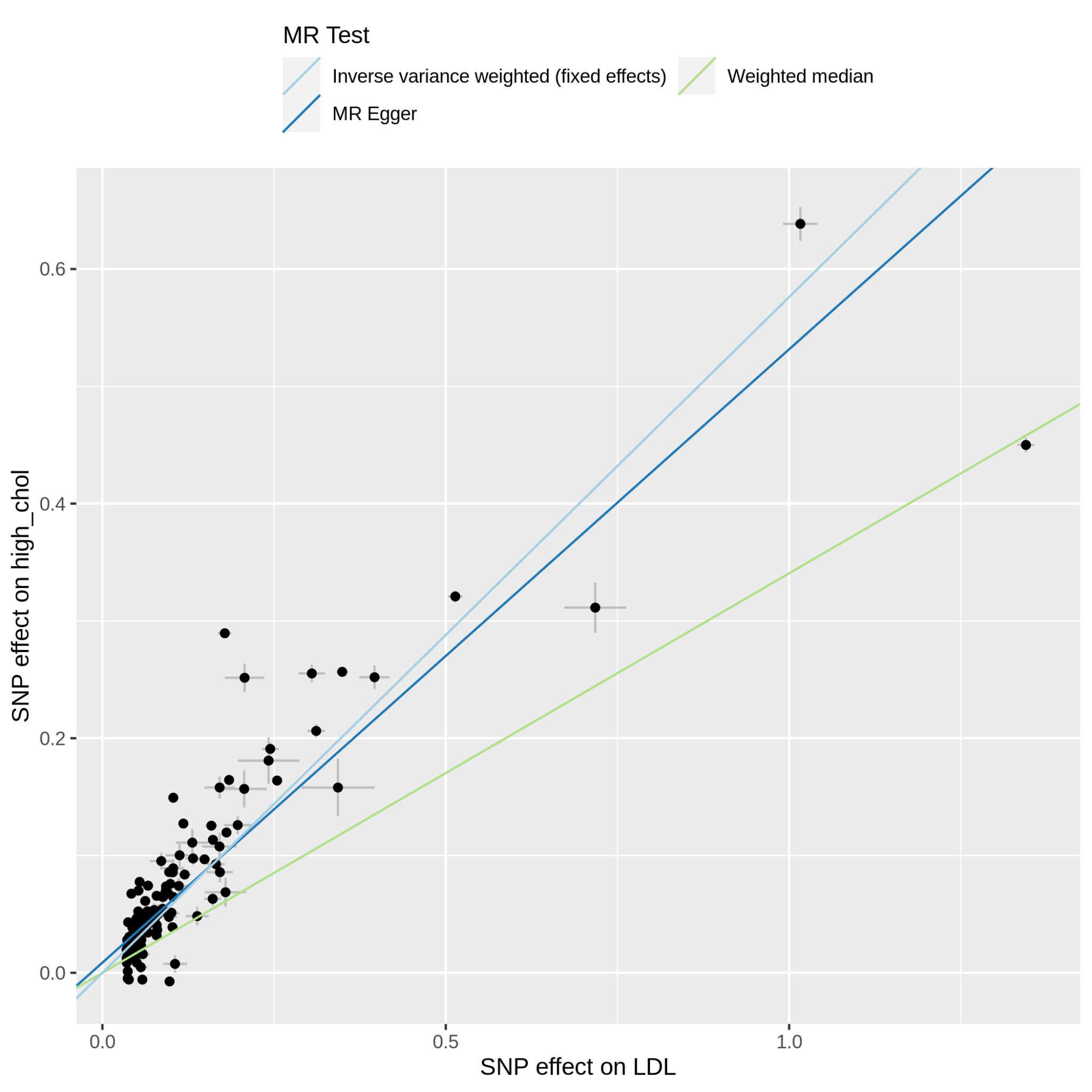

Supplementary Figure 11.20.1: MR scatter plot of LDL-C vs high_chol, based on the full set of LDL-C genetic instruments. The dot and horizontal bar along the x-axis denotes the GWAS marginal effect size estimate and 95% CI for LDL-C from the UKB cohort; the dot and vertical bar along the y-axis denotes the GWAS marginal effect size estimate and 95% CI for high_chol from the 23andMe cohort. The colored lines represent the 2SMR (IVW, MR-Egger, weighted median) estimates of genetically instrumented LDL-C onto high_chol.

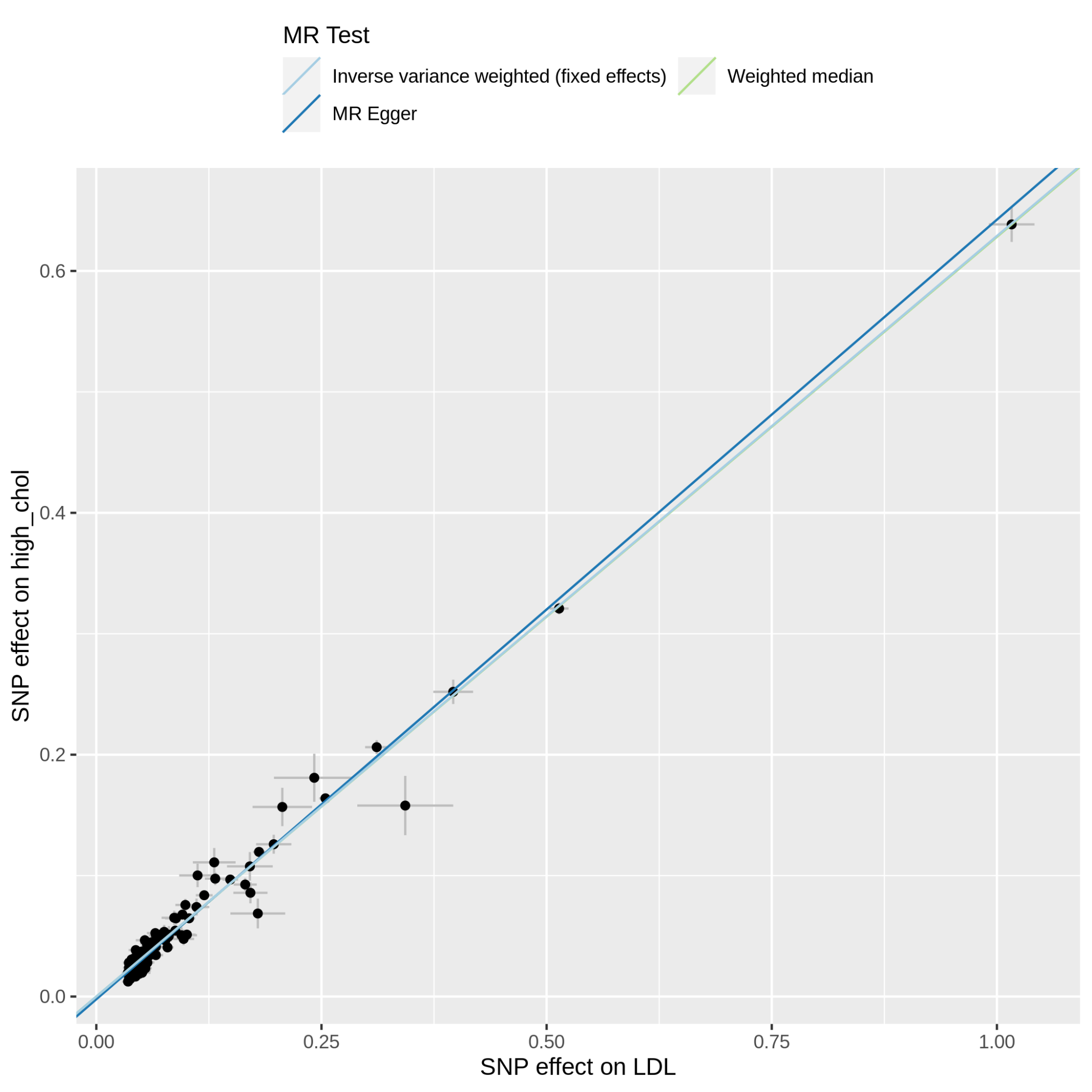

Supplementary Figure 11.20.2: MR scatter plot of LDL-C vs high_chol, based on the reduced set of LDL-C genetic instruments (after heterogeneity and Steiger filtering). The dot and horizontal bar along the x-axis denotes the GWAS marginal effect size estimate and 95% CI for LDL-C from the UKB cohort; the dot and vertical bar along the y-axis denotes the GWAS marginal effect size estimate and 95% CI for high_chol from the 23andMe cohort. The colored lines represent the 2SMR (IVW, MR-Egger, weighted median) estimates of genetically instrumented LDL-C onto high_chol.

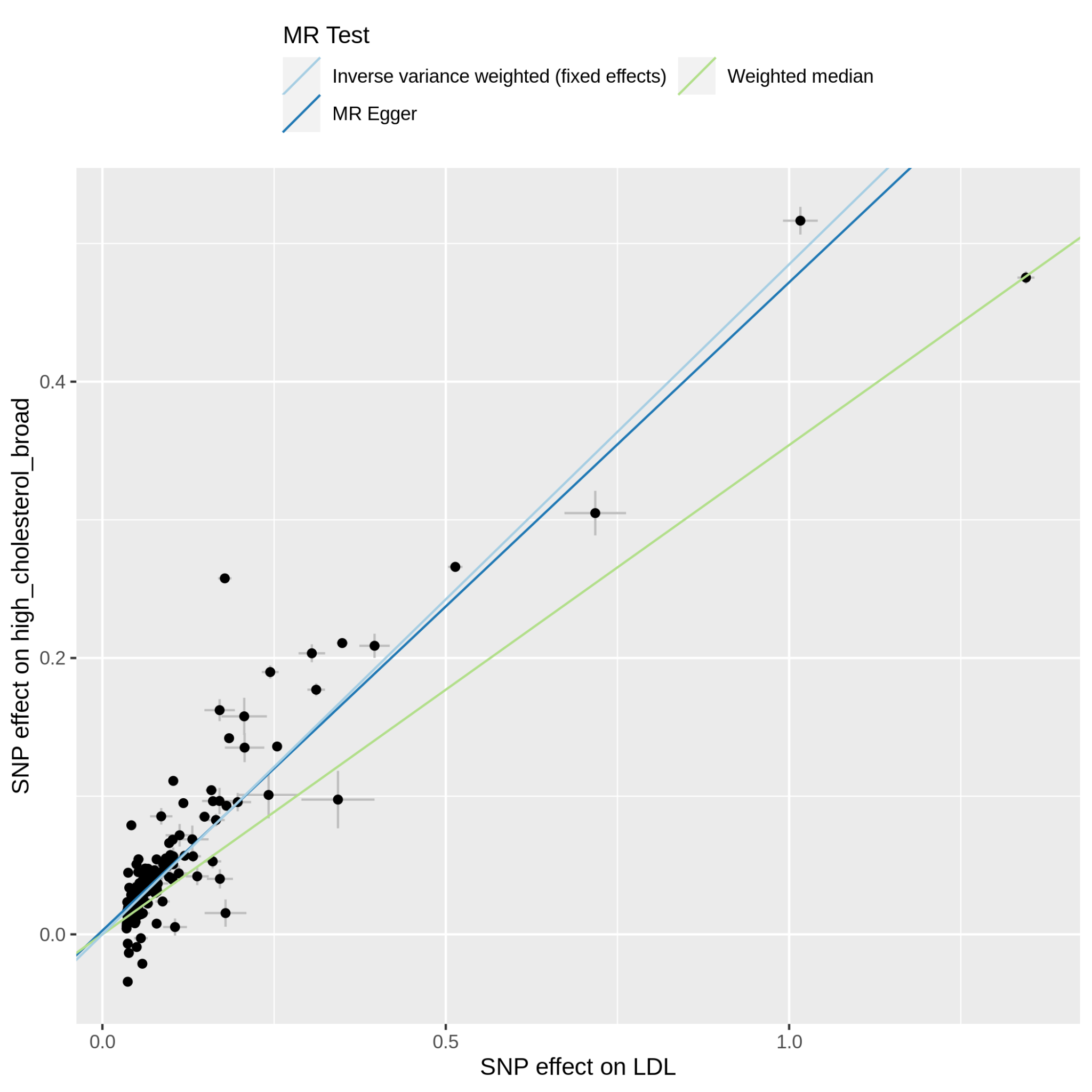

Supplementary Figure 11.21.1: MR scatter plot of LDL-C vs high_cholesterol_broad, based on the full set of LDL-C genetic instruments. The dot and horizontal bar along the x-axis denotes the GWAS marginal effect size estimate and 95% CI for LDL-C from the UKB cohort; the dot and vertical bar along the y-axis denotes the GWAS marginal effect size estimate and 95% CI for high_cholesterol_broad from the 23andMe cohort. The colored lines represent the 2SMR (IVW, MR-Egger, weighted median) estimates of genetically instrumented LDL-C onto high_cholesterol_broad.

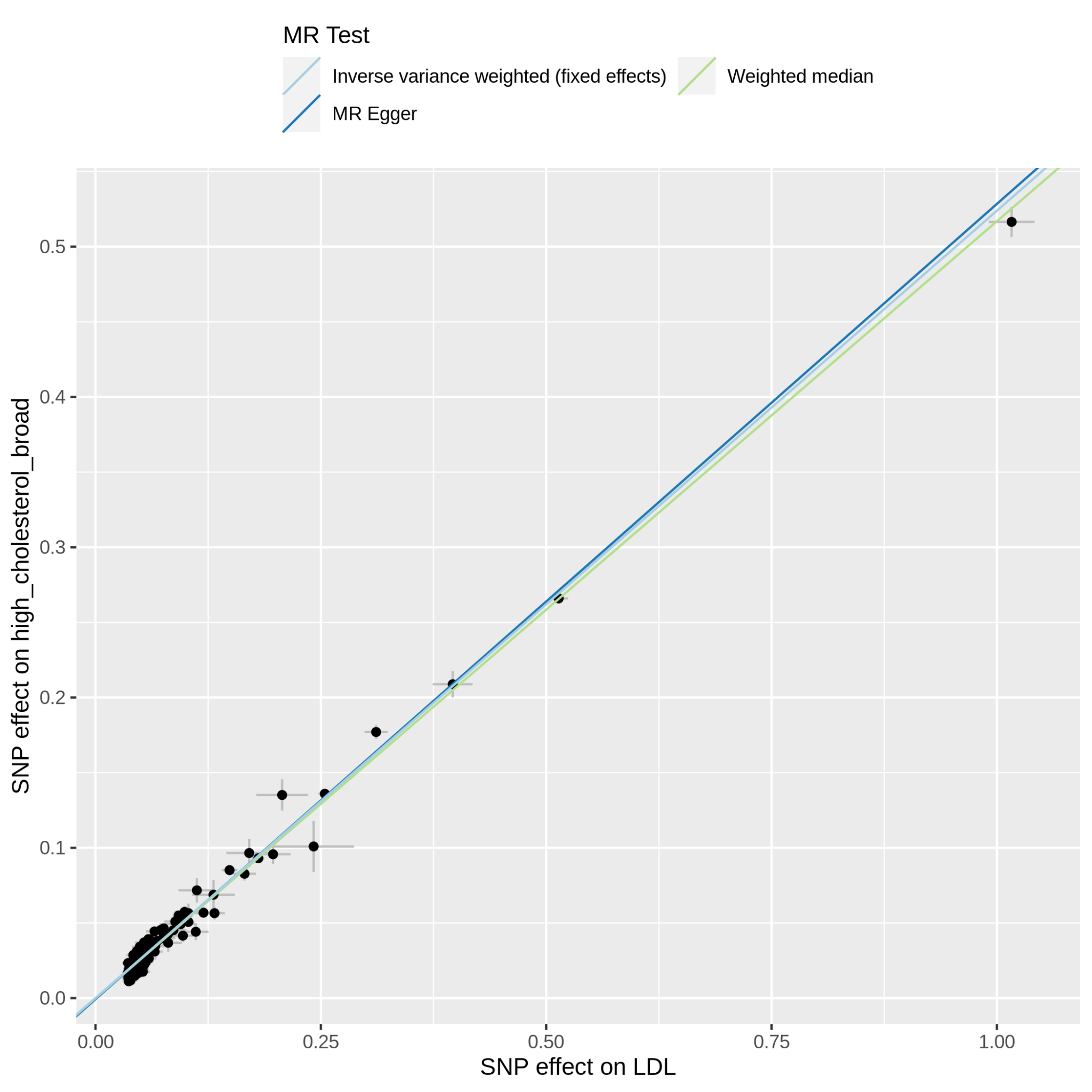

Supplementary Figure 11.21.2: MR scatter plot of LDL-C vs high_cholesterol_broad, based on the reduced set of LDL-C genetic instruments (after heterogeneity and Steiger filtering). The dot and horizontal bar along the x-axis denotes the GWAS marginal effect size estimate and 95% CI for LDL-C from the UKB cohort; the dot and vertical bar along the y-axis denotes the GWAS marginal effect size estimate and 95% CI for high_cholesterol_broad from the 23andMe cohort. The colored lines represent the 2SMR (IVW, MR-Egger, weighted median) estimates of genetically instrumented LDL-C onto high_cholesterol_broad.

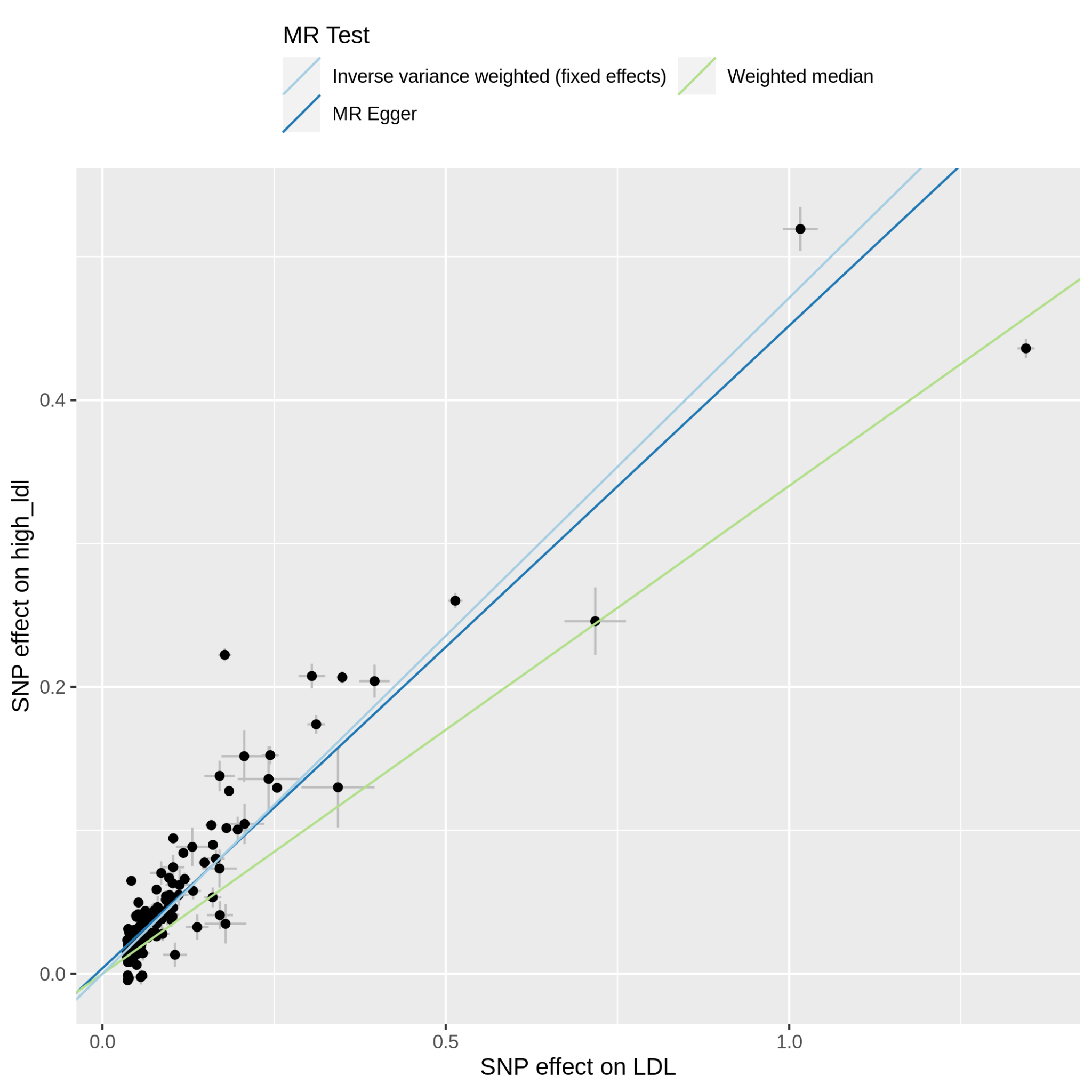

Supplementary Figure 11.22.1: MR scatter plot of LDL-C vs high_ldl, based on the full set of LDL-C genetic instruments. The dot and horizontal bar along the x-axis denotes the GWAS marginal effect size estimate and 95% CI for LDL-C from the UKB cohort; the dot and vertical bar along the y-axis denotes the GWAS marginal effect size estimate and 95% CI for high_ldl from the 23andMe cohort. The colored lines represent the 2SMR (IVW, MR-Egger, weighted median) estimates of genetically instrumented LDL-C onto high_ldl.

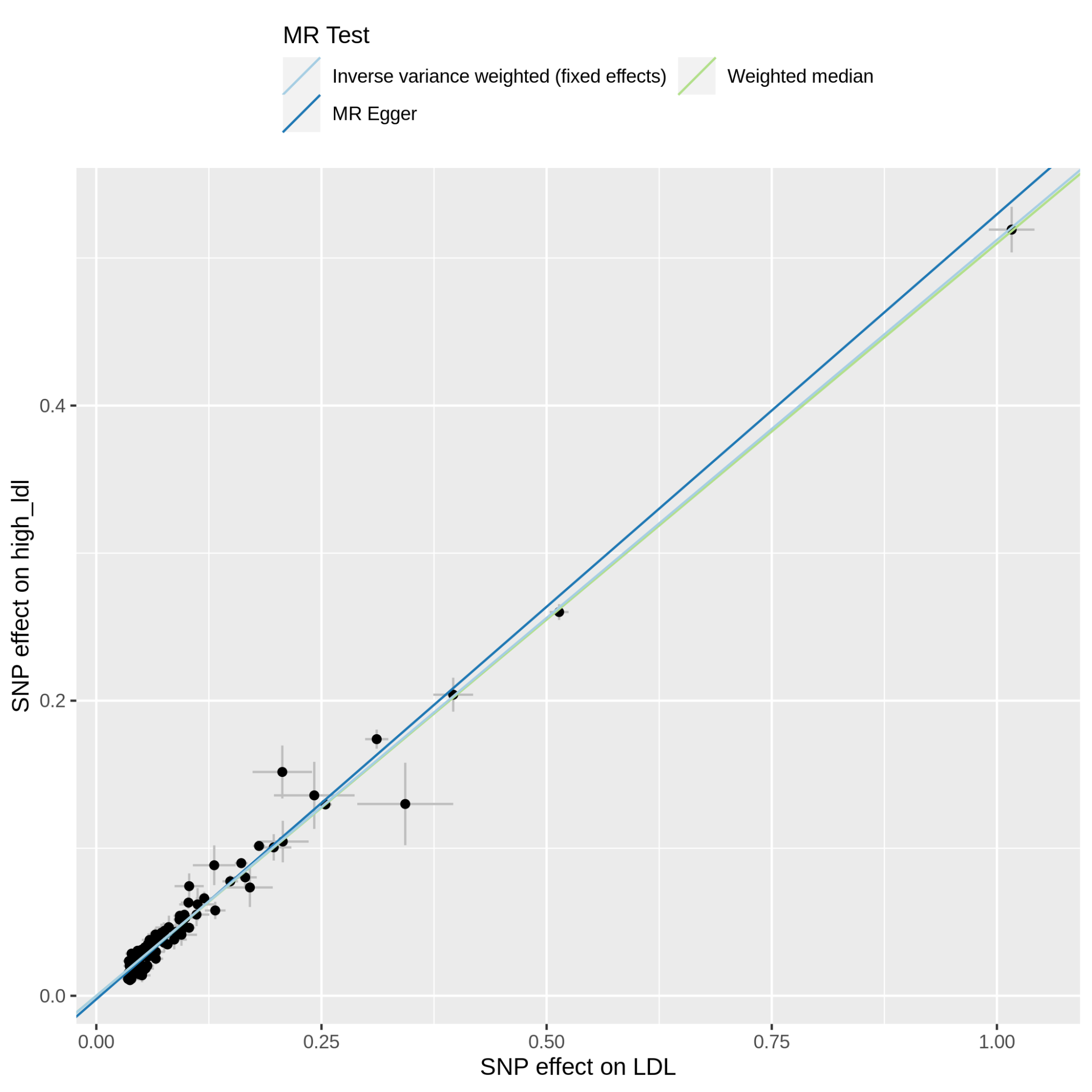

Supplementary Figure 11.22.2: MR scatter plot of LDL-C vs high_ldl, based on the reduced set of LDL-C genetic instruments (after heterogeneity and Steiger filtering). The dot and horizontal bar along the x-axis denotes the GWAS marginal effect size estimate and 95% CI for LDL-C from the UKB cohort; the dot and vertical bar along the y-axis denotes the GWAS marginal effect size estimate and 95% CI for high_ldl from the 23andMe cohort. The colored lines represent the 2SMR (IVW, MR-Egger, weighted median) estimates of genetically instrumented LDL-C onto high_ldl.

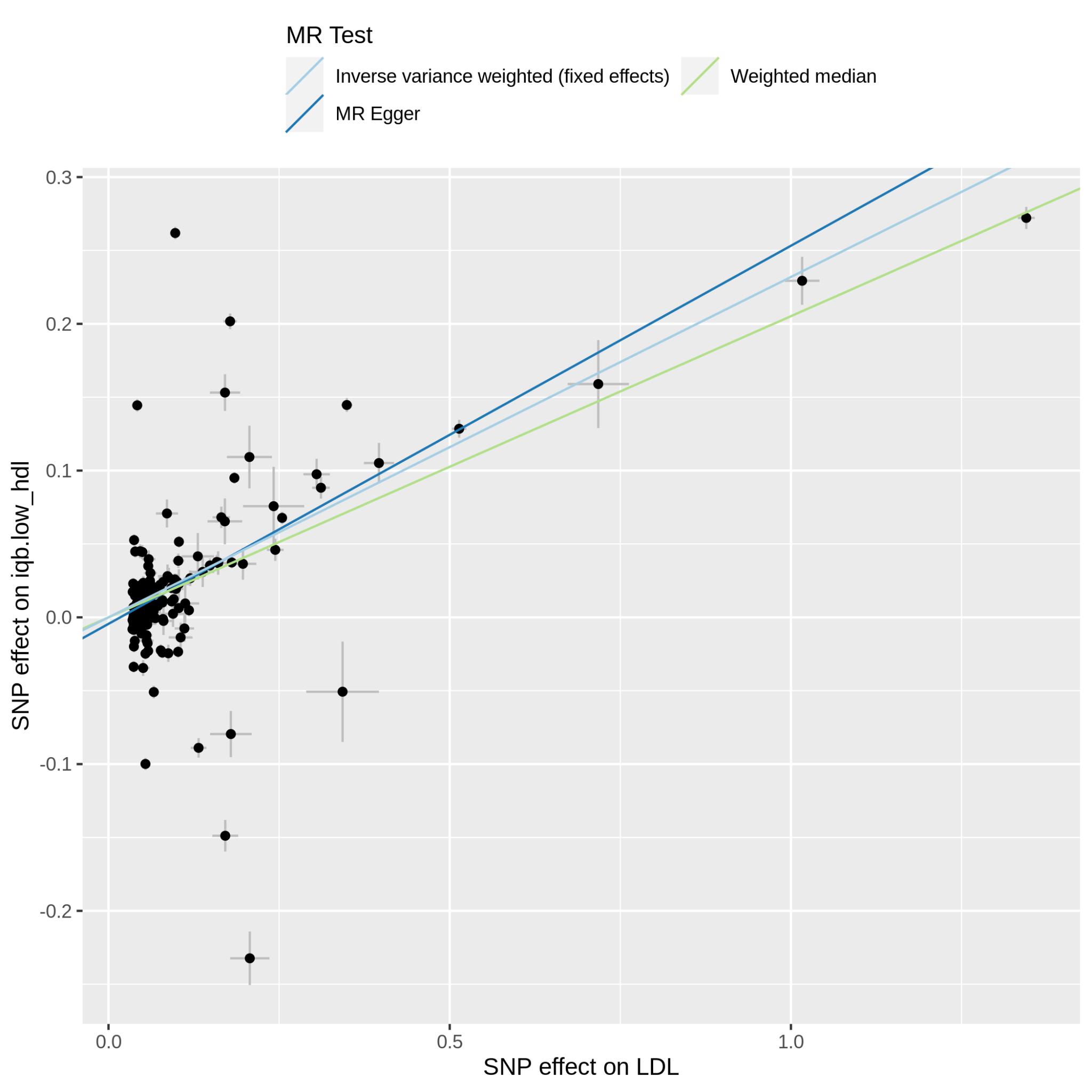

Supplementary Figure 11.23.1: MR scatter plot of LDL-C vs iqb.low_hdl, based on the full set of LDL-C genetic instruments. The dot and horizontal bar along the x-axis denotes the GWAS marginal effect size estimate and 95% CI for LDL-C from the UKB cohort; the dot and vertical bar along the y-axis denotes the GWAS marginal effect size estimate and 95% CI for iqb.low_hdl from the 23andMe cohort. The colored lines represent the 2SMR (IVW, MR-Egger, weighted median) estimates of genetically instrumented LDL-C onto iqb.low_hdl.

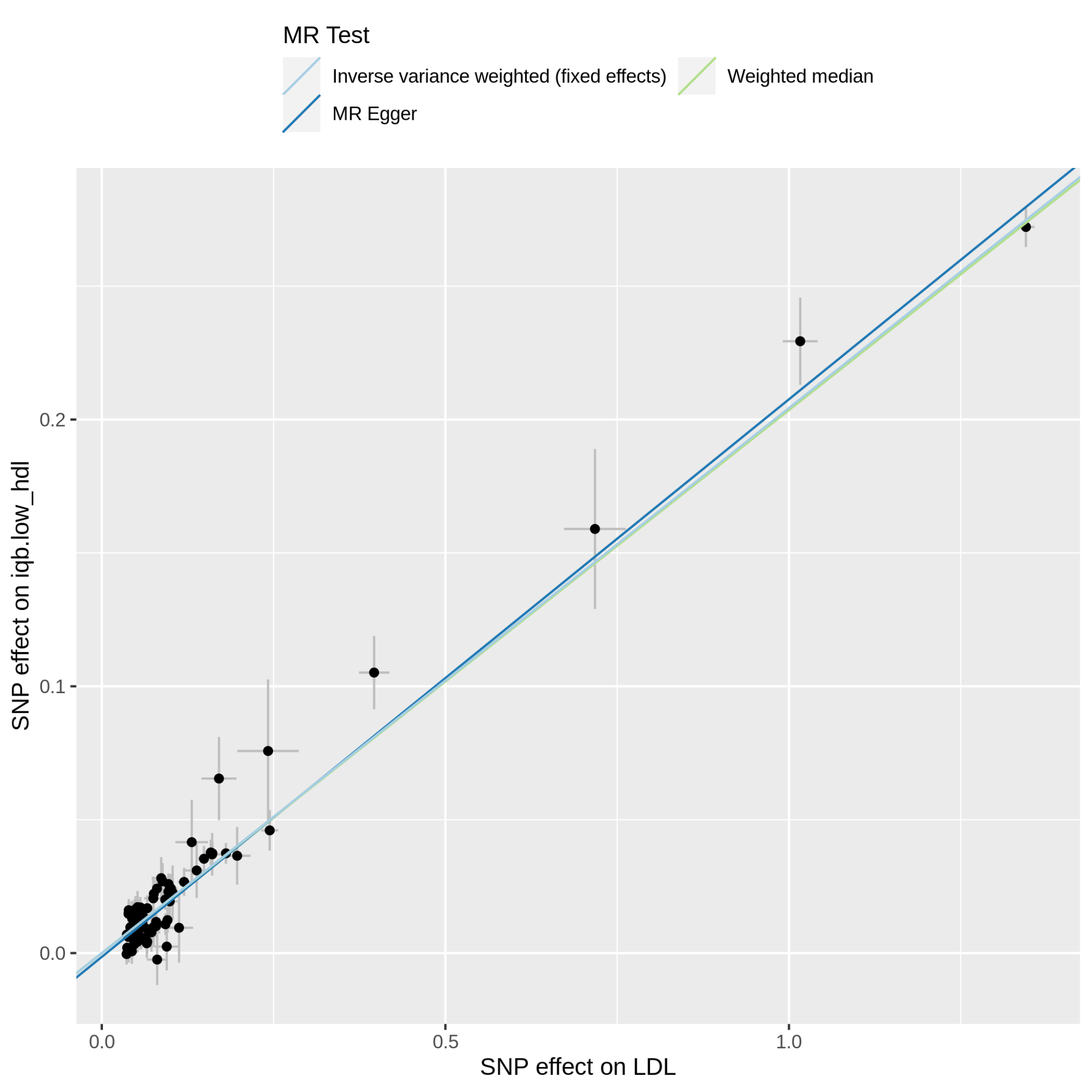

Supplementary Figure 11.23.2: MR scatter plot of LDL-C vs iqb.low_hdl, based on the reduced set of LDL-C genetic instruments (after heterogeneity and Steiger filtering). The dot and horizontal bar along the x-axis denotes the GWAS marginal effect size estimate and 95% CI for LDL-C from the UKB cohort; the dot and vertical bar along the y-axis denotes the GWAS marginal effect size estimate and 95% CI for iqb.low_hdl from the 23andMe cohort. The colored lines represent the 2SMR (IVW, MR-Egger, weighted median) estimates of genetically instrumented LDL-C onto iqb.low_hdl.

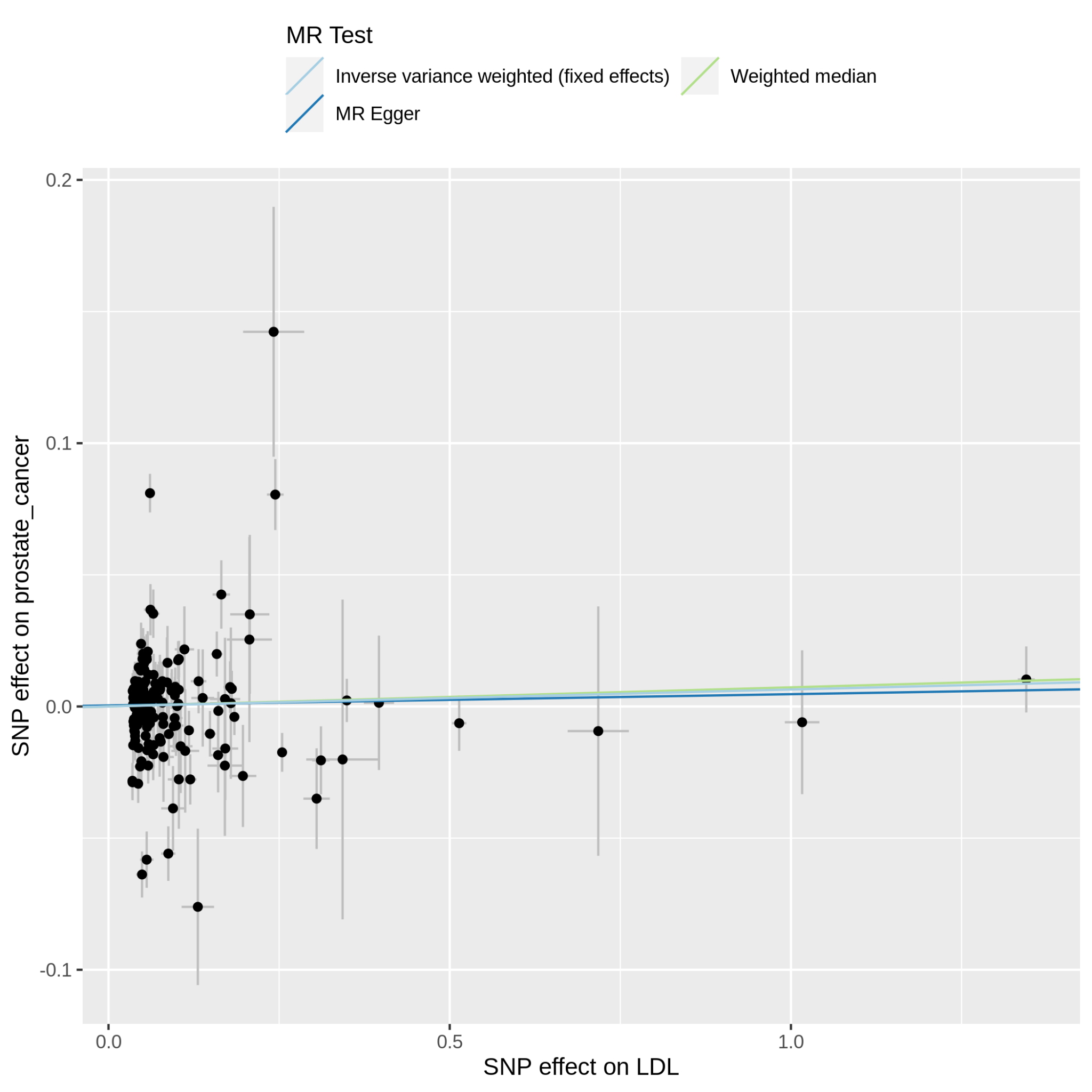

Supplementary Figure 11.24.1: MR scatter plot of LDL-C vs prostate_cancer, based on the full set of LDL-C genetic instruments. The dot and horizontal bar along the x-axis denotes the GWAS marginal effect size estimate and 95% CI for LDL-C from the UKB cohort; the dot and vertical bar along the y-axis denotes the GWAS marginal effect size estimate and 95% CI for prostate_cancer from the 23andMe cohort. The colored lines represent the 2SMR (IVW, MR-Egger, weighted median) estimates of genetically instrumented LDL-C onto prostate_cancer.

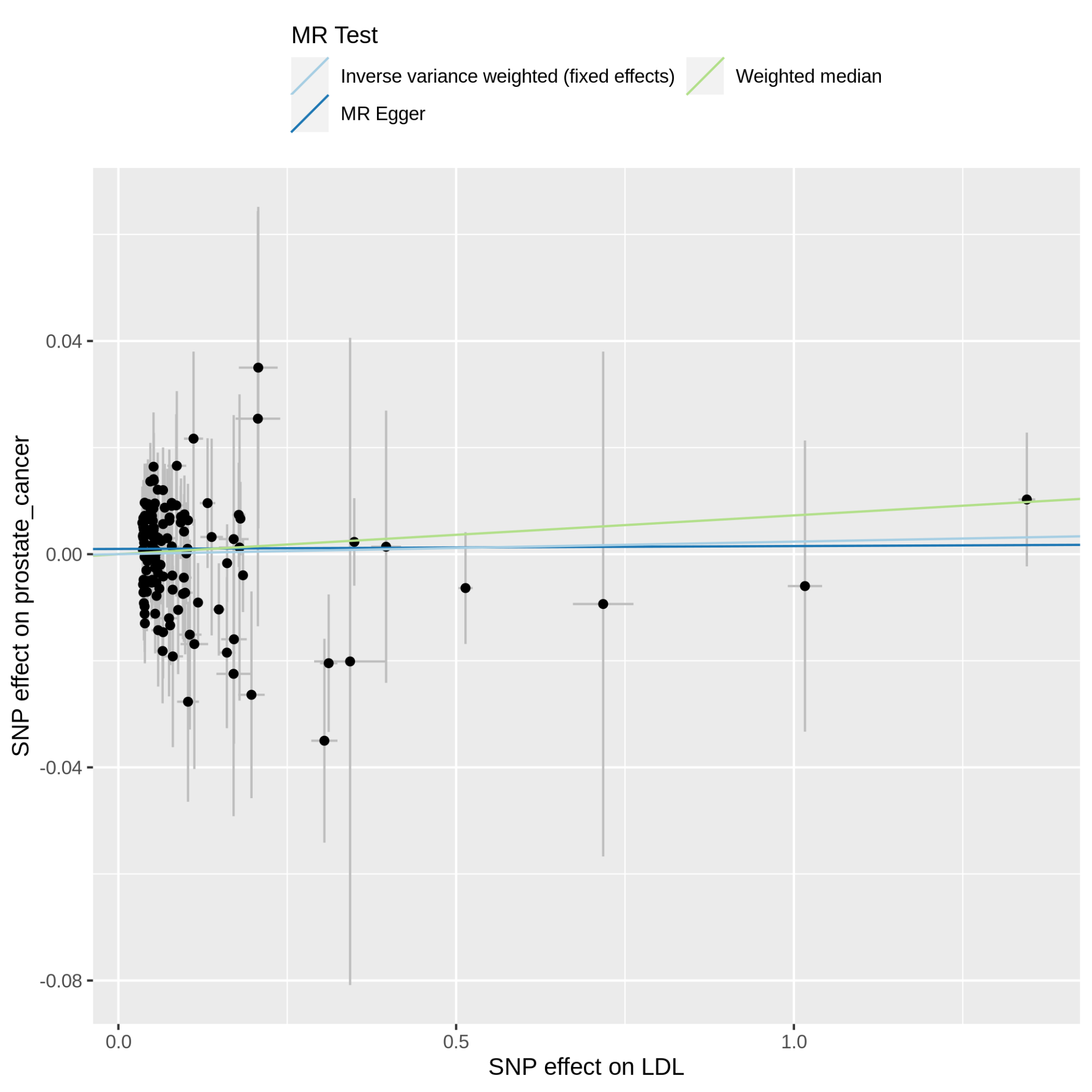

Supplementary Figure 11.24.2: MR scatter plot of LDL-C vs prostate_cancer, based on the reduced set of LDL-C genetic instruments (after heterogeneity and Steiger filtering). The dot and horizontal bar along the x-axis denotes the GWAS marginal effect size estimate and 95% CI for LDL-C from the UKB cohort; the dot and vertical bar along the y-axis denotes the GWAS marginal effect size estimate and 95% CI for prostate_cancer from the 23andMe cohort. The colored lines represent the 2SMR (IVW, MR-Egger, weighted median) estimates of genetically instrumented LDL-C onto prostate_cancer.

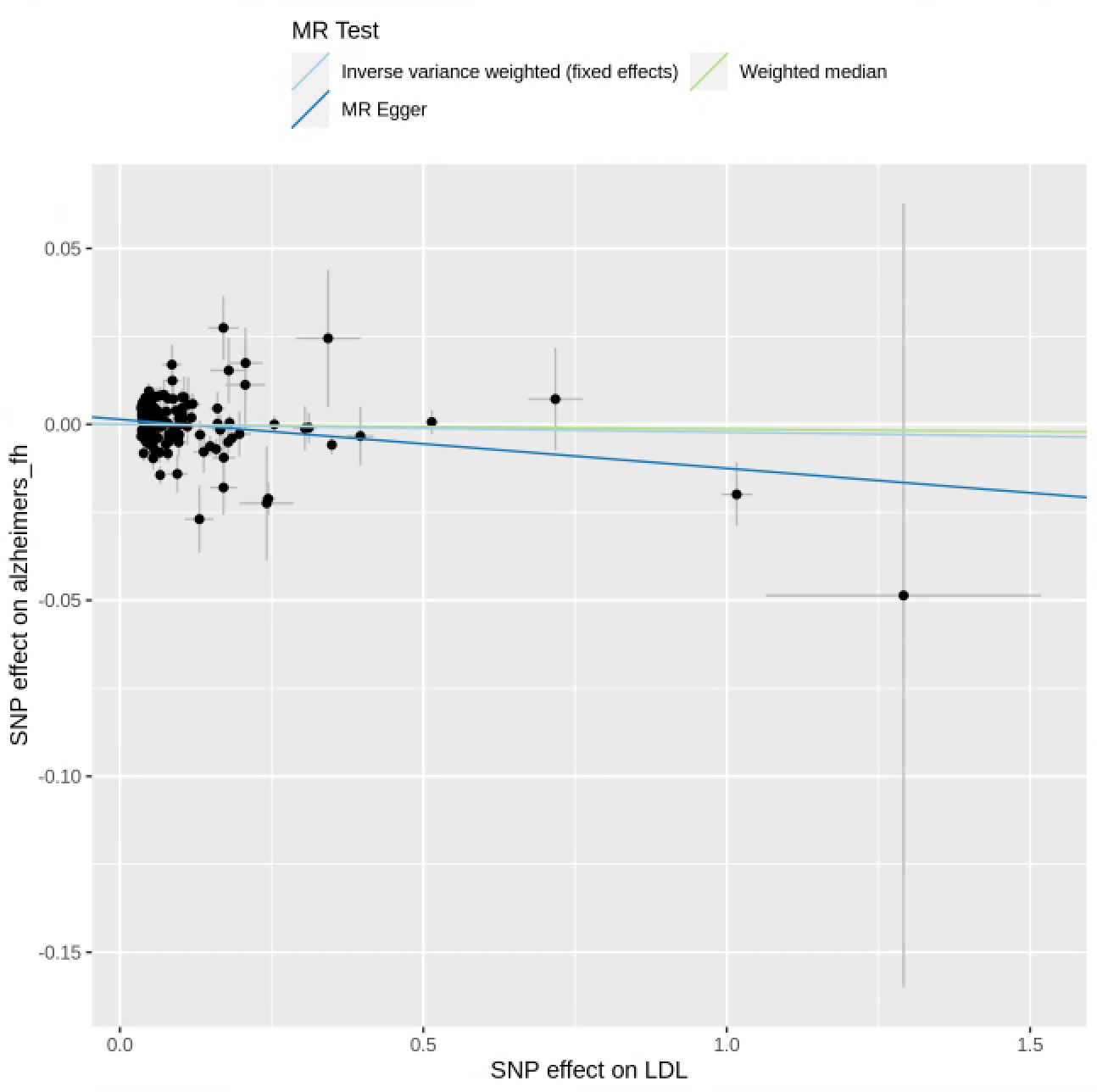

Supplementary Figure 11.25.1: MR scatter plot of LDL-C vs alzheimers_fh, based on the full set of LDL-C genetic instruments. The dot and horizontal bar along the x-axis denotes the GWAS marginal effect size estimate and 95% CI for LDL-C from the UKB cohort; the dot and vertical bar along the y-axis denotes the GWAS marginal effect size estimate and 95% CI for alzheimers_fh from the 23andMe cohort. The colored lines represent the 2SMR (IVW, MR-Egger, weighted median) estimates of genetically instrumented LDL-C onto alzheimers_fh.

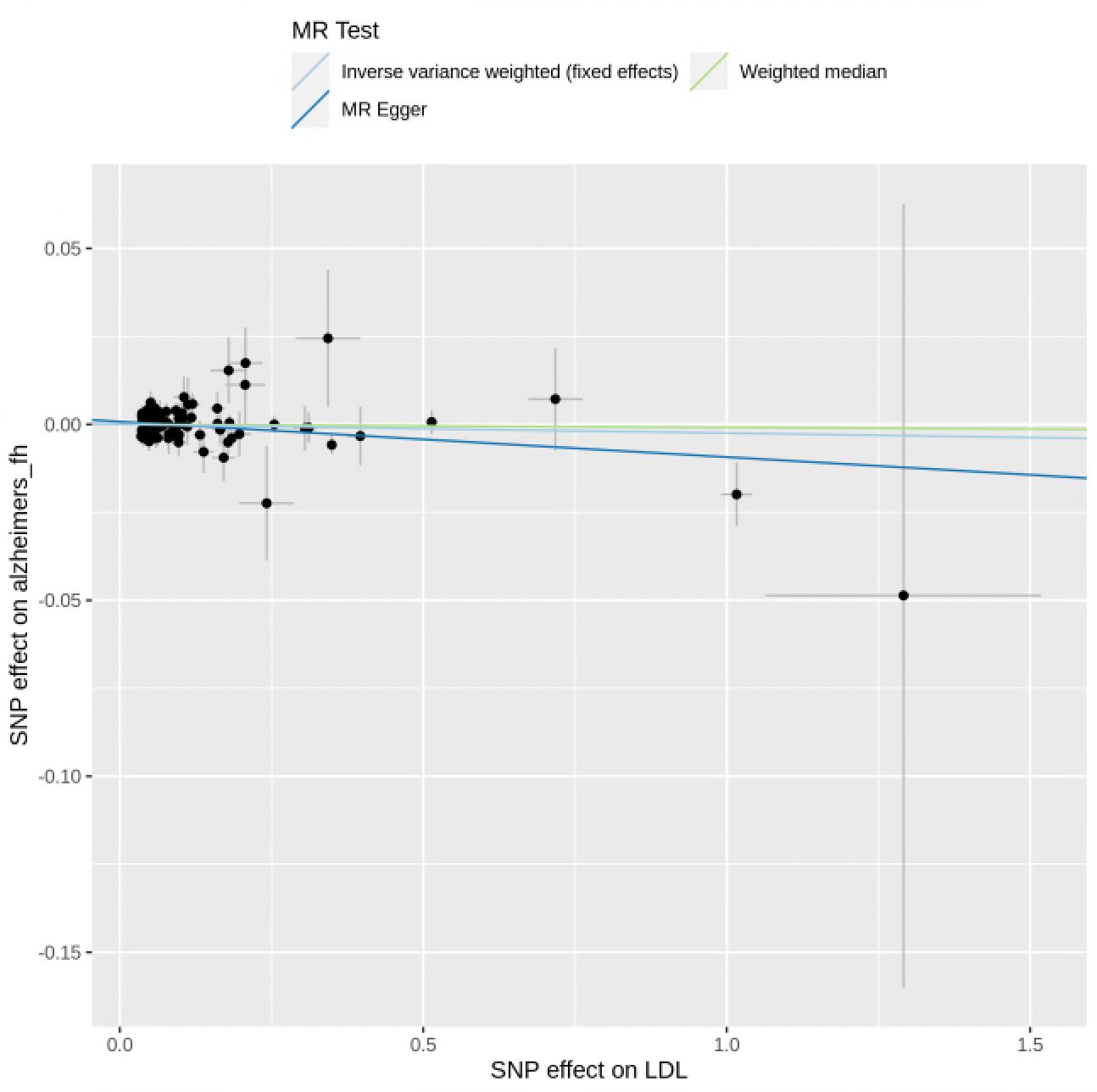

Supplementary Figure 11.25.2: MR scatter plot of LDL-C vs alzheimers_fh, based on the reduced set of LDL-C genetic instruments (after heterogeneity and Steiger filtering). The dot and horizontal bar along the x-axis denotes the GWAS marginal effect size estimate and 95% CI for LDL-C from the UKB cohort; the dot and vertical bar along the y-axis denotes the GWAS marginal effect size estimate and 95% CI for alzheimers_fh from the 23andMe cohort. The colored lines represent the 2SMR (IVW, MR-Egger, weighted median) estimates of genetically instrumented LDL-C onto alzheimers_fh.
