## Supplementary Notes for "Evaluating genetically-predicted causal effects of lipoprotein(a) in human diseases: a phenome-wide Mendelian randomization study"

### Genetic Association Analysis

We performed genome-wide association analysis (GWAS) on the UK Biobank Lp(a), LDL-C and apoB measurements (90% training samples) using REGENIE v3.2.5. We selected a set of variants with high genotyping quality for the first step of REGENIE. The variant selection criteria included: minor allele frequency (MAF) > 0.1, minor allele count (MAC) > 100, Hardy-Weinberg equilibrium (HWE) test p-value > $10^{-15}$ and genotyping missing call rate < 10% (Sun, B.B *et al.*, 2022). The first step of REGENIE yielded leave one chromosome out (LOCO) phenotypic predictions, which were incorporated as offsets in the second step of REGENIE to perform variant association analyses using a standard linear regression model. In the second step of REGENIE, we focused on ~13.7 million common variants with high imputation quality: MAF > 0.001 and imputation INFO > 0.8. The association models for all three lipid traits included the following covariates: age, $age^2$, sex, age × sex, $age^2$ × sex, dummy variables for genotyping arrays, and the first 20 genetic principal components (PCs).

To compensate for GWAS variance inflation due to high polygenicity, we performed LDscore regression (Bulik-Sullivan, B.K. *et al.*, 2015) implemented with "LDSC" v1.0.0 as a genomic control procedure. The LD scores were estimated from the 1000G phase 3 reference panel, based on 5.5 million variants. We used the LD score regression intercept as the genomic control inflation factor ($\lambda$) to adjust the three lipid biomarkers' GWAS effect sizes and standard errors. Specifically, $\lambda = 1.112$ for Lp(a), $\lambda = 1.226$ for LDL-C, and $\lambda = 1.256$ for apoB.

Based on the $\lambda$-adjusted GWAS results, next we identified loci with genome-wide significant associations. We first gathered all SNPs with p-value $< 10^{-5}$ within the vicinity of a genome-wide significant association to define the region boundaries. We then grouped these regions into loci so that two adjacent loci were separated by at least 250 kilobases (kb). We chose the SNP with the smallest p-value in a locus as the lead SNP. The Manhattan and QQ plots for each of those lipid biomarkers are shown in Supplementary Figure 1 - 3.

### *LPA* GRS Instrument Selection for Univariable MR

To pinpoint likely causal variants underlying the Lp(a) genetic association in the *LPA* locus (Supplementary Figure 4.1), we performed statistical fine-mapping with the SuSiE approach (Wang, G. *et al.*, 2020), which accounts for correlations between SNPs due to high LD when selecting SNPs for the credible set construction. We built an in-sample LD reference panel from the Lp(a) training cohort samples and computed the LD matrix using plink2 (Chang, C. C. *et al.*, 2015). We used the Lp(a) training cohort GWAS summary statistics and its in-sample LD panel to run the SuSiE algorithm within the 1 Mb window centered at the transcription start site (TSS) of the *LPA* gene (GRCh38: chr6: 159,664,259 - 161,664,259). We set the SuSiE algorithm parameters to allow up to 40 credible sets, with a minimum squared correlation equals to 0.3 allowed in a credible set, and the remaining parameters set to their default values. The algorithm returned 32 credible sets, each containing only one SNP with a posterior inclusion probability (PIP) greater than 0.95 (Supplementary Figure 4.2). We next performed a multivariable linear regression model including those 32 SNPs, with the same set of adjusting covariates used in the Lp(a) GWAS run, plus the Lp(a) leave chromosome 6 out phenotypic prediction from the first step of Lp(a) REGENIE as an offset to account for population structure. In this multivariable linear regression model, three SNPs were no longer nominally significant (p-value > 0.01) when conditioning on the other SNPs. Thus, we further removed those three SNPs which do not contribute much in the aggregated model, and used the conditional effect sizes of the remaining 29 SNPs (Supplementary Table 2) as the genetic instruments for the *LPA* GRS. We constructed *LPA* GRS on UKB 10% testing samples to evaluate the prediction performance, and on ~7.3M 23andMe research participants to perform the univariable 2SLS MR analysis.

**LDL-C and apoB GRS Instrument Selection for Univariable MR**

The λ-adjusted GWASs yielded 169 and 175 genome-wide significant hits for LDL-C and apoB, respectively (Supplementary Figure 2.1, 3.1). We used the GWAS marginal effect sizes of those SNPs to construct the LDL-C and apoB GRS (Supplementary Table 3, 4) on UKB 10% testing samples to evaluate the prediction performance of the LDL-C and apoB GRS, and on ~7.3M 23andMe research participants to perform the univariable 2SLS MR analysis.

Among the 169 SNPs used for constructing the LDL-C GRS, only one SNP (rs118039278) came from the *LPA* locus and explained 0.17% of the variation in LDL-C (Supplementary Table 3). Together the 169 SNPs explained 9.18% of the LDL-C variation in total, which means the SNP from *LPA* locus (rs118039278) accounted for only 1.85% among the variance explained by the 169 SNPs. Among the 175 SNPs used for constructing the apoB GRS, only one SNP (rs10455872) came from the *LPA* locus and explained 0.06% of the variation in apoB (Supplementary Table 4). The 175 SNPs explained 11.16% of the apoB variation in total, which means the SNP from *LPA* locus (rs10455872) accounted for only 0.54% among the variance explained. Additionally, our distance-based lead SNP selection algorithm (as described in the Genetic Association Analysis section in this Supplementary Notes) assures that any genetic instruments for LDL-C and apoB that are outside of the *LPA* locus would be in low LD with those 29 genetic instruments for Lp(a) which all reside in the *LPA* locus. As such, we are not concerned with including the *LPA* locus in the LDL-C and apoB GRS construction.

Note: *APOE* is a known strong risk factor for Alzheimer's disease and associated with higher LDL-C levels. In our case, both the LDL-C and apoB GRS included one variant located in the *APOE* region, namely, rs1065853. Inclusion of such a variant from the *APOE* region in the LDL-C and apoB GRS can potentially induce an association between the lipid traits and Alzheimer's disease due to horizontal pleiotropy through *APOE*. Previous studies (Wingo, A.P *et al.*, 2022; Korthauer, L.E. *et al.*, 2022) reported that LDL-C is a risk factor for Alzheimer's disease independent of *APOE*. We initially observed strong associations for genetically predicted LDL-C and apoB levels with the family history of Alzheimer's disease when we included rs1065853 as a genetic instrument for those two lipid entities: LDL-C GRS with Alzheimer's family history had OR = 1.037, 95% CI = [1.034, 1.041], p-value = $5.178\times10^{-90}$; apoB GRS with Alzheimer's family history had OR = 1.041, 95% CI = [1.037, 1.044], p-value = $2.596\times10^{-118}$. And those signals became null using the *APOE*-depleted GRS after we removed rs1065853 from the genetic instruments for LDL-C and apoB (both p-values > 0.3). We also observed that the univariable MR results for LDL-C and apoB on any other phenotypes were very similar with or without rs1065853 included in the GRS. Thus, we reported the *APOE*-depleted univariable MR results for LDL-C and apoB on Alzheimer's family history but kept the full sets of genetic instruments on other phenotypes in Supplementary Table 5.

**GRS Instrument Selection for Multivariable MR**

In the multivariable MR of Lp(a) with LDL-C, the *LPA* and LDL-C GRS contained the joint set of 198 SNPs that were used to construct univariable *LPA* GRS (29 SNPs) and univariable LDL-C GRS (169 SNPs). Specifically, for the *LPA* GRS, the 29 SNPs had the same conditional SNP weights as used in the univariable *LPA* GRS, whereas the remaining 169 SNPs had their SNP weights extracted from the marginal effect size estimates of the Lp(a) GWAS. For the LDL-C GRS, all 198 SNPs had their SNP weights extracted from the marginal effect size estimates of the LDL-C GWAS. We illustrate the genetic instruments selection scheme for Lp(a) and LDL-C in Supplementary Figure 8, Panel A.

In the multivariable MR of Lp(a) with apoB, the *LPA* and apoB GRS contained the joint set of 203 SNPs that were used to construct univariable *LPA* GRS (29 SNPs) and univariable apoB GRS (174 out of 175 SNPs, since one SNP [rs10455872] is duplicated with the *LPA* GRS instruments). Specifically, for the *LPA* GRS, the 29 SNPs had the same conditional SNP weights as used in the univariable *LPA* GRS, whereas the remaining 174 SNPs had their SNP weights extracted from the marginal effect size estimates of the Lp(a) GWAS. For the apoB GRS, all 203 SNPs had their SNP weights extracted from the marginal effect size estimates of the apoB GWAS. We illustrate the genetic instruments selection scheme for Lp(a) and apoB in Supplementary Figure 8, Panel B.

.

**Likelihood Ratio Test**

Of those 25 binary diseases/traits identified as potentially being the causal consequence of genetically-instrumented elevated Lp(a) from the univariable 2SLS MR, we next performed likelihood ratio tests (LRT) to quantify whether genetically predicted Lp(a) imposed similar disease risk for non-MACE endpoints compared to LDL-C and apoB, respectively. For the pairwise comparison between Lp(a) vs LDL-C on a given binary phenotype, we ran a logistic regression model under the null hypothesis that the *LPA* and LDL-C GRS had the same effect size:

$$log(\frac{P}{1-P}) = X\alpha + (G_1 + G_2)\beta \qquad (H_0),$$

and a logistic regression model under the alternative hypothesis that the *LPA* and LDL-C GRS had different effect sizes:

$$log(\frac{P}{1-P}) = X\alpha + G_1\beta_1 + G_2\beta_2 \qquad (H_1),$$

where $X$ denotes the design matrix of adjusting covariates (age, sex, PCs and genotyping platforms), $G_1$ denotes the *LPA* GRS and $G_2$ denotes the LDL-C GRS. We constructed the likelihood ratio statistics as -2 × $[log\ likelihood(H_0) - log\ likelihood(H_1)]$, and obtained the LRT p-value against the chi-square distribution with 1 degree of freedom. The pairwise comparison between Lp(a) vs apoB on a given phenotype was performed with the same procedure, with $G_2$ representing the apoB GRS instead.

The LRT results (Supplementary Table 6) suggested that among the 18 cardiovascular related diseases, genetically-instrumented Lp(a) had similar effect sizes compared to genetically-instrumented LDL-C and apoB, with the exception of peripheral artery disease, aortic stenosis and dilated cardiomyopathy, where Lp(a) had significantly larger effect sizes than LDL-C and apoB. Among the 5 risk factors for CVD, the *LPA* GRS in general had similar effect sizes with the LDL-C and apoB GRS on high blood pressure, but the LDL-C and apoB GRS had significantly different effect sizes than the *LPA* GRS on high cholesterol, high LDL and low HDL. LRT also suggested statistical difference between the risk of Alzhemir's disease family history among genetically-instrumented lipid traits (i.e. Lp[a] vs LDL-C, Lp[a] vs apoB, both with LRT p-values < 0.03), for which only Lp(a) had suggested provisional causal relationships.

To assess whether Lp(a) confers an effect on disease risk that is invariant to the LDL-C/apoB levels, we ran an interaction model to verify such a hypothesis. Specifically, we ran a logistic regression model with both the main effects and interaction effect included:

$$log(\frac{P}{1-P}) = X\alpha + G_1\beta_1 + G_2\beta_2 + (G_1 * G_2)\beta_3$$

where $X$ denotes the same design matrix of adjusting covariates as described above, $G_1$ and $G_2$ denote the main effects for *LPA* and LDL-C GRS, respectively, and $G_1 * G_2$ denotes the interaction effect for *LPA* and LDL-C GRS. The interaction model between Lp(a) vs apoB on a given phenotype was performed using the same procedure, with $G_2$ representing the apoB GRS instead. Among those 25 diseases/traits for which Lp(a) had a suggestive causal relationship with, none of them had a nominally significant interaction term (i.e. $\beta_3$ had a p-value > 0.05 in the interaction model for each of the 25 diseases/traits), suggesting that Lp(a) had an additive effect on the disease outcome which would not vary according to the LDL-C/apoB levels.

**Colocalization Analysis**

To evaluate whether Lp(a) and disease outcomes shared a single causal variant in the *LPA* locus, we performed colocalization analysis using the "coloc" package v5.1.0 in R v3.6.2. We evaluated whether Lp(a) levels shared the same causal variant with each of the 25 diseases / traits from the provisional MR-PheWAS findings. We used the "coloc.abf" function with its default parameter values to perform the colocalization analysis. We considered the posterior probability (PP) of a shared causal variant with PP > 0.8 as evidence of colocalization.

The colocalization algorithm assumed only one single causal variant exists for any given trait in any genomic region (Wallace, C., 2013). Giambartolomei, C. *et al.*, (2014) proposed the conditional leave each out (CLEO) framework to isolate and refine independent signals from a locus. The CLEO algorithm aimed to handle multiple independent associations in a locus by separating the statistical support for each variant conditional on the remaining causal signals being considered, in order to achieve more accurate coloc inference. We ran CLEO analysis on both UKB Lp(a) levels and 23andMe disease outcomes in the *LPA* region, and used the CLEO summary statistics to conduct the coloc analysis. In summary, for each of the 25 disease outcomes, we first performed a stepwise conditional analysis beginning with the index variant in the genomic region, and continued with the forward selection procedure to add additional significant lead variants one at a time. After the conditional (i.e. forward selection) step-down analysis, we applied CLEO for each lead variant identified in the conditional step-down, leaving them out one at a time and recomputing the association test statistics for all variants in the locus using the remaining lead variants as covariates. For Lp(a) concentrations, we skipped the forward selection/ step-down procedure and used the 29 genetic instruments returned by SuSiE credible sets (Supplementary Table 2), followed by CLEO to obtain the conditional summary statistics for each of those 29 lead SNPs.

For a given pair of Lp(a) versus disease outcome, we used all possible combinations of the Lp(a) CLEO summary statistics of each step versus the disease outcome CLEO summary statistics of each step as input data to conduct the colocalization analysis. In each Lp(a) versus disease outcome pair, we ranked the colocalization posterior probability (PP) across all possible combinations of CLEO steps, and assessed whether the highest colocalization PP showed strong evidence (with PP > 0.8) that a particular disease outcome shared the same underlying causal variant with Lp(a) in the region, to provide additional evidence in supporting the MR assumptions.

In summary, among the 18 cardiovascular related diseases, colocalization signals were observed on most of the Lp(a) vs disease pairs, except for transient ischemic attack and dilated cardiomyopathy. We also observed colocalization analysis among all five risk factors for CVD, and the family history of Alzheimir's disease. Colocalization between Lp(a) and prostate cancer suggested that they have different causal variants in the region (Supplementary Table 7; Supplementary Figure 9.1 - 9.25).

**Univariable Mendelian Randomization Sensitivity Analysis**

Besides the primary 2SLS approach with the individual-level data, we performed additional univariable two-sample MR analyses (2SMR) using the summary-level statistics to verify the robustness of our findings. Specifically, we applied inverse variance weighting (IVW), MR-Egger and weighted median methods as a set of complementary sensitivity analyses. To assess the instrumental variable assumptions, we used Cochran's Q-statistics to evaluate heterogeneity and MR-Egger intercept to evaluate directional horizontal pleiotropy. To improve validity of the MR inferences, we further applied heterogeneity and Steiger filtering to remove variants that were deemed violating the model assumptions and re-estimated the casual estimates on the remaining set of SNPs. We implemented those 2SMR analyses in the "TwoSampleMR" package v0.56 in R v3.6.2, with the exposure summary statistics extracted from the UKB Lp(a), LDL-C, or apoB genetic association analysis, and the disease outcome summary statistics extracted from the 23andMe GWASs.

The IVW, MR-Egger and weighted median methods using the full sets of SNPs (i.e. 29 for Lp(a), 169 for LDL-C and 175 for apoB) all yielded similar MR estimates compared with the corresponding 2SLS MR estimates (Supplementary Table 8, 9, 10). Intercepts from MR-Egger suggested no evidence of directional horizontal pleiotropy for the Lp(a) MR inference among all 25 phenotypes (p-value > 0.05; Supplementary Table 11), and no evidence of directional horizontal pleiotropy for most of the LDL-C and apoB MR inference, with the exception of dilated cardiomyopathy, high cholesterol, high LDL and high blood pressure (Supplementary Table 12, 13). Cochran's Q statistics suggested evidence of effect size heterogeneity for all three lipid biomarkers across the 25 phenotypes (Supplementary Table 14, 15, 16). Therefore, we applied heterogeneity and Steiger filtering to each lipid biomarkers' genetic instruments, which removed around 50% of the SNPs on average for each biomarker (Supplementary Table 17, 18, 19). The 2SMR analysis on the remaining set of SNPs yielded directionally consistent effects compared with using the full set of SNPs, albeit larger standard errors and p-values due to reduced power (Supplementary Table 17, 18, 19; Supplementary Figure 10.1.1-10.25.2, 11.1.1-11.25.2, 12.1.1-12.25.2). We note that with the SNP filtering, Lp(a) no longer had a significant MR signal with prostate cancer from IVW, MR-Egger and weighted median (Supplementary Table 17; Supplementary Figure 10.24.2).

**Multivariable Mendelian Randomization Sensitivity Analysis**

We performed additional multivariable two-sample MR analyses (2SMR) with the summary-level statistics to assess the MR assumptions. Specifically, we used the conditional F statistics to assess the instrumental variable strength and conditional cochran's Q statistics to assess pleiotropy. In the multivariable MR with Lp(a) and LDL-C, conditional F statistics for both lipids were much larger than 10 (conditional F = 583.441 for Lp[a], and 181.129 for LDL-C), suggesting strong instrument strength for both exposures. The conditional Cochran's Q statistics indicated evidence of horizontal pleiotropy (Q statistics = 604.959, p-value = $8.798 \times 10^{-45}$). Thus, we used the Q minimization approach (Sanderson, E., Spiller, W. and Bowden, J., 2021) to obtain the direct causal effect estimates rather than the IVW approach, which yielded an OR = 1.077 for Lp(a) and OR = 1.103 for LDL-C. Similarly, in the multivariable MR with Lp(a) and apoB, conditional F statistics suggested strong instrument strength for both lipids (conditional F = 580.016 for Lp[a], and 223.851 for apoB), and conditional Cochran's Q statistics suggested evidence of horizontal pleiotropy (Q statistics = 613.473, p-value = $1.525 \times 10^{-44}$). The direct causal effect estimates obtained from the Q minimization approach yielded an OR = 1.075 for Lp(a) and OR = 1.138 for apoB.

**Deriving an Estimate of Lp(a) Lowering Required to Achieve a CVD Risk Reduction Equivalent to that of LDL-C**

It is of value to identify the value by which Lp(a) might need to be lowered in order to derive a clinical effect comparable to statins. Large-scale randomized clinical trial evidence demonstrates that a 1 mmol/L (38.67 mg/dL) reduction in LDL-C through statin treatment leads to a 25% reduction in the relative risk of major vascular events (Collins, R. *et al.*, 2016). Our finding showed that a *LPA* GRS scaled to a difference in Lp(a) of 59.632 nmol/L (equivalent to 27.354 mg/dL) led to a genetically-predicted equivalent causal effect on MACE as a LDL-C GRS scaled to a 0.298 mmol/L (equivalent to 11.524 mg/dL) difference in LDL-C. In other words, using the same units (mg/dL), in order to achieve a 25% relative risk reduction (RRR) of CVD, a 27.354/11.524*38.67 lowering of Lp(a) would be required, which is 91.79 mg/dL.
