## Supplementary Tables for "Evaluating genetically-predicted causal effects of lipoprotein(a) in human diseases: a phenome-wide Mendelian randomization study": Supplementary_Table_1.docx

**Supplementary Table 1: Pairwise phenotypic and genetic correlation between Lp(a), LDL-C and apoB in the UKB cohort.** (A) pairwise Pearson correlation coefficients between measured Lp(a), LDL-C and apoB; (B) pairwise cross-trait LD Score (LDSC) regression estimates between Lp(a), LDL-C and apoB GWAS; (C) pairwise Pearson correlation coefficients between *LPA*, LDL-C and apoB GRS.

| (A) | Pearson correlation based on measured values in UKB | | |
| --- | --- | --- | --- |
|  | Lp(a) | LDL-C | apoB |
| Lp(a) | 1 | 0.111 | 0.112 |
| LDL-C | 0.111 | 1 | 0.958 |
| apoB | 0.112 | 0.958 | 1 |

| (B) | LDSC based on UKB GWAS summary statistics | | |
| --- | --- | --- | --- |
|  | Lp(a) | LDL-C | apoB |
| Lp(a) | 1 | 0.206 | 0.193 |
| LDL-C | 0.206 | 1 | 0.946 |
| apoB | 0.193 | 0.946 | 1 |

| (C) | Pearson correlation based on UKB GRS | | |
| --- | --- | --- | --- |
|  | *LPA* | LDL-C | apoB |
| *LPA* | 1 | 0.065 | 0.057 |
| LDL-C | 0.065 | 1 | 0.915 |
| apoB | 0.057 | 0.915 | 1 |
